# Multi-disease risk prediction models for population-scale personalised screening

**DOI:** 10.64898/2026.02.02.26345271

**Authors:** RR Oexner, R Schmitt, M Kals, H Ahn, SS Khawaja, D Biswas, RA Shah, P Chowienczyk, A Zoccarato, P Palta, K Theofilatos, AM Shah

## Abstract

An increased emphasis on disease prevention is essential to improve population health and reduce healthcare resource needs. Key requirements for effective population scale prevention programmes are an optimal balance between simplicity of screening and accuracy of risk prediction at individual level. We developed models of varying complexity to predict the incident risk of 24 diseases in the UK Biobank population, testing combinations of modalities including established clinical variable-based risk scoring, ^1^H-NMR metabolomics, polygenic risk scores (PRS), and self-answered questionnaires with individual electronic health record (EHR)-based past medical history (PMH). Our results show that prediction models that utilise just questionnaire and PMH data (“No Needle” model) or with added metabolomics and PRS (“Single Blood Draw” model) exhibit robust discriminative performance, at least as good or better than a comprehensive clinical variable model or established cardiovascular disease (CVD) risk scores. The “No Needle” model was also validated in an independent prospective cohort, the Estonian Biobank. We also tested scenarios for deployment of these models to improve the effectiveness of the NHS Health Check, a population-scale UK screening programme. Our results suggest high potential for less resource-intensive approaches than current clinically-based paradigms for prediction of incident disease.

---

Chronic diseases impose an increasingly unsustainable burden on healthcare systems worldwide, especially related to multimorbid ageing populations.^1^ Many of the most common conditions, such as cardiovascular disease (CVD), dementia, cancer and other age-related conditions, are potentially preventable by simple lifestyle changes (e.g. smoking cessation, diet) or inexpensive pharmacotherapy (e.g. statins) or are amenable to effective treatment with earlier diagnosis.^2–7^ Current prevention programmes, however, are limited in their population reach and effectiveness. Measures such as public health messaging are untargeted while screening programmes are typically based on simple cut-offs (e.g. age) and have variable take-up among the general population who consider themselves healthy.^8^ Moreover, most screening programmes focus on single diseases (e.g. breast cancer) whereas chronic and often lifestyle-related conditions increasingly occur in combination. There is therefore an imperative to develop more effective risk prediction and early screening approaches that accurately identify high-risk individuals at population scale and facilitate implementation of person-targeted prevention programmes with higher impact on population health and healthcare resource requirements.

Different approaches have been suggested to guide population-scale stratification based on predicted risk of incident disease. The current conventional approach is actively healthcare professional driven clinical risk scoring involving a combination of demographic, physical measurement, clinical chemistry, interview and PMH data. In general, this depends upon individuals periodically attending an in-person visit with a healthcare professional and is typically restricted to a few common conditions.^9–11^ Proposed approaches to improve risk scoring include the incorporation of additional predictors or more complex analytical approaches.^12–14^ More recently, several studies have suggested the potential for multi-disease risk prediction through deep profiling of biomarkers such as metabolomics or PRS.^15–17^ Nevertheless, a comparative assessment of different potential screening modalities in combinatorial scenarios that are actionable at population scale is lacking.

In this study, we developed and tested different risk prediction models across a multi-disease spectrum, with particular focus on the assessment of a range of scalable scenarios that would be suitable for population-level disease screening. Our results indicate that data that are in principle collectable in non-healthcare community settings (self-completed questionnaires, self-collected blood sample) provide robust discriminative performance. We also model how such approaches could be integrated with established conventional screening programmes such as the UK NHS Health check, to enhance overall effectiveness. Taken together, our work demonstrates the potential for population-scale community-based screening approaches that would facilitate prevention programmes targeted to high-risk individuals.

## Results

We analysed prospective data from the UK Biobank study for the incidence of 24 conditions across a multi-disease spectrum (see Online Methods), including CVD, cancer and dementia diagnoses and several common diseases with existing actionable preventive measures (Supplementary Table 1).^15,16,18^ We developed and tested the performance of several different risk prediction models (Figure 1). As a clinical benchmark for multi-disease risk (“Clinical” model), we used a previously described score that incorporates multiple predictors across demography, lifestyle, PMH, physical measurements, clinical chemistry and medication.^14^ A second approach utilised baseline data from self-answered standardised touchscreen questionnaires completed by all UK Biobank participants and baseline PMH extracted from their EHR (“No Needle Model”). The questionnaires span domains including sociodemographics, lifestyle and environment, early life, family history, psychosocial factors, health and medical history, and sex-specific factors. A third approach added metabolomics and PRS data, which can in principle be assayed from a single venesection, to the “No Needle Model” (“Single Blood Draw Model”). We used the UK Biobank “Standard PRS” set^18^ as a uniform multi-PRS predictor matrix while for metabolomics we used the full available 168 metabolite dataset^16^. All participants with available baseline ^1^H-NMR measurements and PRS were included while those with a baseline diagnosis of any of the 24 conditions were excluded (final cohort size *n* = 239,207). The dataset was randomly split into 50% training and test partitions (Table 1). Unless stated otherwise, all subsequent results were obtained by evaluating the UKB test split. The approaches used for data missingness, imputation and post-imputation processing are described in the Online Methods. We trained elastic net (EN)-penalized Cox proportional hazards (Cox-PH) models for each unique predictor and endpoint combination, then assessed discriminative performance via derivation of Harrell’s C (see Online Methods for detailed model training approach; Supplementary Tables 2-5 for comprehensive results).

**Figure 1:**
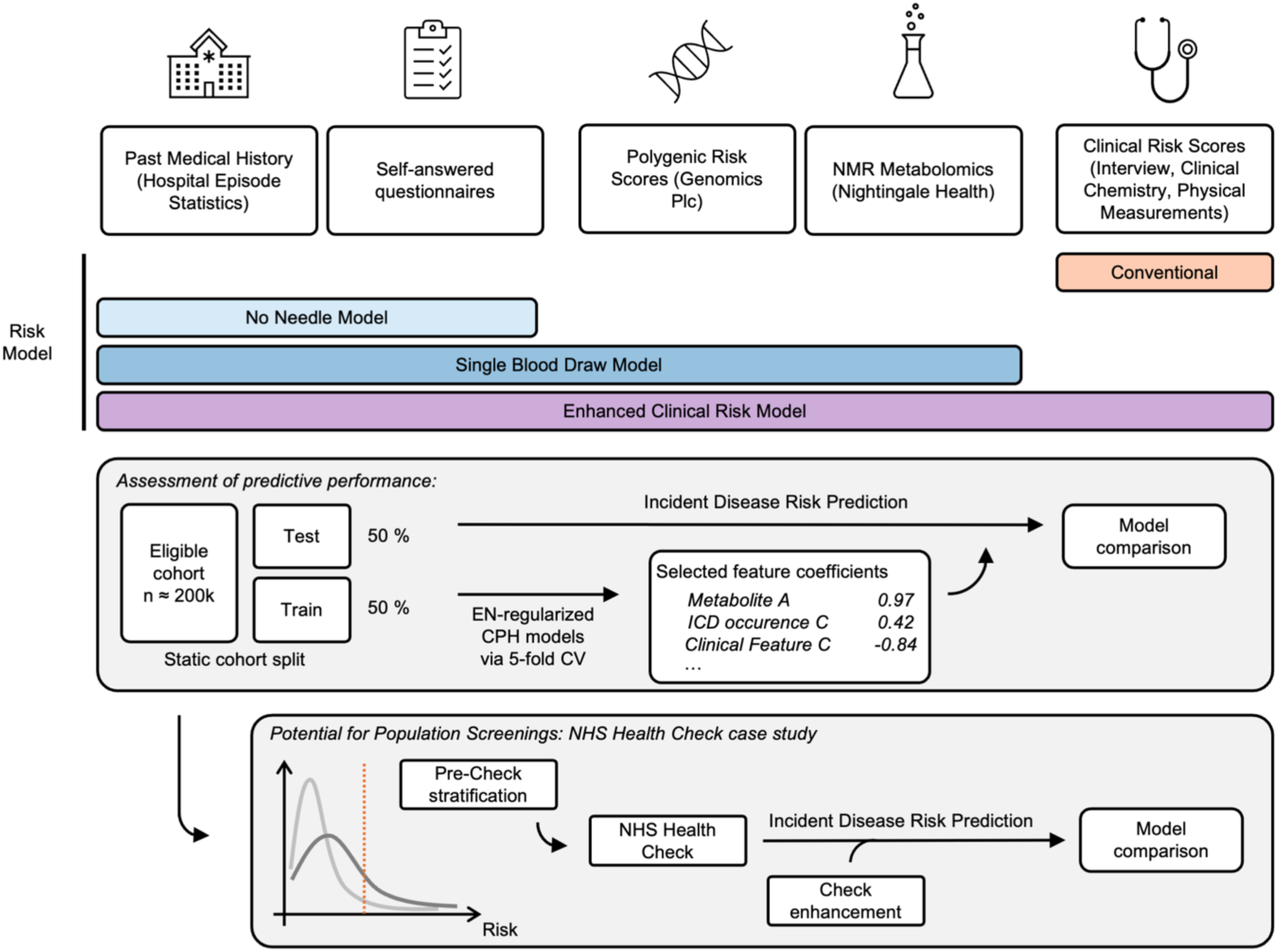
Study Overview: Within this study, we compared combinations of different predictor modalities including readily available and self-reported data points. Models were trained on a 50% training partition and evaluated on the unseen, 50% test split. We also assessed how the NHS Health Check might be improved through (I) “No Needle Model”-driven, remote pre-check stratification and subsequent invitation of a high-risk subset only and/or (II) enhancement of the check via additional incorporation of different predictive assays. NMR: proton nuclear magnetic resonance, CPH: Cox proportional hazard, NHS: National Health Service.

**Table 1:**
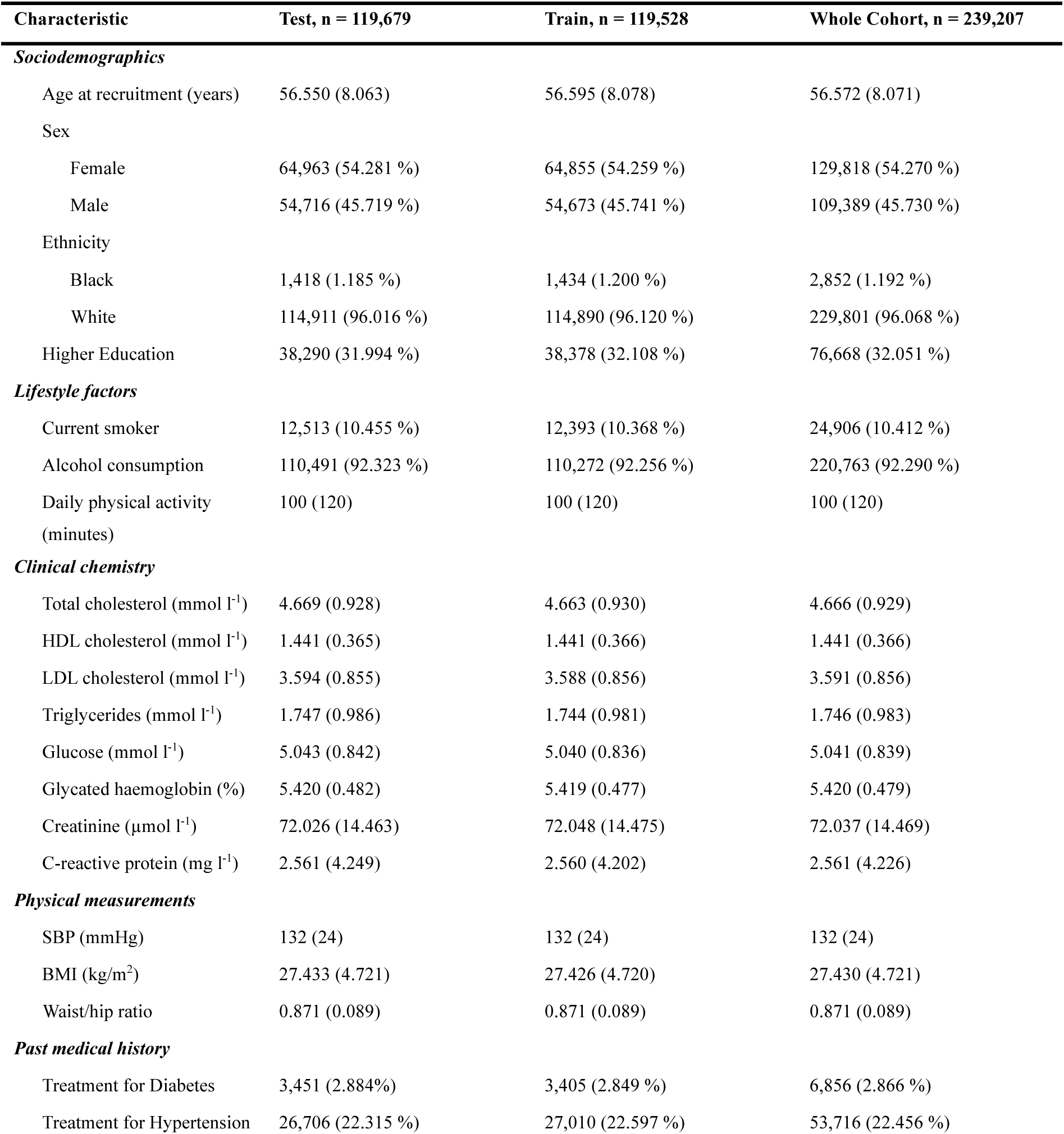
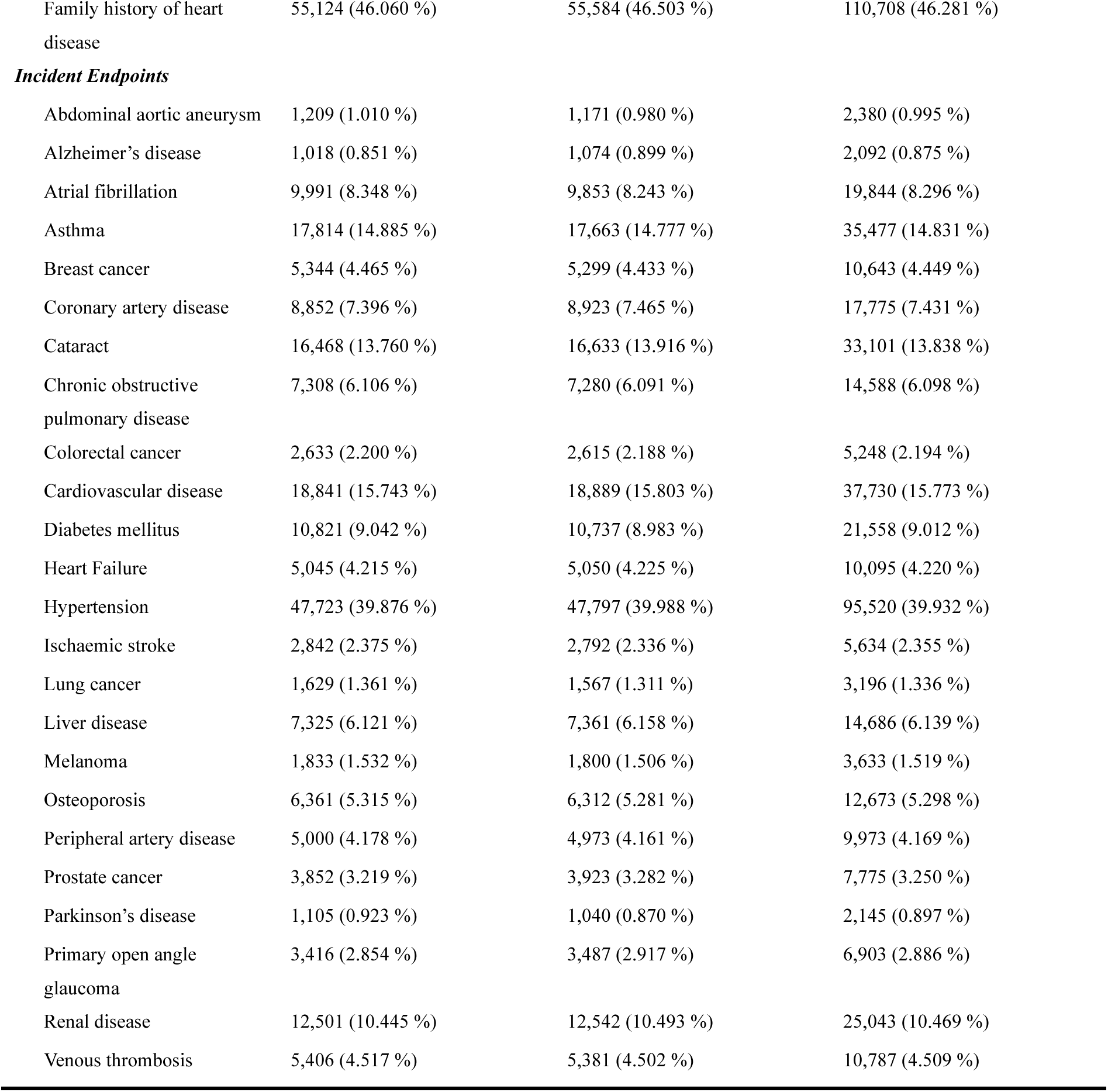
Baseline characteristics: The table details baseline characteristics for the training and test partitions, including incident endpoint frequencies. Assignment to training and test splits was randomized, which is mirrored in near-identical cohort characteristics.

The addition of Questionnaire/PMH data to the “Clinical” model significantly improved performance compared to the clinical benchmark for 18 of 24 endpoints, while also incorporating PRS and metabolomics improved performance for all 24 endpoints (Fig. 2A, left 2 panels). In several instances, for example for some cancers or asthma, the improvement was substantial (ΔC > 0.05 over the clinical benchmark). Next, we assessed the performance of the “No Needle” and the “Single Blood Draw” models, which can potentially be deployed in non-healthcare community settings, as compared to the clinical benchmark (Fig. 2A, right 2 panels). We observed very robust discriminative performance of the “No Needle” model which at least matched the “Clinical” model for 21 of 24 endpoints and showed significant improvement for 10 endpoints. The “Single Blood Draw” model significantly outperformed the clinical benchmark for 20 of 24 assessed endpoints. Fig. 2B displays the risk stratification by risk tertile for each of the 24 endpoints when using the “No Needle” model.

**Figure 2:**
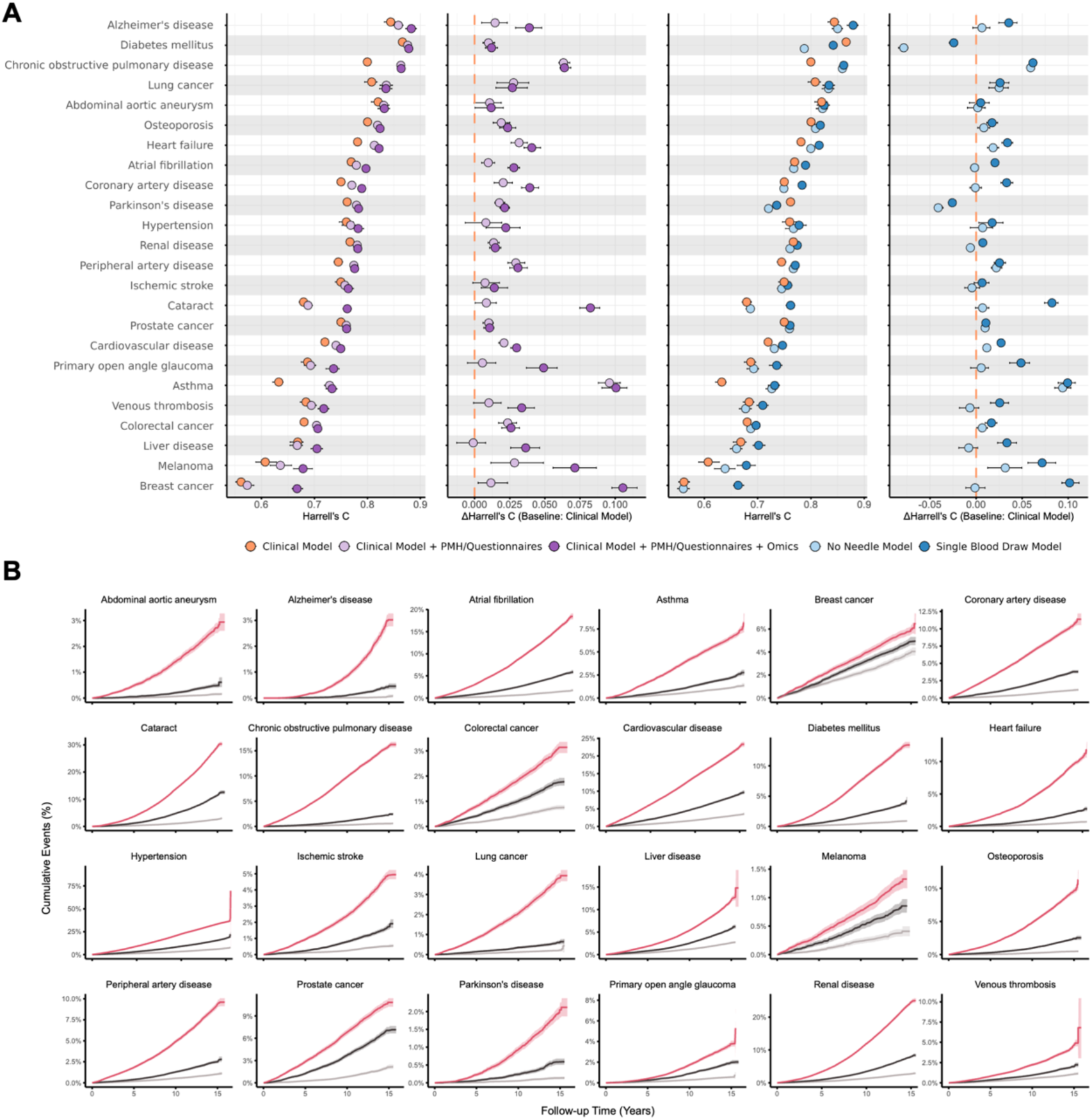
Comparison of discriminative performance across a multi-disease spectrum: Absolute Harrell’s C (Panel A, left sides) and ΔC (Panel A, right sides; benchmark “Clinical Model”) across endpoints, for different risk modelling strategies: a clinical benchmark with/without additional incorporation of past medical history (PMH), self-answered questionnaires, polygenic risk scores (PRS) and metabolomics (left panels), or the isolated usage of such assays (PMH/Questionnaires, “No Needle Model” or PMH/Questionnaires + Omics, “Single Blood Draw Model”). Further details shown in Figure 1 and *Methods*, Table 2). Error bars represent bootstrapping-derived 95% confidence intervals. Panel B shows risk tertile (red: highest risk, dark grey: intermediate risk, light grey: low risk) stratification for the “No Needle Model” scenario, with subplots corresponding to the respective endpoints.

Among the endpoints that display good discriminative performance with the “No Needle” and “Single Blood Draw” models are multiple CVD endpoints for which there are well established risk scoring strategies, such as SCORE2, PREVENT and QRISK3.^9–11^ We extracted the predictors for these risk scores in our study population (Figure 3A) and compared performance against the “No Needle” and “Single Blood Draw” models. Of the three established risk scores, QRISK3 is the most extensive, was developed for the UK population, and showed the best overall performance in the UK Biobank cohort (Figure 3B). However, the “No Needle” model showed better performance than QRISK3 for 3 of the 5 endpoints and matched it for the other 2 endpoints (Figure 3B). The “Single Blood Draw” model provided improved discrimination across all 5 tested cardiovascular endpoints. Consistent with these findings, both receiver operating curve (ROC) and decision curve analysis (DCA) plots demonstrated similar results (Figure 3C and D, respectively).

**Figure 3:**
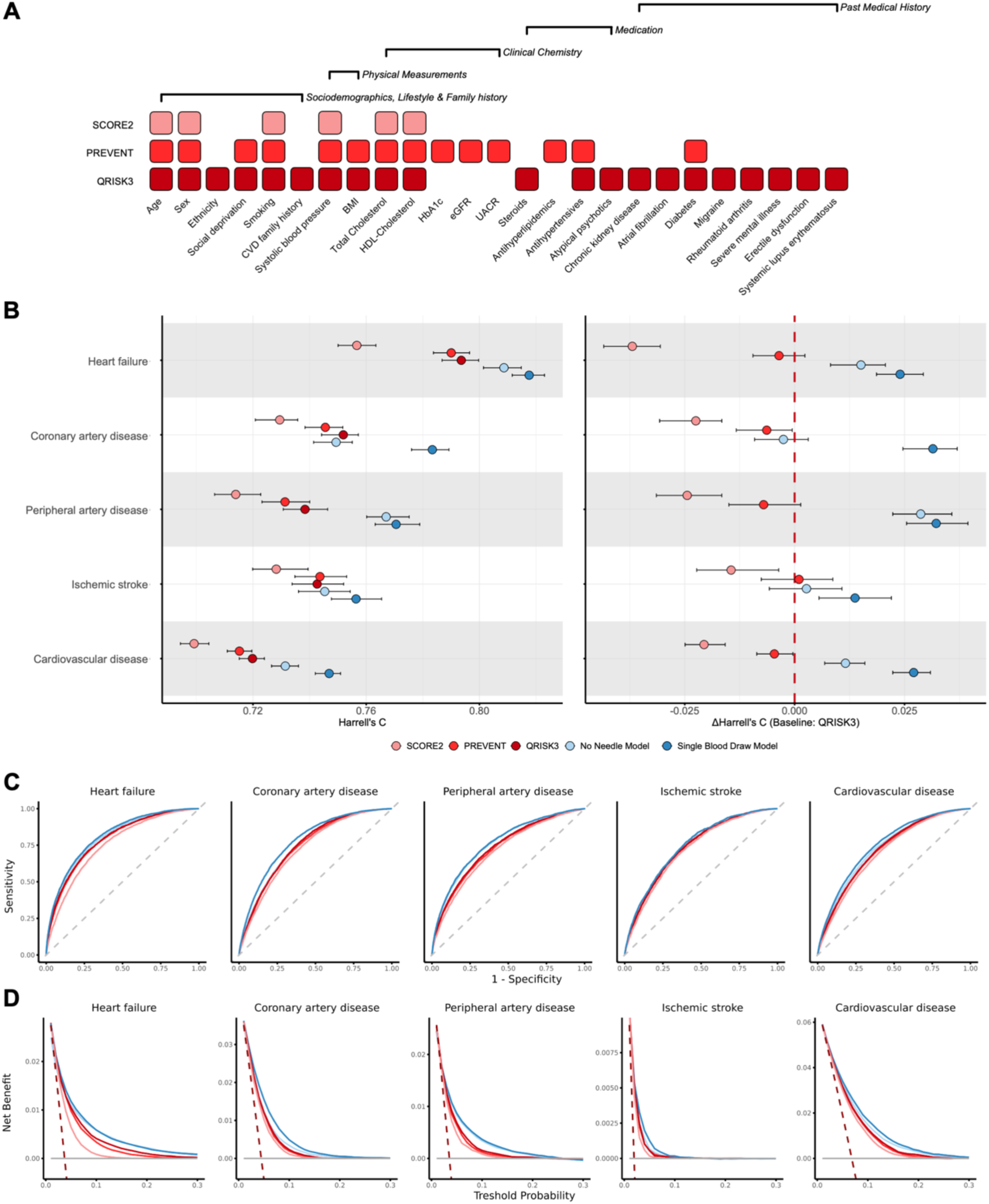
Cardiovascular disease risk stratification: We compared potentially remote models (“No Needle Model” and “Single Blood Draw Model”; for details: Figure 1 and *Methods*, Table 2) against three cardiovascular risk scores (SCORE2, PREVENT and QRISK3; Panel A) for both absolute Harrell’s C (Panel B, left side) and ΔC (Panel B, right side; benchmark “QRISK3”) across a range of cardiovascular disease spectrum endpoints. Error bars represent bootstrapping-derived 95% confidence intervals. Receiver operating characteristic (ROC) (Panel C) and decision curve analysis (DCA) (Panel D) plots are also shown.

To validate our approach to incident risk prediction in an independent prospective population, we analysed data from 49,620 first-phase participants in the Estonian Biobank cohort, which has similar baseline self-completed questionnaire data^19^. We found that the “No Needle” model demonstrated very good discriminative performance for CVD endpoints as compared to established risk scores (Supplementary Fig. 1, Supplementary Table 6).

We next analysed which of the main predictor modalities (clinical, questionnaires/PMH, metabolomics, PRS; Supplementary Table 5 for per-predictor-matrix-performance) contributed most to model performance for the different endpoints, by summing up absolute coefficient values (Fig. 4A). The results indicate consistently large contributions of the self-answered questionnaires for the majority of disease endpoints including CVD. Other approaches contributed in a largely endpoint-specific manner (e.g. genetic risk was relatively important for the prediction of breast or colorectal cancer). We further investigated the contribution of self-answered questionnaires to multi-disease risk score by question domain. This analysis indicated that questions across all main domains contributed to the risk score for most disease endpoints, but the relative contribution of questionnaire domains was heterogeneous across endpoints (Fig. 4B).

**Figure 4:**
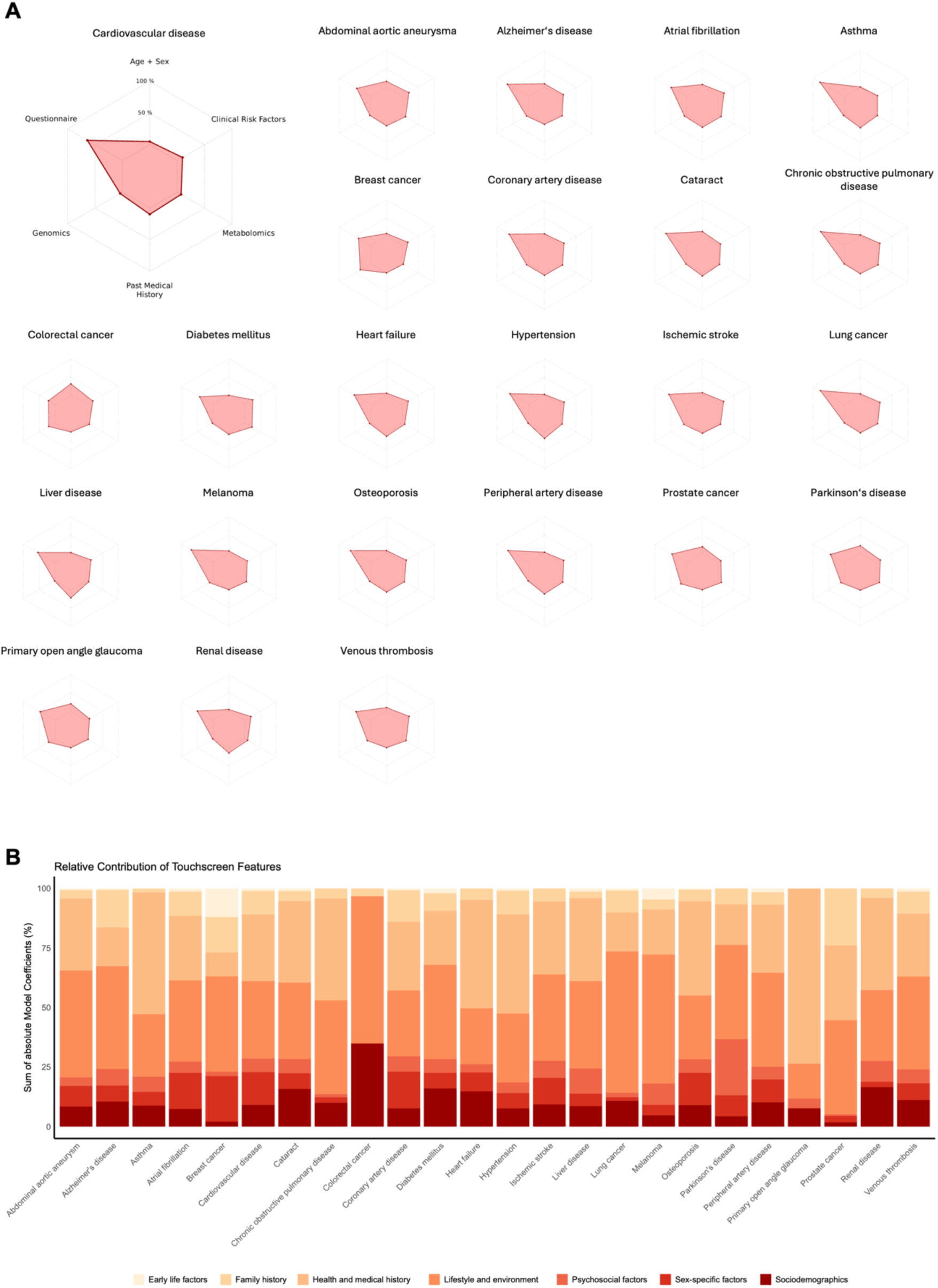
Feature importance: Contributions of predictor modalities are illustrated in radar plots showing the relative summed absolute coefficients (Panel A). A similar approach was taken to assess the contribution of self-answered questionnaire subcategories (Panel B). Coefficients were extracted from the “Clinical Model + PMH/Questionnaires + Omics” model, i.e. the model utilising all benchmarked input features.

With the idea that self-answered questionnaires and self-collected blood samples for ‘omics can in principle be obtained in non-healthcare community settings, we modelled how their use might enhance population-level screening programmes. We based our modelling on the NHS Health Check, a UK public health screening programme for healthy, middle-aged individuals to facilitate primary prevention of CVD, kidney disease (KD) and diabetes mellitus (DM). We compared the current approach of inviting all individuals aged 40-74 for an in-person check (“1-step NHS Health Check”; see Online Methods) versus additional incorporation of PMH and self-answered questionnaires (“1-step NHS Health Check + PMH/Questionnaires”). We also modelled pre-Check risk stratification based on the “No Needle” model to identify individuals with >5% predicted 10-year risk for any NHS Health Check endpoint, followed by an in-person Health Check of only the high-risk subset, with or without additional incorporation of PRS and ^1^H-NMR metabolomics (“2-step NHS Health Check”, “2-step NHS Health Check + Omics”) – Figure 5A. The incorporation of self-answered questionnaires/PMH data to the standard NHS Health Check improved discriminative performance versus the standard NHS Health Check (Figure 5B): KD ΔC = 0.025 [95% CI 0.014 – 0.036], DM ΔC = 0.017 [95% CI 0.006 −0.026], and CVD ΔC = 0.015 [95% CI 0.008 −0.021]. Applying the “No Needle” model to the UK Biobank population indicated that a substantial proportion of people had a < 5% predicted 10-year risk of DM, KD or CVD (Figure 5C, upper panel). Moreover, we display excellent model calibration across endpoints (per decile; Figure 5C, upper panel). Applying the 2-step NHS Health Check model resulted in significantly improved risk prediction in this group for KD and comparable performance for CVD but worse performance for DM as compared to the 1-step NHS Health Check (Fig. 5C). The addition of ‘omics to a 2-step strategy resulted in further improvement in prediction of CVD.

**Figure 5:**
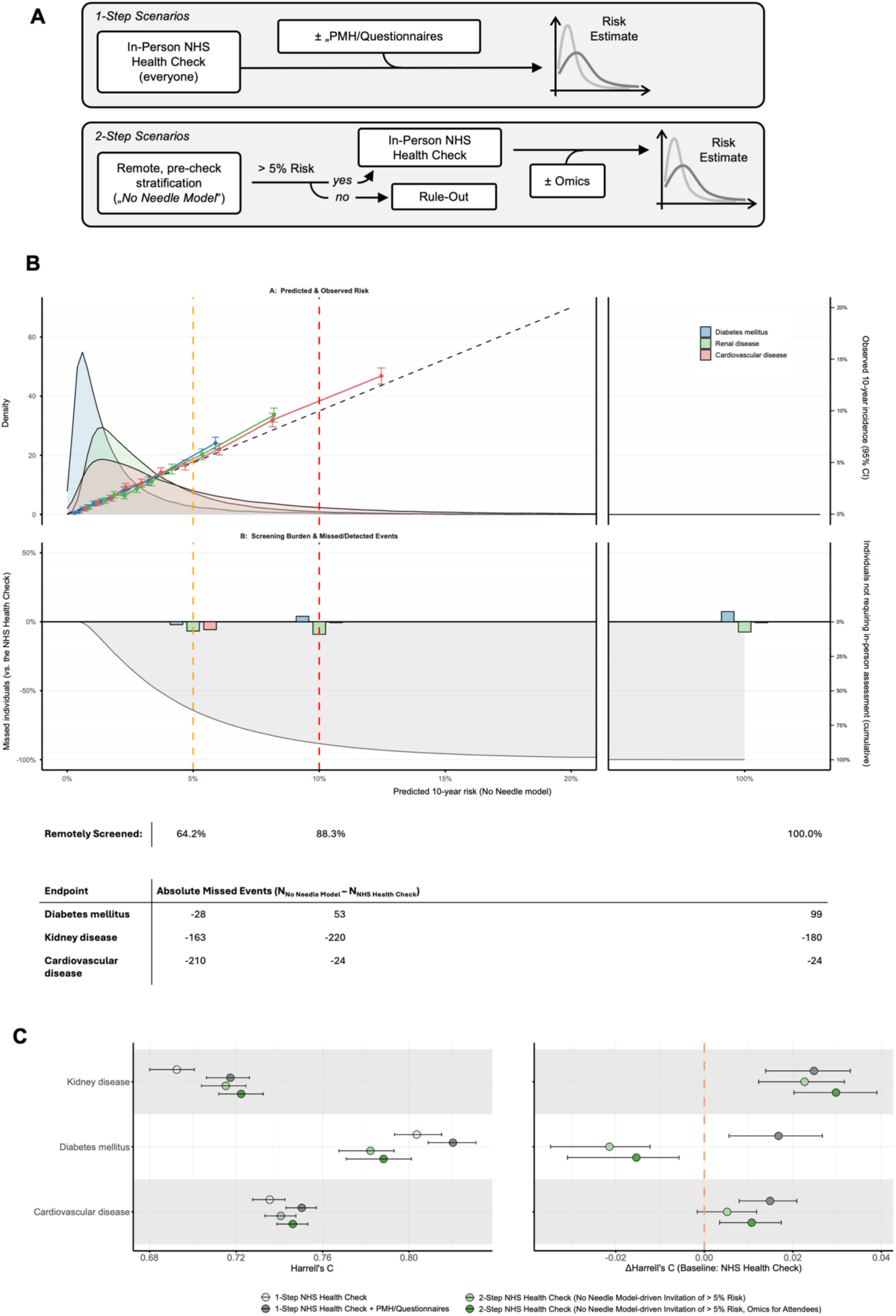
Modelling the NHS Health Check (NHS-HC): We defined different scenarios within the existing NHS-HC framework, where for 2-Step models, absolute 10-year risk predictions were used to determine whether in-person attendance (i.e., performing additional NHS-HC measurements) is required (Panel A; for details: Figure 1, Methods, Table 2). Models were benchmarked against the existing state-of-the-art (“1-Step NHS Health Check”) for both absolute Harrell’s C (Panel B, left side) and ΔC (Panel B, right side) across the three endpoints central to the NHS-HC (kidney disease, diabetes mellitus and cardiovascular disease). Error bars represent bootstrapping-derived 95% confidence intervals. Within Panel C, we additionally model the consequences of the remote pre-screening-pathways: the upper calibration plot displays predicted 10-year risk vs observed 10-year incidence for each endpoint, across deciles (error bars represent 95% confidence intervals derived from Kaplan–Meier estimates), with endpoint-specific risk distributions overlaid. The lower panel summarises the screening burden trade-off: the proportion of individuals who would not require an in-person National Health Service (NHS) Health Check (“remotely screened out”) at each threshold, alongside the change in missed vs detected events compared with the standard NHS-HC strategy. Vertical dashed lines indicate example referral risk (5 and 10%) thresholds. The table reports both the number of fully remotely screened individuals and the absolute differences in missed events at the illustrated risk thresholds. PMH: past medical history; CI: confidence interval.

Finally, we assessed the trade-off between the screening workload and detection of at-risk individuals as the referral threshold is varied, reporting both the proportion “remotely screened out” and the change in missed (vs. detected) events relative to the standard (1-step) NHS Health Check. At a referral threshold of > 5% predicted 10-year risk, 64.2% of individuals would be screened out remotely, whereas at a 10% threshold this increases to 88.3% (Fig. 5C). The overlaid bar plots and the table in Fig. 5C report the corresponding absolute differences in missed events for each endpoint at these thresholds, highlighting how tightening the threshold reduces in-person checks without necessarily reducing event capture (for example, if inviting at a 5% threshold, we would detect more cases for each of the three assessed endpoints).

## Discussion

Stratification of the general population by incident risk of major chronic diseases is a vital component of strategies to implement effective primary prevention efforts and reduce long-term healthcare system costs. Key considerations in the choice of different risk prediction approaches^20^ that could be deployed include the accuracy of stratification, the logistics of screening procedures, the extent to which the target population engages with the programme, and the range of chronic diseases that are covered – which collectively contribute to programme cost-effectiveness, efficacy and safety. With these factors in mind, in the current study we wished to test multimodal approaches of varying logistical complexity including the leveraging of person-provided digital information and ‘omics data obtained through increasingly simple sample collection and assay.^21^ Our major finding is that a screening approach based solely on self-answered questionnaire and EHR–based PMH (“No Needle” model) demonstrates robust discriminative accuracy for multi-disease risk prediction. Such information can in principle be collected in non-healthcare settings at relatively low cost and without direct clinical involvement, factors that are of particular interest given current challenges in healthcare infrastructure, workforce capacity^22^ and the low uptake of in-person prevention programmes.^23^ These results align with previous work showing that self-reported lifestyle information and EHR-derived predictors capture a broad range of latent health states and behaviours associated with chronic disease onset.^24,25^

We found that predictions based on the “No Needle” model in many cases matched or even exceeded the discriminative accuracy of a clinical risk scoring benchmark, which is probably too complex and costly for population-level deployment, or established cardiovascular risk scoring approaches such as QRISK3. External validation in the Estonian Biobank cohort confirmed that key findings translate across populations with distinct ancestry and healthcare structures, supporting the generalisability of our approach. Moreover, we found that the simple data contributing to the “No Needle” model can be sensibly combined with the more complex clinical score to unlock substantial discriminative accuracy gains across endpoints. It could also be leveraged for pre-screening or decentralised triage prior to more focused clinician-delivered screening, as modelled for the UK NHS Health Check programme.

An additional finding in this study was that the incorporation of readily scalable omics assays such as ¹H-NMR metabolomics and PRS (estimating inherited predisposition) provided discriminative improvement across nearly all cardiometabolic and neurodegenerative endpoints, consistent with prior findings that circulating metabolite profiles reflect both genetic and environmental disease determinants^15,26–28^ and that polygenic predisposition contributes predictive information beyond clinical risk factors.^16,29–31^ This detailed benchmarking of multiple risk prediction approaches in the current study provides a basis to design and test differently sequenced or combined multimodal models to determine the most effective population scale prevention programmes.

We also determined which features contributed to the predictive value of different models. For example, our most extensive model for CVD prediction (“Clinical Risk + PMH/Questionnaires + omics”) contained many expected features such as high coefficients for age, systolic blood pressure, self-reported family history of heart disease, and Genomics PLC’s standard coronary artery disease PRS. However, other features did not have an established *a priori* link to specific endpoints or flagged potentially missed diagnoses, e.g. the participant’s answer to the touchscreen questionnaire question “Do you ever have any pain or discomfort in your chest?”. This highlights the additional value of a systematic questionnaire-based approach.

The suggested “No Needle Model” strategy could in principle be employed via a web interface or an App and therefore be highly scalable, without the need for an in-person consultation. From a translational standpoint, we modelled how such risk stratification could be implemented as part of the UK NHS Health Check. Our results suggest significant potential for remotely conducted pre-check stratification followed by in-person screening for higher-risk population subsets. Further opportunities arise from the option to self-collect a blood sample for ‘omics profiling in a person’s home via a posted kit.^21^

Strengths of this study are the development of risk prediction models in a large cohort, international validation, improvement versus the current clinical gold-standard across a range of disease endpoints, and the modelling of added value to existing healthcare workflow. Our study also has some limitations. First, we relied for our predictive models on linear algorithms which may not capture non-linear relationships that might be present in such complex datasets. However, this facilitates interpretability in a setting where risk communication and patient trust are central. Moreover, such linear algorithms have been found to display robust performance in previous survival task benchmarking efforts.^32^ Secondly, we tested prediction for a selected range of conditions, including some that may have interlinked aetiology. Our aim was to provide proof-of-concept for a wide range of common conditions; future work can expand or modify the list of conditions depending on other considerations, such as cost-effectiveness. Thirdly, the UK Biobank cohort is recognised not to be fully representative of the general UK population but has the advantage of substantial size, depth of characterisation and prolonged follow-up. Fourthly, external validation of this work posed challenges related to predictor availability. Nevertheless, we were able to underscore the transferability and generalisability of key aspects of the current results via validation in the Estonian Biobank cohort. Finally, formal prospective testing and assessment of cost-effectiveness would be needed to translate this work into a public health programme.

In conclusion, this work demonstrates a clear opportunity to re-imagine preventive screening as a dynamic, potentially simple, community-based multi-disease programme that complements clinician-dependent personalised screening and intervention. By systematically comparing predictor modalities, we provide an evidence-based foundation for the testing of different multimodal risk prediction approaches for the incidence of multiple chronic diseases to reduce avoidable morbidity and mortality.

## Funding

RRO was supported by the British Heart Foundation (BHF) (FS/19/58/34895, RE25626) and UK Biobank “getting started” and “enhanced” credit programmes. MK and PP were supported by the European Union and Estonian Research Council through the Mobilitas 3.0 (MOB3JD1203) and by Estonian Research Council Grant PRG1291 (Systematic phenome-wide search for genetic modulators in health and disease). RAS was funded by a National Institute for Health and Care Research (NIHR) Academic Clinical Fellowship. AMS is supported by the BHF (CH/1999001/11735, RG/20/3/34823, RE/18/2/34213) as are KT (PG/20/10387, RE25626) and AZ (PG/24/11770).

## Conflicts of interest

D.B. has stock options in Ataraxis AI. D.B. is an inventor on an international patent application relating to methods for predicting survival rates of patients with cancer (PCT/GB2020/050221) and on a related U.S. patent (US12416051B2). Under institutional policies, D.B. may be entitled to a share of revenues arising from licensing of this intellectual property. AMS serves as an adviser to Forcefield Therapeutics. All other authors have nothing to disclose.

## Acknowledgements

This research was conducted using the UK Biobank Resource under application number 98729. We acknowledge the participants of UK Biobank and the Estonian Biobank for their contributions. The Estonian Biobank Research Team was responsible for data collection, genotyping, quality control and imputation and consisted of Andres Metspalu, Mait Metspalu, Lili Milani, Reedik Mägi, Mari Nelis and Georgi Hudjashov. The Estonian Genome Center data analyses were carried out in the High Performance Computing Center, University of Tartu.

## Methods

### 1. Study design and population

This study was conducted within the UK Biobank (UKB), a large-scale prospective cohort study of 500,000 individuals aged between 40-69 years at recruitment,^33^ with an average follow-up in the current work of ca. 12 years across endpoints. All participants underwent comprehensive baseline characterisation via self-answered questionnaires, verbal interviews, a series of physical measurements, and extensive biological sampling. In addition, enrolment comprised linkage to electronic health records (EHR) (Hospital Episode Statistics in England, Patient Episode Database for Wales, and Scottish Morbidity Record) and death registers (NHS England and NHS Central Register) with continuous follow-up and ongoing, regular annual new data releases.

External validation was conducted within the Estonian Biobank (EstBB) cohort, a population-based cohort study which has recruited over 200,000 adult participants in two phases, representing approximately 20% of the Estonian adult population.^19^ We utilised first-phase participants recruited between 2002 and 2015, mainly through general practitioners (GPs). At baseline, participants completed detailed questionnaires covering lifestyle, health history, and environmental exposures, accompanied by physical measurements and blood sample collection. Participant records are continuously updated via linkage to several national EHR and death registries.

### 2. Endpoint definition

24 endpoints were chosen across a multi-disease spectrum in line with previous work^15,16,18^, incorporating multiple cardiovascular, cancer and dementia diagnoses and several common diseases with existing, actionable preventative measures. Incident disease status was determined from hospital inpatient records (including OPCS and ICD codes), death and cancer registries, and verbal interviews during repeated study visits. Endpoint incidence was defined as earliest occurrence from either source. Endpoint definitions were adapted as previously defined^15,18,34,35^. Supplementary Table 1 provides comprehensive phenotype definitions including the utilised data fields and codes.

### 3. Predictor modalities and risk models

#### 3.1 Benchmark clinical risk score

Our clinical benchmark was previously described and (in addition to age and sex, which were included in all predictor matrices; Supplementary Table 7) consists of multiple predictors across a broad spectrum of physical measurements, clinical chemistry, family and past medical history, and medications (Supplementary Table 8).^15^ Although likely to be too extensive and complex to deploy at population scale, it serves as a strict benchmark with predictive value across a multi-disease spectrum.

#### 3.2 Omics assays

##### Polygenic risk scores

Genomics PLC developed a range of PRS on a collection of non-UKB GWAS datasets, which were subsequently derived for all UKB participants. Scores were ancestry-centred and corrected for the first ten genetic principal components.^36^ These are available to UKB researchers as “Standard PRS” (data category ID 301), which we included for all endpoints under investigation and used as a uniform, multi-PRS predictor matrix as model training input across endpoints (see Supplementary Table 9).

##### ^1^H-NMR metabolomics

At the time of conducting this study, UKB provides ^1^H-NMR-based measurements of 168 circulating metabolites for ca. 280,000 individuals. Non-fasting, venous EDTA plasma samples were analysed for UKB by Nightingale Health Inc, using a high-throughput platform with regulatory approval. We utilised all original metabolite measurements (168 metabolites, with a strong focus on lipids and lipid-subspecies; Supplementary Table 10).

#### 3.3 PMH and self-answered questionnaires

##### Past medical history

PMH was extracted via utilisation of inpatient electronic health records (datafield ID 41270) before enrolment and simplified to a Boolean event indicator across ICD subcategories (e.g. “I26: Pulmonary embolism”, containing “I26.0: Pulmonary embolism with mention of acute cor pulmonale” and “I26.9: Pulmonary embolism without mention of acute cor pulmonale”). We did not incorporate the time of diagnosis. We excluded at a minority-class threshold of 0.1 % (final predictors detailed in Supplementary Table 11).

##### Self-answered questionnaires

UKB participants self-answered standardised questionnaires on a touchscreen device during their initial assessment centre visit. These span multiple sub-categories (“Sociodemographics”, “Lifestyle and environment”, “Early life factors”, “Family history”, “Psychosocial factors”, “Health and medical history” and “Sex-specific factors”), each covered with multiple questions (8-155 questions per sub-category; UKB category ID 100025). Factor variables were one-hot encoded (mixed-type variables to tertiles of the continuous variable and the individual factors). We excluded at a minority-class threshold of 0.1 % (final predictors detailed in Supplementary Table 12).

#### 3.4 Cardiovascular risk scores

We extracted the predictors of three nationally-employed cardiovascular risk scores according to U.K. (QRISK3^10^), European (SCORE2^9^), and U.S. (PREVENT^11^) guidelines. An overview of the predictor variables is shown in Figure 3. Detailed information regarding extraction, including the respective data fields and codes, is reported in Supplementary Tables 13-15.

#### 3.5 NHS Health Check

The NHS Health Check (NHS-HC) is an in-person preventative health screening programme targeting the healthy, middle-aged U.K. population. It aims to detect cardiovascular risk factors via a relatively simple combination of physical measurements (weight and blood pressure), clinical chemistry (including cholesterol, glucose or HbA1C) and lifestyle factors (smoking, physical activity and alcohol consumption). Overall, the NHS-HC is effective^37^, but uptake is relatively low and varies according to sociodemographic background^8^. Moreover, although standardised, practitioners regularly skip important parts of the check (particularly assessing diabetic risk, physical activity and alcohol consumption)^23^. We benchmarked against a complete NHS-HC and therefore included all recommended predictors. A detailed feature list including UKB field IDs is shown in Supplementary Table 16.

**Table 2:** Overview of the modelled risk scoring strategies: In this work, we test a variety of scenarios, ranging from clinical benchmarks to integrated multi-omic models. The table comprehensively lists the predictors that the respective models were trained on.

| Model | Included Predictors |
| --- | --- |
| Clinical Model | Clinical Risk Score (3.1 Benchmark Clinical Risk Score) |
| Clinical Model +<br>PMH/Questionnaires | Clinical Risk Score (3.1 Benchmark Clinical Risk Score)<br>Past medical history (3.3 PMH and self-answered<br>questionnaires)<br>Self-answered questionnaires (3.3 PMH and self-answered<br>questionnaires) |
| Clinical Model +<br>PMH/Questionnaires + Omics | Clinical Risk Score (3.1 Benchmark Clinical Risk Score)<br>Past medical history (3.3 PMH and self-answered<br>questionnaires)<br>Self-answered questionnaires (3.3 PMH and self-answered<br>questionnaires)<br>1H-NMR Metabolomics (3.2 Omics assays)<br>Polygenic risk scores (3.2 Omics assays) |
| No Needle Model | Past medical history (3.3 PMH and self-answered<br>questionnaires)<br>Self-answered questionnaires (3.3 PMH and self-answered<br>questionnaires) |
| Single Blood Draw Model | Past medical history (3.3 PMH and self-answered<br>questionnaires)<br>Self-answered questionnaires (3.3 PMH and self-answered<br>questionnaires)<br>1H-NMR Metabolomics (3.2 Omics assays)<br>Polygenic risk scores (3.2 Omics assays) |
| QRISK3 | QRISK3 variables (3.5 Cardiovascular risk scores) |
| SCORE2 | SCORE2 variables (3.5 Cardiovascular risk scores) |
| PREVENT | PREVENT variables (3.5 Cardiovascular risk scores) |
| 1-Step NHS Health Check | NHS Health Check variables (3.5 NHS Health Check) |
| 1-Step NHS Health Check +<br>PMH/Questionnaires | NHS Health Check variables (3.5 NHS Health Check)<br>Past medical history (3.3 PMH and self-answered<br>questionnaires)<br>Self-answered questionnaires (3.3 PMH and self-answered<br>questionnaires) |
| 2-Step NHS Health Check (No<br>Needle Model-driven Invitation of<br>> 5% Risk) | NHS Health Check variables (3.5 NHS Health Check)<br>Past medical history (3.3 PMH and self-answered<br>questionnaires)<br>Self-answered questionnaires (3.3 PMH and self-answered<br>questionnaires) |
| 2-Step NHS Health Check (No<br>Needle Model-driven Invitation of<br>> 5% Risk + Omics for attendees) | NHS Health Check variables (3.5 NHS Health Check)<br>Past medical history (3.3 PMH and self-answered<br>questionnaires)<br>Self-answered questionnaires (3.3 PMH and self-answered<br>questionnaires)<br>1H-NMR Metabolomics (3.2 Omics assays; for ca. 45% subset<br>of the population)<br>Polygenic risk scores (3.2 Omics assays, for ca. 45% subset of<br>the population) |

### 4. Cohort splitting, Preprocessing and Imputation

The dataset was randomly split into 50% training and test partitions. Table 1 reports details on the baseline characteristics of the two populations.

Variables with > 20% missingness were excluded pre-imputation. Imputation was performed via an imputation with chained equations algorithm with a random forest predictor, utilising the *miceRanger* R package (version 1.5.0). The imputation model was trained on the training partition and subsequently applied to the full dataset without retraining to avoid data leakage.

Post-imputation processing included replacing zeroes with 1/10^th^ of the median for all continuous variables (assuming detection thresholds and technical artefacts), alongside a logarithmic transformation and standard scaling (utilising the training subset’s mean and standard deviation (SD)). For blood measurements, we conducted outlier removal as described below (“Inclusion and Exclusion criteria”).

### 5. Inclusion and exclusion criteria

We included all UKB participants with available baseline ^1^H-NMR measurements and PRS (*n* = 271,669). Subsequent exclusion steps included: (I) missingness-based exclusion: we excluded participants with > 20 % missingness within any predictor matrix; (II) exclusion of individuals with outlier blood (clinical chemistry and metabolomics) measurements > 5 SDs from the mean; (III) For the respective models (and evaluations), participants with a baseline diagnosis by the criteria outlined above (Methods: 2. Endpoint definitions; Supplementary Table 1), were excluded (final cohort size *n* = 239,207).

### 6. Model Fitting and Evaluation

Elastic net (EN)-penalized Cox proportional hazards (Cox-PH) models were trained for each unique predictor and endpoint combination, utilising training partition data points only. To allow a fair comparison, this included cardiovascular risk scores (QRISK3, SCORE2 and PREVENT), for which we refitted models using the respective predictor matrices as defined by the score (Figure 3, Supplementary Tables 13-15).

This was implemented via scikit-survival’s CoxnetSurvivalAnalysis(): In short, we fit an initial model at a fixed L1/L2 regularization ratio of 0.5 (l1_ratio: 0.5), the model automatically determines alpha_max_ (the smallest alpha leading all model coefficients to become zero) and subsequently chooses alpha values along a definable regularization path (alpha_min_ratio: 0.01; thus in our case ranging from 0.01 x alpha_max_ to alpha_max_; we restricted this to five alpha values). Hyperparameter optimisation for Harrell’s C was conducted using an internal-to-training 5-fold cross-validation approach, testing the obtained alpha grid and ten l1_ratios linearly spaced between 0.1 and 1. A final model was fitted on the full training partition using the optimised hyperparameters.

The trained models were subsequently applied to obtain risk predictions for the unseen test partition. Subsequent bootstrapping across 100 iterations allowed us to obtain confidence intervals (Supplementary Tables 2-5 for comprehensive results). Individual feature contributions to the respective risk scores were obtained by summing the absolute coefficients.

### 7. Modelling screening scenarios within the NHS Health Check

The NHS Health Check aims to predict 10-year risk for heart disease and stroke (summarized as CVD in our analysis), diabetes mellitus (DM) and kidney disease (KD).^38^ We therefore restricted our analysis to this time span. This was achieved via artificial right-censoring at 10 years (recoding observations past 10 years of follow-up, and subsequent clipping of follow-up times at 10 years). The NHS Health Check also only assesses healthy individuals without a previous diagnosis of a range of conditions (e.g. diagnosed hypertension) or readily implemented risk factor control (e.g. individuals already receiving statins)^38^. We accordingly excluded all individuals with such diagnoses or pharmacotherapy regimens. This is relevant because it removed large numbers of participants with obvious high-risk constellations and instead restricted subsequent analyses to relatively healthy individuals, who might otherwise potentially be overlooked; NHS Health Check exclusion criteria were present in ca. 25% of individuals.

Finally, we fitted EN-regularized Cox-PH models as outlined above. For two-step approaches (schematic overview in Figure 5A), we built two models. A first model (“Step 1”) was trained to obtain 10-year risk predictions; these models were obtained by utilising remotely collectable data (self-answered questionnaires and PMH). A second model (“Step 2”) was trained on high-predicted-risk individuals using the “Step 1” predictions (> 5% risk for any of the endpoints; 36% of the population). “Step 2” models used additional predictors that would be obtained as part of an in-person NHS Health Check, in one scenario with the additional inclusion of genomic and metabolomic profiling. The obtained models were applied to the test partition in the same order: (I) “Step 1” predictions, which were retained as final risk estimates for individuals with < 5% risk across the three endpoints, followed by (II) “Step 2” predictions for individuals with > 5% “Step 1” risk for any of the endpoints, which were retained as final risk estimates where calculated.

### 8. External Validation in Estonian Biobank

Due to the large number of investigated predictors, there is no single cohort allowing for comprehensive external validation of our results. However, we were able to externally validate a key piece of our work – the predictive accuracy of an in-principle remote screening scenario (i.e. the “No Needle Model”) against cardiovascular risk score benchmarks -in EstBB^19^. The “Clinical Model” benchmark was not replicable due to several missing predictors, and weights for the utilised PRS (by Genomics plc) are not publicly available.

In the first instance, we carefully matched the self-reported information following the information outlined in Supplementary Table 17 (including information on mapping and transformation steps). We chose slightly more permissive missingness thresholds (25%), which allowed us to replicate 195 of 1252 (ca. 16 %) questionnaire features. For the cardiovascular risk score benchmarks, some single features were unavailable (two optional PREVENT features (uACR and HbA1c), the utilised deprivation index (used by PREVENT and QRISK3), family history of heart disease and the standard deviation of the two most recent SBP readings (both used by QRISK3)). We then followed the exact same preprocessing routine as for the UKB dataset (including imputation and scaling).

Within the UKB training set, we refitted our models on the predictor subset available within EstBB, and – using the obtained coefficients – constructed linear predictors within EstBB. Finally, again following the identical steps as in our UKB test set, we were able to obtain performance and bootstrapping-derived confidence estimates in EstBB.

### 9. Significance, software and code availability

Models were compared using false discovery rate (FDR; following the Benjamini-Hochberg procedure)-adjusted, paired t-tests. Significance was defined as FDR-adjusted p < 0.05. Analyses were conducted on UKB’s Research Analysis Platform (hosted by DNAnexus®, powered by Amazon Web Services). Different parts of the workflow were performed in R 4.4.0 and Python 3.9.16. Large language models were used to assist with coding steps and proof-reading. All code will be made publicly available via GitHub upon publication and can be shared with reviewers upon request.

### 10. Data availability

The UKB dataset is publicly available to approved researchers at https://www.ukbiobank.ac.uk/enable-your-research. We provide definitions for endpoints and predictor matrices as well as detailed performance metrics in our supplementary material.

## Notes

### Author Declarations

North West - Haydock Research Ethics Committee gave ethical approval for this work. This ethical approval letter can be accessed via: https://www.ukbiobank.ac.uk/wp-content/uploads/2025/01/Ethics-approval-renewal-2021.pdf

