## Supplementary Material for "Multi-disease risk prediction models for population-scale personalised screening"

**Supplementary Tables**

**Supplementary Table 1.** Endpoint Definitions

**Supplementary Table 2.** Absolute Performance of Modelled Scenarios

**Supplementary Table 3.** Absolute Performance for CVD Risk Stratification Models

**Supplementary Table 4.** Absolute Performance for NHS-HC Scenarios

**Supplementary Table 5.** Absolute Performance of Benchmarked Predictor Matrices

**Supplementary Table 6.** External Validation: Absolute Performance in EstBB

**Supplementary Table 7.** Predictors: Age & Sex

**Supplementary Table 8.** Predictors: Clinical Risk

**Supplementary Table 9.** Predictors: Polygenic Risk Scores (PRS)

**Supplementary Table 10.** Predictors: ¹H-NMR Metabolomics

**Supplementary Table 11.** Predictors: Past Medical History (PMH)

**Supplementary Table 12.** Predictors: Self-Reported Information (separate Excel file)

**Supplementary Table 13.** Predictors: SCORE-2

**Supplementary Table 14.** Predictors: PREVENT

**Supplementary Table 15.** Predictors: QRISK3

**Supplementary Table 16.** Predictors: NHS-HC

**Supplementary Table 17.** External Validation: Predictor Mapping in EstBB (separate Excel file)

**Supplementary Table 18.** External Validation: Endpoint Frequency in EstBB

**Supplementary Figures**

**Supplementary Figure 1.** External validation of “No Needle Model”-based risk prediction in the Estonian Biobank.

**Supplementary Tables**

| **Field names** | **Field ID** | **Coding** | **Meaning** |
| --- | --- | --- | --- |
| ***Abdominal aortic aneurysm*** |  |  |  |
| Diagnoses - ICD10  Underlying (primary) cause of death: ICD10  Contributory (secondary) causes of death: ICD10 | 41270  40001  40002 | I71, I710, I711, I712, I713, I714, I715, I716, I718, I719 | Aortic aneurysm and dissection |
| Diagnoses - ICD9 | 41271 | 4410, 4411, 4412, 4413, 4414, 4415, 4416 | Dissecting aneurysm (any part), Thoracic aneurysm, ruptured, Thoracic aneurysm without mention of rupture, Abdominal aneurysm, ruptured, Abdominal aneurysm without mention of rupture, Aortic aneurysm of unspecified site, ruptured, Aortic aneurysm of unspecified site without mention of rupture |
| ***Alzheimer’s disease*** |  |  |  |
| Diagnoses - ICD10  Underlying (primary) cause of death: ICD10  Contributory (secondary) causes of death: ICD10 | 41270  40001  40002 | F00, F000, F001, F002, F009;  G30, G300, G301, G308, G309 | Dementia in Alzheimer's disease;  Alzheimer’s disease |
| Diagnoses - ICD9 | 41271 | 3310 | Alzheimer’s disease |
| Non-cancer illness code, self-reported | 20002 | 1263 | dementia/alzheimers/cognitive impairment |
| ***Atrial fibrillation*** |  |  |  |
| Diagnoses - ICD10  Underlying (primary) cause of death: ICD10  Contributory (secondary) causes of death: ICD10 | 41270  40001  40002 | I48, I480, I481, I482, I483, I484, I489 | Atrial fibrillation and flutter |
| Diagnoses - ICD9 | 41271 | 4273 | Atrial fibrillation and flutter |
| Operative procedures - OPCS4 | 41272 | K571;  K621, K622, K623, K624 | Percutaneous transluminal ablation of atrioventricular node;  Percutaneous transluminal ablation of pulmonary vein to left atrium conducting system,  Percutaneous transluminal ablation of atrial wall for atrial flutter,  Percutaneous transluminal ablation of conducting system of heart for atrial flutter NEC,  Percutaneous transluminal internal cardioversion NEC |
| Non-cancer illness code, self-reported | 20002 | 1471 | atrial fibrillation |
| Operation code | 20004 | 1524 | cardioversion |
| ***Asthma*** |  |  |  |
| Diagnoses - ICD10  Underlying (primary) cause of death: ICD10  Contributory (secondary) causes of death: ICD10 | 41270  40001  40002 | J45, J450, J451, J458, J459;  J46 | Asthma;  Status asthmaticus |
| Diagnoses - ICD9 | 41271 | 493, 4930, 49300, 49309, 4931, 49310, 49319, 4939, 49390, 49399 | Asthma |
| Non-cancer illness code, self-reported | 20002 | 1111 | asthma |
| ***Breast cancer*** |  |  |  |
| Diagnoses - ICD10  Underlying (primary) cause of death: ICD10  Contributory (secondary) causes of death: ICD10 | 41270  40001  40002 | C50, C500, C501, C502, C503, C504, C505, C506, C508, C509 | Malignant neoplasm of breast |
| Diagnoses - ICD9 | 41271 | 1740, 1741, 1742, 1743, 1744, 1745, 1746, 1748, 1749;  175 | Malignant neoplasm of female breast; Malignant neoplasm of male breast |
| Cancer code, self-reported | 20001 | 1002 | breast cancer |
| ***Cataracts*** |  |  |  |
| Diagnoses - ICD10  Underlying (primary) cause of death: ICD10  Contributory (secondary) causes of death: ICD10 | 41270  40001  40002 | H25, H250, H251, H252, H258, H259;  H26, H260, H261, H262, H263, H264, H268, H269 | Senile cataract; Other cataract |
| Diagnoses - ICD9 | 41271 | 3660, 3662, 3663, 3664, 3665, 3668 | Infant, juvenile and presenile cataract, Traumatic cataract, Cataract secondary to ocular disorders, Cataract associated with other disorders, After-cataract, Other specified cataract |
| ***Coronary artery disease*** |  |  |  |
| Diagnoses - ICD10  Underlying (primary) cause of death: ICD10  Contributory (secondary) causes of death: ICD10 | 41270  40001  40002 | I21, I210, I211, I212, I213, I214, I219;  I22, I220, I221, I228, I229;  I23, I231, I232, I233, I236, I238;  I241;  I252 | Acute myocardial infarction;  Subsequent myocardial infarction;  Certain current complications following acute myocardial infarction;  Acute ischaemic heart disease;  Old myocardial infarction |
| Operative procedures - OPCS4 | 41272 | K401, K402, K403, K404;  K411, K412, K413, K414;  K451, K452, K453, K454, K455;  K491, K492, K498, K499;  K502;  K751, K752, K753, K754, K758, K759 | Saphenous vein graft replacement of coronary arteries;  Autograft replacement of coronary arteries NEC;  Mammary/thoracic artery to coronary artery anastomoses;  Percutaneous transluminal balloon angioplasty of coronary arteries;  Percutaneous transluminal coronary thrombolysis using streptokinase;  PTCA and insertion of drug-eluting or other stents into coronary artery |
| Non-cancer illness code, self-reported | 20002 | 1075 | heart attack/myocardial infarction |
| Operation code | 20004 | 1070, 1095 | coronary angioplasty (ptca) +/- stent, coronary artery bypass grafts (cabg) |
| ***Chronic obstructive pulmonary disease*** | | |  |
| Diagnoses - ICD10  Underlying (primary) cause of death: ICD10  Contributory (secondary) causes of death: ICD10 | 41270  40001  40002 | J40;  J41, J410, J411, J418;  J42;  J43, J430, J431, J432, J438, J439;  J44, J440, J441, J448, J449;  J47 | Bronchitis not specified as acute or chronic;  Simple and mucopurulent chronic bronchitis;  Unspecified chronic bronchitis;  Emphysema;  Other chronic obstructive pulmonary disease;  Bronchiectasis |
| Diagnoses - ICD9 | 41271 | 490;  4910, 4911, 4919;  492, 4929;  494, 4949 | Bronchitis not specified as acute or chronic;  Chronic bronchitis;  Emphysema;  Bronchiectasis |
| Non-cancer illness code, self-reported | 20002 | 1112, 1113, 1472 | chronic obstructive airways disease/copd, emphysema/chronic bronchitis, emphysema |
| ***Colorectal cancer*** |  |  |  |
| Diagnoses - ICD10  Underlying (primary) cause of death: ICD10  Contributory (secondary) causes of death: ICD10 | 41270  40001  40002 | C18, C180, C181, C182, C183, C184, C185, C186, C187, C188, C189;  C19;  C20 | Malignant neoplasm of colon;  Malignant neoplasm of rectosigmoid junction;  Malignant neoplasm of rectum |
| Diagnoses - ICD9 | 41271 | 1530, 1531, 1532, 1533, 1534, 1535, 1536, 1537, 1538, 1539;  1540, 1541 | Malignant neoplasm of colon;  Malignant neoplasm of rectosigmoid junction, Malignant neoplasm of rectum |
| Cancer code, self-reported | 20001 | 1019, 1020, 1022, 1023 | small intestine/small bowel cancer, large bowel cancer/colorectal cancer, colon cancer/sigmoid cancer, rectal cancer |
| ***Cardiovascular disease*** |  |  |  |
| Diagnoses - ICD10  Underlying (primary) cause of death: ICD10  Contributory (secondary) causes of death: ICD10 | 41270  40001  40002 | G45, G450, G451, G452, G453, G454, G458, G459;  I20, I200, I201, I208, I209;  I21, I210, I211, I212, I213, I214, I219;  I22, I220, I221, I228, I229;  I23, I231, I232, I233, I236, I238;  I24, I240, I241, I248, I249;  I25, I250, I251, I252, I253, I254, I255, I256, I258, I259;  I63, I630, I631, I632, I633, I634, I635, I636, I638, I639;  I64 | Transient cerebral ischaemic attacks and related syndromes;  Angina pectoris;  Acute myocardial infarction;  Subsequent myocardial infarction;  Certain current complications following acute myocardial infarction;  Other acute ischaemic heart diseases;  Chronic ischaemic heart disease;  Cerebral infarction;  Stroke, not specified as haemorrhage or infarction; |
| Diagnoses - ICD9 | 41271 | 410, 4109;  411, 4119;  412, 4129;  413, 4139;  414, 4140, 4141, 4148, 4149;  42979;  434, 4340, 4341, 4349;  436, 4369 | Acute myocardial infarction;  Other acute and subacute forms of ischaemic heart disease;  Old myocardial infarction;  Angina pectoris;  Other forms of chronic ischaemic heart disease;  Ill-defined descriptions and complications of heart disease; Occlusion of cerebral arteries;  Acute but ill-defined cerebrovascular disease |
| Operative procedures - OPCS4 | 41272 | K40, K401, K402, K403, K404, K408, K409;  K41, K411, K412, K413, K414, K418, K419;  K42, K421, K422, K423, K424, K428, K429;  K43, K431, K432, K433, K434, K438, K439;  K44, K441, K442, K448, K449;  K45, K451, K452, K453, K454, K455, K456, K458, K459;  K46, K461, K462, K463, K464, K465, K468, K469;  K471;  K49, K491, K492, K493, K494, K498, K499;  K50, K501, K502, K503, K504, K508, K509;  K75, K751, K752, K753, K754, K758, K759 | Saphenous vein graft replacement of coronary artery;  Other autograft replacement of coronary artery;  Allograft replacement of coronary artery;  Prosthetic replacement of coronary artery;  Other replacement of coronary artery;  Connection of thoracic artery to coronary artery;  Other bypass of coronary artery;  Endarterectomy of coronary artery;  Transluminal balloon angioplasty of coronary artery;  Other therapeutic transluminal operations on coronary artery;  Percutaneous transluminal balloon angioplasty and insertion of stent into coronary artery |
| Non-cancer illness code, self-reported | 20002 | 1074, 1075, 1082, 1583 | angina, heart attack/myocardial infarction, transient ischaemic attack (tia), ischaemic stroke |
| Operation code | 20004 | 1070, 1071, 1095, 1105, 1109, 1514 | coronary angioplasty (ptca) +/- stent, other arterial surgery/revascularisation procedures, coronary artery bypass grafts (cabg), carotid artery surgery/endarterectomy, carotid artery angioplasty +/- stent, coronary angiogram |
| ***Diabetes Mellitus*** |  |  |  |
| Diagnoses - ICD10  Underlying (primary) cause of death: ICD10  Contributory (secondary) causes of death: ICD10 | 41270  40001  40002 | E10, E100, E101, E102, E103, E104, E105, E106, E107, E108, E109;  E11, E110, E111, E112, E113, E114, E115, E116, E117, E118, E119;  E12, E120, E121, E122, E123, E124, E125, E126, E127, E128, E129;  E13, E130, E131, E132, E133, E134, E135, E136, E137, E138, E139;  E14, E140, E141, E142, E143, E144, E145, E146, E147, E148, E149 | Insulin-dependent diabetes mellitus;  Non-insulin-dependent diabetes mellitus;  Malnutrition-related diabetes mellitus;  Other specified diabetes mellitus;  Unspecified diabetes mellitus |
| Diagnoses - ICD9 | 41271 | 250, 2500, 25000, 25001, 25009, 2501, 25010, 25011, 25019, 2502, 25020, 25021, 25029, 2503, 2504, 2505, 2506, 2507, 2509, 25090, 25091, 25099 | Diabetes mellitus |
| Non-cancer illness code, self-reported | 20002 | 1220, 1222, 1223 | diabetes, type 1 diabetes, type 2 diabetes |
| ***Heart failure*** |  |  |  |
| Diagnoses - ICD10  Underlying (primary) cause of death: ICD10  Contributory (secondary) causes of death: ICD10 | 41270  40001  40002 | I110;  I130, I132;  I255;  I420, I425, I428, I429;  I500, I501, I509 | Hypertensive heart disease with heart failure;  Hypertensive heart and renal disease with heart failure;  Atherosclerotic heart disease;  Cardiomyopathy;  Heart failure |
| Diagnoses - ICD9 | 41271 | 4254;  4280, 4281, 4289 | Other primary cardiomyopathies;  Congestive heart failure, Left heart failure, Heart failure, unspecified |
| Non-cancer illness code, self-reported | 20002 | 1076, 1079 | heart failure/pulmonary odema, cardiomyopathy |
| ***Hypertension*** |  |  |  |
| Diagnoses - ICD10  Underlying (primary) cause of death: ICD10  Contributory (secondary) causes of death: ICD10 | 41270  40001  40002 | I10; I15, I150, I151, I152, I159 | Essential (primary) hypertension; Secondary hypertension |
| Diagnoses - ICD9 | 41271 | 4010, 4011, 4019; 4050, 4051, 4059 | Essential hypertension; Secondary hypertension |
| Non-cancer illness code, self-reported | 20002 | 1065, 1072 | hypertension, high blood pressure |
| ***Ischaemic stroke*** |  |  |  |
| Diagnoses - ICD10  Underlying (primary) cause of death: ICD10  Contributory (secondary) causes of death: ICD10 | 41270  40001  40002 | I63, I630, I631, I632, I633, I634, I635, I636, I638, I639;  I64 | Cerebral infarction;  Stroke, not specified as haemorrhage or infarction |
| Diagnoses - ICD9 | 41271 | 436 | Acute but ill-defined cerebrovascular disease |
| ***Lung cancer*** |  |  |  |
| Diagnoses - ICD10  Underlying (primary) cause of death: ICD10  Contributory (secondary) causes of death: ICD10 | 41270  40001  40002 | C33;  C34, C340, C341, C342, C343, C348, C349 | Malignant neoplasm of trachea;  Malignant neoplasm of bronchus and lung |
| Diagnoses - ICD9 | 41271 | 1620, 1622, 1623, 1624, 1625, 1628, 1629 | Malignant neoplasm of trachea, bronchus and lung |
| ***Liver disease*** |  |  |  |
| Diagnoses - ICD10  Underlying (primary) cause of death: ICD10  Contributory (secondary) causes of death: ICD10 | 41270  40001  40002 | B15, B150, B159;  B16, B160, B161, B162, B169;  B17, B170, B171, B172, B178, B179;  B18, B180, B1800, B1809, B181, B1810, B1819, B182, B188, B189;  B19, B190, B199;  C22, C220, C221, C222, C223, C224, C227, C229;  E83, E830, E831, E832, E833, E834, E835, E838, E839;  E88, E880, E881, E882, E883, E888, E889;  I85, I850, I859;  K70, K700, K701, K702, K703, K704, K709;  K72, K720, K721, K729;  K73, K730, K731, K732, K738, K739;  K74, K740, K741, K742, K743, K744, K745, K746;  K75, K750, K751, K752, K753, K754, K758, K759;  K76, K760, K761, K762, K763, K764, K765, K766, K767, K768, K769;  R18;  Z94, Z940, Z941, Z942, Z943, Z944, Z945, Z946, Z947, Z948, Z949 | Acute hepatitis A;  Acute hepatitis B;  Other acute viral hepatitis;  Chronic viral hepatitis;  Unspecified viral hepatitis;  Malignant neoplasm of liver and intrahepatic bile ducts;  Disorders of mineral metabolism;  Other metabolic disorders;  Oesophageal varices;  Alcoholic liver disease;  Hepatic failure, not elsewhere classified;  Chronic hepatitis, not elsewhere classified;  Fibrosis and cirrhosis of liver;  Other inflammatory liver diseases;  Other diseases of liver;  Ascites;  Transplanted organ and tissue status |
| Diagnoses - ICD9 | 41271 | 0700, 0701, 0706, 0709;  1550, 1551, 1552;  2750, 2751, 2752, 2753, 2759;  4560, 4561;  5710, 5711, 5713, 5714, 5716;  5720, 5721, 5723, 5724;  5734, 5739 | Viral hepatitis a with hepatic coma, Viral hepatitis a without mention of hepatic coma, Unspecified viral hepatitis with hepatic coma, Unspecified viral hepatitis without mention of hepatic coma;  Malignant neoplasm of liver and intrahepatic bile ducts;  Disorders of iron metabolism, Disorders of copper metabolism, Disorders of magnesium metabolism, Disorders of phosphorus metabolism, Unspecified disorders of mineral metabolism;  Oesophageal varices with bleeding, Oesophageal varices without mention of bleeding;  Alcoholic fatty liver, Acute alcoholic hepatitis, Alcoholic liver damage, unspecified, Chronic hepatitis, Biliary cirrhosis;  Abscess of liver, Portal pyaemia, Portal hypertension, Hepatorenal syndrome;  Hepatic infarction, Disorders of liver, unspecified |
| ***Melanoma*** |  |  |  |
| Diagnoses - ICD10  Underlying (primary) cause of death: ICD10  Contributory (secondary) causes of death: ICD10 | 41270  40001  40002 | C43, C430, C431, C432, C433, C434, C435, C436, C437, C438, C439 | Malignant melanoma |
| Diagnoses - ICD9 | 41271 | 1720, 1721, 1722, 1723, 1724, 1725, 1726, 1727, 1729 | Malignant melanoma |
| Cancer code, self-reported | 20001 | 1059 | malignant melanoma |
| ***Osteoporosis*** |  |  |  |
| Diagnoses - ICD10  Underlying (primary) cause of death: ICD10  Contributory (secondary) causes of death: ICD10 | 41270  40001  40002 | M80, M800, M8000, M8001, M8002, M8003, M8004, M8005, M8006, M8007, M8008, M8009, M801, M8010, M8011, M8012, M8013, M8014, M8015, M8016, M8017, M8018, M8019, M802, M8020, M8021, M8022, M8023, M8024, M8025, M8026, M8027, M8028, M8029, M803, M8030, M8031, M8032, M8033, M8034, M8035, M8036, M8037, M8038, M8039, M804, M8040, M8041, M8042, M8043, M8044, M8045, M8046, M8047, M8048, M8049, M805, M8050, M8051, M8052, M8053, M8054, M8055, M8056, M8057, M8058, M8059, M808, M8080, M8081, M8082, M8083, M8084, M8085, M8086, M8087, M8088, M8089, M809, M8090, M8091, M8092, M8093, M8094, M8095, M8096, M8097, M8098, M8099;  M81, M810, M8100, M8101, M8102, M8103, M8104, M8105, M8106, M8107, M8108, M8109, M811, M8110, M8111, M8112, M8113, M8114, M8115, M8116, M8117, M8118, M8119, M812, M8120, M8121, M8122, M8123, M8124, M8125, M8126, M8127, M8128, M8129, M813, M8130, M8131, M8132, M8133, M8134, M8135, M8136, M8137, M8138, M8139, M814, M8140, M8141, M8142, M8143, M8144, M8145, M8146, M8147, M8148, M8149, M815, M8150, M8151, M8152, M8153, M8154, M8155, M8156, M8157, M8158, M8159, M816, M8160, M8161, M8162, M8163, M8164, M8165, M8166, M8167, M8168, M8169, M818, M8180, M8181, M8182, M8183, M8184, M8185, M8186, M8187, M8188, M8189, M819, M8190, M8191, M8192, M8193, M8194, M8195, M8196, M8197, M8198, M8199;  M82, M820, M8200, M8201, M8202, M8203, M8204, M8205, M8206, M8207, M8208, M8209, M821, M8210, M8211, M8212, M8213, M8214, M8215, M8216, M8217, M8218, M8219, M823, M828, M8280, M8281, M8282, M8283, M8284, M8285, M8286, M8287, M8288, M8289 | Osteoporosis with pathological fracture;  Osteoporosis without pathological fracture;  Osteoporosis in diseases classified elsewhere; |
| Non-cancer illness code, self-reported | 20002 | 1309 | Osteoporosis |
| ***Peripheral artery disease*** |  |  |  |
| Diagnoses - ICD10  Underlying (primary) cause of death: ICD10  Contributory (secondary) causes of death: ICD10 | 41270  40001  40002 | I70, I700, I7000, I7001, I701, I7010, I7011, I702, I7020, I7021, I708, I7080, I7081, I709, I7090, I7091;  I71, I710, I711, I712, I713, I714, I715, I716, I718, I719;  I72, I720, I721, I722, I723, I724, I725, I726, I728, I729;  I73, I730, I731, I738, I739;  I74, I740, I741, I742, I743, I744, I745, I748, I749;  I77, I770, I771, I772, I773, I774, I775, I776, I778, I779;  I78, I780, I781, I788, I789;  I79, I790, I791, I792, I798 | Atherosclerosis;  Aortic aneurysm and dissection;  Other aneurysms;  Other peripheral vascular diseases;  Arterial embolism and thrombosis;  Other disorders of arteries and arterioles;  Diseases of capillaries |
| Diagnoses - ICD9 | 41271 | 4400, 4401, 4402, 4408, 4409;  4410, 4411, 4412, 4413, 4414, 4415, 4416;  4430, 4431, 4438, 4439;  4440, 4441, 4442, 4448, 4449;  4470, 4471, 4472, 4473, 4474, 4475;  4480, 4481 | Atherosclerosis;  Aortic aneurysm and dissection;  Other peripheral vascular disease;  Arterial embolism and thrombosis;  Arterial malformations;  Diseases of capillaries |
| ***Prostate cancer*** |  |  |  |
| Diagnoses - ICD10  Underlying (primary) cause of death: ICD10  Contributory (secondary) causes of death: ICD10 | 41270  40001  40002 | C61 | Malignant neoplasm of prostate |
| Diagnoses - ICD9 | 41271 | 185 | Malignant neoplasm of prostate |
| Cancer code, self-reported | 20001 | 1044 | prostate cancer |
| ***Parkinson’s disease*** |  |  |  |
| Diagnoses - ICD10  Underlying (primary) cause of death: ICD10  Contributory (secondary) causes of death: ICD10 | 41270  40001  40002 | G20 | Parkinson's disease |
| Diagnoses - ICD9 | 41271 | 3320 | Parkinson's disease |
| Non-cancer illness code, self-reported | 20002 | 1262 | parkinsons disease |
| ***Primary open angle glaucoma*** | |  |  |
| Diagnoses - ICD10  Underlying (primary) cause of death: ICD10  Contributory (secondary) causes of death: ICD10 | 41270  40001  40002 | H401, H409 | Primary open-angle glaucoma, Open-angle glaucoma, unspecified |
| Non-cancer illness code, self-reported | 20002 | 1277 | glaucoma |
| ***Renal disease*** |  |  |  |
| Diagnoses - ICD10  Underlying (primary) cause of death: ICD10  Contributory (secondary) causes of death: ICD10 | 41270  40001  40002 | N00, N000, N001, N002, N003, N004, N005, N006, N007, N008, N009;  N01, N010, N011, N012, N013, N014, N015, N016, N017, N018, N019;  N02, N020, N021, N022, N023, N024, N025, N026, N027, N028, N029;  N03, N030, N031, N032, N033, N034, N035, N036, N037, N038, N039;  N04, N040, N041, N042, N043, N044, N045, N046, N047, N048, N049;  N05, N050, N051, N052, N053, N054, N055, N056, N057, N058, N059;  N06, N060, N061, N062, N063, N064, N065, N066, N067, N068, N069;  N07, N070, N071, N072, N073, N074, N075, N076, N077, N078, N079;  N08, N080, N081, N082, N083, N084, N085, N088;  N10;  N11, N110, N111, N118, N119;  N12;  N13, N130, N131, N132, N133, N134, N135, N136, N137, N138, N139;  N14, N140, N141, N142, N143, N144;  N15, N150, N151, N158, N159;  N16, N160, N161, N162, N163, N164, N165, N168;  N17, N170, N171, N172, N178, N179;  N18, N180, N181, N182, N183, N184, N185, N188, N189;  N19;  N25, N250, N251, N258, N259;  N26;  N27, N270, N271, N279;  N28, N280, N281, N288, N289;  N29, N290, N291, N298; | Acute nephritic syndrome;  Rapidly progressive nephritic syndrome;  Recurrent and persistent haematuria;  Chronic nephritic syndrome;  Nephrotic syndrome;  Unspecified nephritic syndrome;  Isolated proteinuria with specified morphological lesion;  Hereditary nephropathy, not elsewhere classified;  Glomerular disorders in diseases classified elsewhere;  Acute tubulo-interstitial nephritis;  Chronic tubulo-interstitial nephritis;  Tubulo-interstitial nephritis, not specified as acute or chronic;  Obstructive and reflux uropathy;  Drug- and heavy-metal-induced tubulo-interstitial and tubular conditions;  Other renal tubulo-interstitial diseases;  Renal tubulo-interstitial disorders in diseases classified elsewhere;  Acute renal failure;  Chronic renal failure;  Unspecified renal failure;  Disorders resulting from impaired renal tubular function;  Unspecified contracted kidney;  Small kidney of unknown cause;  Other disorders of kidney and ureter, not elsewhere classified;  Other disorders of kidney and ureter in diseases classified elsewhere |
| Diagnoses - ICD9 | 41271 | 5810, 5812, 5813;  5822;  5845, 5846, 5847, 5848, 5849;  585;  586;  587;  5880, 5888, 5889;  5890, 5891, 5899;  5902, 5909;  5932, 5935 | Nephrotic syndrome with lesion of proliferative glomerulonephritis, Nephrotic syndrome with lesion of membranoproliferative glomerulonephritis, Nephrotic syndrome with lesion of minimal change glomerulonephritis;  Chronic glomerulonephritis with lesion of membranoproliferative glomerulonephritis;  Acute renal failure;  Chronic renal failure;  Renal failure, unspecified;  Renal sclerosis, unspecified;  Renal osteodystrophy, Other specified disorders resulting from impaired renal function, Disorders resulting from impaired renal function, unspecified;  Small kidney of unknown cause;  Renal and perinephric abscess, Infection of kidney, unspecified;  Cyst of kidney, acquired, Hydroureter |
| ***Venous thrombosis*** |  |  |  |
| Diagnoses - ICD10  Underlying (primary) cause of death: ICD10  Contributory (secondary) causes of death: ICD10 | 41270  40001  40002 | I26, I260, I269;  I81;  I82, I820, I821, I822, I823, I828, I829;  O082;  O223;  O871 | Pulmonary embolism;  Portal vein thrombosis;  Other venous embolism and thrombosis;  Obstetric pulmonary embolism;  Deep phlebothrombosis in pregnancy;  Septic pelvic thrombophlebitis |
| Diagnoses - ICD9 | 41271 | 4150, 4151;  452;  4530, 4531, 4532, 4533, 4538, 4539 | Acute cor pulmonale, Pulmonary embolism;  Portal vein thrombosis;  Budd-Chiari syndrome, Thrombophlebitis migrans, Embolism and thrombosis of vena cava, renal vein, other specified veins, unspecified site |
| Non-cancer illness code, self-reported | 20002 | 1068, 1093, 1094 | venous thromboembolic disease, pulmonary embolism +/- dvt, deep venous thrombosis (dvt) |

**Supplementary Table 1: Endpoint definitions:** The table details endpoint definitions, including the UKB field IDs which information was extracted from and the respective disease codes alongside their meaning.

| **Endpoint** | **No Needle Model** | **Single Blood Draw Model** | **Clinical Model** | **Clinical Model + PMH/Questionnaires** | **Clinical Model + PMH/Questionnaires + Omics** |
| --- | --- | --- | --- | --- | --- |
| AAA | 0.822 (0.831 - 0.810) | 0.825 (0.835 - 0.812) | 0.820 (0.830 - 0.807) | 0.831 (0.841 - 0.819) | 0.832 (0.842 - 0.820) |
| AD | 0.850 (0.859 - 0.840) | 0.879 (0.889 - 0.868) | 0.844 (0.852 - 0.832) | 0.858 (0.867 - 0.849) | 0.883 (0.892 - 0.872) |
| AF | 0.768 (0.772 - 0.763) | 0.790 (0.794 - 0.785) | 0.769 (0.774 - 0.764) | 0.779 (0.784 - 0.774) | 0.797 (0.801 - 0.793) |
| AS | 0.727 (0.736 - 0.716) | 0.732 (0.741 - 0.720) | 0.633 (0.641 - 0.621) | 0.729 (0.737 - 0.719) | 0.733 (0.742 - 0.722) |
| BC | 0.561 (0.572 - 0.550) | 0.663 (0.674 - 0.655) | 0.562 (0.572 - 0.552) | 0.573 (0.586 - 0.563) | 0.667 (0.678 - 0.658) |
| CAD | 0.749 (0.757 - 0.743) | 0.783 (0.790 - 0.776) | 0.750 (0.756 - 0.743) | 0.771 (0.778 - 0.764) | 0.789 (0.796 - 0.783) |
| CAT | 0.760 (0.764 - 0.756) | 0.761 (0.764 - 0.756) | 0.750 (0.753 - 0.745) | 0.760 (0.764 - 0.756) | 0.761 (0.764 - 0.757) |
| COPD | 0.859 (0.863 - 0.855) | 0.862 (0.866 - 0.858) | 0.800 (0.805 - 0.794) | 0.863 (0.868 - 0.860) | 0.864 (0.869 - 0.860) |
| CRC | 0.660 (0.671 - 0.649) | 0.702 (0.714 - 0.691) | 0.668 (0.678 - 0.654) | 0.667 (0.677 - 0.654) | 0.705 (0.715 - 0.694) |
| CVD | 0.731 (0.735 - 0.728) | 0.747 (0.750 - 0.743) | 0.720 (0.723 - 0.715) | 0.741 (0.743 - 0.737) | 0.750 (0.753 - 0.745) |
| DM | 0.788 (0.792 - 0.782) | 0.842 (0.846 - 0.837) | 0.866 (0.872 - 0.862) | 0.876 (0.881 - 0.872) | 0.878 (0.882 - 0.874) |
| HF | 0.809 (0.816 - 0.802) | 0.818 (0.824 - 0.811) | 0.800 (0.807 - 0.793) | 0.819 (0.826 - 0.813) | 0.824 (0.830 - 0.817) |
| HT | 0.721 (0.726 - 0.716) | 0.736 (0.740 - 0.732) | 0.762 (0.765 - 0.758) | 0.779 (0.783 - 0.776) | 0.783 (0.787 - 0.780) |
| ISS | 0.745 (0.755 - 0.737) | 0.756 (0.765 - 0.747) | 0.750 (0.761 - 0.740) | 0.757 (0.768 - 0.748) | 0.764 (0.774 - 0.754) |
| LC | 0.833 (0.843 - 0.821) | 0.834 (0.844 - 0.823) | 0.808 (0.818 - 0.795) | 0.836 (0.847 - 0.824) | 0.835 (0.846 - 0.823) |
| LD | 0.687 (0.693 - 0.680) | 0.697 (0.703 - 0.690) | 0.680 (0.688 - 0.674) | 0.704 (0.711 - 0.697) | 0.706 (0.713 - 0.699) |
| MEL | 0.639 (0.658 - 0.619) | 0.679 (0.696 - 0.662) | 0.607 (0.628 - 0.589) | 0.636 (0.657 - 0.617) | 0.679 (0.696 - 0.661) |
| OP | 0.800 (0.806 - 0.793) | 0.815 (0.822 - 0.809) | 0.781 (0.788 - 0.775) | 0.813 (0.820 - 0.807) | 0.822 (0.829 - 0.816) |
| PAD | 0.767 (0.773 - 0.761) | 0.771 (0.778 - 0.765) | 0.745 (0.753 - 0.737) | 0.774 (0.781 - 0.768) | 0.776 (0.783 - 0.770) |
| PC | 0.687 (0.694 - 0.678) | 0.762 (0.770 - 0.754) | 0.680 (0.688 - 0.671) | 0.688 (0.696 - 0.680) | 0.762 (0.770 - 0.754) |
| PD | 0.767 (0.779 - 0.752) | 0.778 (0.791 - 0.762) | 0.760 (0.773 - 0.747) | 0.768 (0.780 - 0.752) | 0.782 (0.793 - 0.767) |
| POAG | 0.693 (0.702 - 0.680) | 0.736 (0.746 - 0.722) | 0.687 (0.696 - 0.675) | 0.693 (0.703 - 0.681) | 0.736 (0.747 - 0.722) |
| RD | 0.761 (0.765 - 0.757) | 0.774 (0.779 - 0.770) | 0.767 (0.771 - 0.763) | 0.781 (0.785 - 0.777) | 0.782 (0.786 - 0.778) |
| VT | 0.677 (0.688 - 0.666) | 0.710 (0.720 - 0.699) | 0.684 (0.693 - 0.673) | 0.694 (0.704 - 0.683) | 0.718 (0.727 - 0.707) |

**Supplementary Table 2: Absolute performance of the modelled scenarios:** Harrell’s C-index and bootstrapping-derived 95% confidence intervals across heterogenous endpoints and the modelled multi-predictor screening scenarios.

| **Endpoint** | **SCORE-2** | **PREVENT** | **QRISK3** | **No Needle Model** | **Single Blood Draw Model** |
| --- | --- | --- | --- | --- | --- |
| CAD | 0.729 (0.736 - 0.721) | 0.746 (0.752 - 0.738) | 0.752 (0.757 - 0.744) | 0.749 (0.755 - 0.741) | 0.783 (0.789 - 0.776) |
| CVD | 0.699 (0.704 - 0.694) | 0.715 (0.720 - 0.711) | 0.720 (0.724 - 0.715) | 0.731 (0.736 - 0.727) | 0.747 (0.751 - 0.742) |
| HF | 0.757 (0.763 - 0.750) | 0.790 (0.797 - 0.784) | 0.794 (0.800 - 0.787) | 0.809 (0.815 - 0.801) | 0.818 (0.823 - 0.812) |
| ISS | 0.728 (0.739 - 0.720) | 0.744 (0.753 - 0.735) | 0.743 (0.752 - 0.734) | 0.745 (0.754 - 0.736) | 0.756 (0.765 - 0.748) |
| PAD | 0.714 (0.723 - 0.706) | 0.731 (0.740 - 0.723) | 0.738 (0.746 - 0.731) | 0.767 (0.775 - 0.760) | 0.771 (0.779 - 0.763) |

**Supplementary Table 3: Absolute performance for CVD risk stratification models:** Harrell’s C-index and bootstrapping-derived 95% confidence intervals across CVD spectrum endpoints and the predictor matrices benchmarked in this regard.

| **Endpoint** | **No Needle Model** | **Single Blood Draw Model** | **1-Step NHS Health Check** | **1-Step NHS Health Check + PMH/Questionnaires** | **2-Step NHS Health Check (No Needle Model-driven Invitation of > 5% Risk)** | **2-Step NHS Health Check (No Needle Model-driven Invitation of > 5% Risk, Omics for Attendees)** |
| --- | --- | --- | --- | --- | --- | --- |
| CVD | 0.738 (0.745 - 0.731) | 0.756 (0.763 - 0.749) | 0.736 (0.743 - 0.728) | 0.750 (0.757 - 0.743) | 0.741 (0.748 - 0.733) | 0.746 (0.753 - 0.739) |
| DM | 0.768 (0.779 - 0.757) | 0.844 (0.856 - 0.832) | 0.804 (0.815 - 0.793) | 0.820 (0.831 - 0.809) | 0.782 (0.793 - 0.768) | 0.788 (0.801 - 0.771) |
| RD | 0.716 (0.725 - 0.705) | 0.732 (0.742 - 0.722) | 0.693 (0.701 - 0.680) | 0.717 (0.726 - 0.706) | 0.715 (0.725 - 0.704) | 0.722 (0.733 - 0.712) |

**Supplementary Table 4: Absolute performance for the modelled NHS-HC scenarios:** Harrell’s C-index and bootstrapping-derived 95% confidence intervals across the NHS-HC-targeted endpoints and the screening scenarios benchmarked in this regard.

| **Endpoint** | **PMH** | **Self-Reported Questionnaires** | **PRS** | **Metabolomics** |
| --- | --- | --- | --- | --- |
| AAA | 0.797 (0.785 - 0.808) | 0.820 (0.807 - 0.834) | 0.781 (0.768 - 0.793) | 0.796 (0.783 - 0.809) |
| AD | 0.828 (0.819 - 0.838) | 0.850 (0.842 - 0.859) | 0.866 (0.858 - 0.873) | 0.829 (0.819 - 0.837) |
| AF | 0.748 (0.745 - 0.753) | 0.766 (0.762 - 0.770) | 0.759 (0.755 - 0.764) | 0.744 (0.740 - 0.749) |
| AS | 0.626 (0.616 - 0.637) | 0.725 (0.716 - 0.734) | 0.590 (0.579 - 0.601) | 0.609 (0.599 - 0.616) |
| BC | 0.540 (0.529 - 0.551) | 0.561 (0.552 - 0.572) | 0.657 (0.648 - 0.668) | 0.542 (0.530 - 0.552) |
| CAD | 0.718 (0.713 - 0.725) | 0.748 (0.743 - 0.754) | 0.738 (0.732 - 0.744) | 0.733 (0.727 - 0.739) |
| CAT | 0.748 (0.744 - 0.751) | 0.759 (0.755 - 0.762) | 0.743 (0.739 - 0.746) | 0.747 (0.743 - 0.750) |
| COPD | 0.746 (0.739 - 0.751) | 0.859 (0.855 - 0.863) | 0.691 (0.683 - 0.697) | 0.758 (0.754 - 0.763) |
| CRC | 0.659 (0.647 - 0.670) | 0.660 (0.647 - 0.672) | 0.698 (0.686 - 0.709) | 0.664 (0.652 - 0.675) |
| CVD | 0.699 (0.696 - 0.704) | 0.730 (0.726 - 0.734) | 0.699 (0.695 - 0.703) | 0.706 (0.702 - 0.710) |
| DM | 0.685 (0.676 - 0.691) | 0.787 (0.781 - 0.791) | 0.693 (0.686 - 0.700) | 0.798 (0.792 - 0.804) |
| HF | 0.777 (0.771 - 0.785) | 0.807 (0.801 - 0.813) | 0.747 (0.741 - 0.753) | 0.773 (0.767 - 0.779) |
| HT | 0.682 (0.679 - 0.686) | 0.719 (0.716 - 0.723) | 0.679 (0.674 - 0.683) | 0.695 (0.691 - 0.699) |
| ISS | 0.725 (0.715 - 0.734) | 0.744 (0.734 - 0.755) | 0.724 (0.715 - 0.734) | 0.727 (0.717 - 0.736) |
| LC | 0.726 (0.713 - 0.738) | 0.832 (0.821 - 0.844) | 0.692 (0.679 - 0.706) | 0.752 (0.739 - 0.765) |
| LD | 0.654 (0.648 - 0.660) | 0.686 (0.680 - 0.691) | 0.606 (0.599 - 0.612) | 0.660 (0.654 - 0.667) |
| MEL | 0.606 (0.588 - 0.620) | 0.637 (0.620 - 0.652) | 0.660 (0.643 - 0.676) | 0.595 (0.578 - 0.611) |
| OP | 0.774 (0.768 - 0.780) | 0.799 (0.793 - 0.806) | 0.767 (0.760 - 0.774) | 0.767 (0.760 - 0.774) |
| PAD | 0.721 (0.715 - 0.727) | 0.766 (0.759 - 0.772) | 0.695 (0.688 - 0.702) | 0.718 (0.710 - 0.726) |
| PC | 0.678 (0.669 - 0.685) | 0.686 (0.676 - 0.693) | 0.757 (0.750 - 0.765) | 0.678 (0.668 - 0.684) |
| PD | 0.763 (0.750 - 0.776) | 0.769 (0.753 - 0.780) | 0.775 (0.761 - 0.791) | 0.761 (0.746 - 0.776) |
| POAG | 0.694 (0.684 - 0.703) | 0.692 (0.681 - 0.701) | 0.733 (0.722 - 0.743) | 0.686 (0.676 - 0.696) |
| RD | 0.728 (0.724 - 0.732) | 0.759 (0.755 - 0.762) | 0.700 (0.695 - 0.704) | 0.745 (0.741 - 0.749) |
| VT | 0.654 (0.644 - 0.664) | 0.677 (0.667 - 0.688) | 0.670 (0.660 - 0.678) | 0.665 (0.655 - 0.675) |

**Supplementary Table 5: Absolute performance of benchmarked predictor matrices:** Harrell’s C-index and bootstrapping-derived 95% confidence intervals across endpoints and all isolated benchmarked predictor matrices.

| **Endpoint** | **SCORE2** | **PREVENT** | **QRISK3** | **No Needle Model** |
| --- | --- | --- | --- | --- |
| CAD | 0.787 (0.779 - 0.795) | 0.782 (0.773 - 0.792) | 0.844 (0.838 - 0.852) | 0.825 (0.816 - 0.832) |
| CVD | 0.729 (0.722 - 0.737) | 0.759 (0.751 - 0.765) | 0.797 (0.790 - 0.803) | 0.790 (0.784 - 0.796) |
| HF | 0.695 (0.689 - 0.701) | 0.788 (0.783 - 0.793) | 0.827 (0.822 - 0.831) | 0.845 (0.841 - 0.850) |
| ISS | 0.756 (0.746 - 0.767) | 0.801 (0.791 - 0.809) | 0.846 (0.836 - 0.855) | 0.848 (0.839 - 0.855) |
| PAD | 0.594 (0.584 - 0.603) | 0.687 (0.677 - 0.695) | 0.763 (0.752 - 0.770) | 0.783 (0.772 - 0.790) |

**Supplementary Table 6: External validation: Absolute performance validated models in EstBB:** Harrell’s C-index and bootstrapping-derived 95% confidence intervals across CVD spectrum endpoints and all validated models.

| **Predictor** | **Unit** | **Data-Coding** | **Source** | **Field ID** | **Transformation** |
| --- | --- | --- | --- | --- | --- |
| Age at recruitment | Continuous, Years |  | Questionnaire/Interview at recruitment | 21022 | Log-standardscaling (mean=0, std=1) |
| Sex | Boolean | 0 = Female, 1= Male | Questionnaire/Interview at recruitment | 31 | Boolean |

**Supplementary Table 7: Predictors: Age & Sex:** The table details each predictor along with its measurement unit, coding scheme, data source, corresponding UKB Field ID, and any transformation applied prior to modelling. All transformations were estimated within the training fold and then applied to the held-out data to avoid information leakage.

| **Predictor** | **Data-coding** | **Unit** | **Source** | **Field ID** | **Transformation** |
| --- | --- | --- | --- | --- | --- |
| Smoking Status | 2 = Current smoker |  | Questionnaire/Interview at recruitment | 20116 | Boolean |
| Alcohol Intake Frequency | 1 = Daily or almost daily |  | Questionnaire/Interview at recruitment | 1558 | Boolean |
| Sum of Duration of Activity |  | Continuous, Minutes | Questionnaire/Interview at recruitment | 884, 894, 904, 914 | Log - standardscaling (mean=0, std=1) |
| Age Completed Full Time Education | -2 = never went to school | Continuous, Years | Questionnaire/Interview at recruitment | 845 | Tertiles |
| Vegetable and Fruit Intake | 0 = No, 1= Yes |  | Questionnaire/Interview at recruitment | 103990, 104400 | Boolean |
| Illness of Family (Type 2 Diabetes) | 9 = Diabetes |  | Questionnaire/Interview at recruitment | 20107, 20110, 20111 | Boolean |
| Type 2 Diabetes | ICD-10: E10, E11, E12, E13, E14; ICD-9: 250; Non-cancer illness codes: 1220, 1222, 1223 |  | Questionnaire/Interview at recruitment or prior GP/HES records | 20002, 40001, 40002, 41270, 41271 | Boolean |
| Body Mass Index |  | Continuous, / | Physical assessment at recruitment | 21001 | Log - standardscaling (mean=0, std=1) |
| Waist-Hip Ratio |  | Continuous,% | Physical assessment at recruitment | 48, 49 | Log - standardscaling (mean=0, std=1) |
| Waist Circumference |  | Continuous, cm | Physical assessment at recruitment | 48 | Log - standardscaling (mean=0, std=1) |
| Weight |  | Continuous, kg | Physical assessment at recruitment | 21002 | Log - standardscaling (mean=0, std=1) |
| Standing Height |  | Continuous, cm | Physical assessment at recruitment | 50 | Log - standardscaling (mean=0, std=1) |
| Mean Systolic Blood Pressure |  | Continuous, mmHg | Physical assessment at recruitment | 4080, 93 | Log - standardscaling (mean=0, std=1) |
| Total Cholesterol |  | Continuous, mmol/L | Blood laboratory assessment at recruitment | 30690 | Log - standardscaling (mean=0, std=1) |
| LDL Cholesterol |  | Continuous, mmol/L | Blood laboratory assessment at recruitment | 30780 | Log - standardscaling (mean=0, std=1) |
| HDL Cholesterol |  | Continuous, mmol/L | Blood laboratory assessment at recruitment | 30760 | Log - standardscaling (mean=0, std=1) |
| Triglycerides |  | Continuous, mmol/L | Blood laboratory assessment at recruitment | 30870 | Log - standardscaling (mean=0, std=1) |
| Glucose |  | Continuous, mmol/L | Blood laboratory assessment at recruitment | 30740 | Log - standardscaling (mean=0, std=1) |
| Glycated haemoglobin (HbA1c) |  | Continuous, mmol/mol | Blood laboratory assessment at recruitment | 30750 | Log - standardscaling (mean=0, std=1) |
| Aspartate Aminotransferase |  | Continuous, U/L | Blood laboratory assessment at recruitment | 30650 | Log - standardscaling (mean=0, std=1) |
| Alanine Aminotransferase |  | Continuous, U/L | Blood laboratory assessment at recruitment | 30620 | Log - standardscaling (mean=0, std=1) |
| Alkaline Phosphatase |  | Continuous, U/L | Blood laboratory assessment at recruitment | 30610 | Log - standardscaling (mean=0, std=1) |
| Albumin |  | Continuous, g/L | Blood laboratory assessment at recruitment | 30600 | Log - standardscaling (mean=0, std=1) |
| Creatinine |  | Continuous, umol/L | Blood laboratory assessment at recruitment | 30700 | Log - standardscaling (mean=0, std=1) |
| Cystatine C |  | Continuous, mg/L | Blood laboratory assessment at recruitment | 30720 | Log - standardscaling (mean=0, std=1) |
| Urea |  | Continuous, mmol/L | Blood laboratory assessment at recruitment | 30670 | Log - standardscaling (mean=0, std=1) |
| Urate |  | Continuous, umol/L | Blood laboratory assessment at recruitment | 30880 | Log - standardscaling (mean=0, std=1) |
| C-reactive Protein |  | Continuous, mg/L | Blood laboratory assessment at recruitment | 30710 | Log - standardscaling (mean=0, std=1) |
| Red Blood Cell (Erythrocyte) Count |  | Continuous, 10^12 cells/Litre | Blood laboratory assessment at recruitment | 30010 | Log - standardscaling (mean=0, std=1) |
| White Blood Cell (Leukocyte) Count |  | Continuous, 10^9 cells/Litre | Blood laboratory assessment at recruitment | 30000 | Log - standardscaling (mean=0, std=1) |
| Platelet Count |  | Continuous, 10^9 cells/Litre | Blood laboratory assessment at recruitment | 30080 | Log - standardscaling (mean=0, std=1) |
| Haemoglobin Concentration |  | Continuous, grams/decilitre | Blood laboratory assessment at recruitment | 30020 | Log - standardscaling (mean=0, std=1) |
| Hematocrit Percentage |  | Continuous,% | Blood laboratory assessment at recruitment | 30030 | Log - standardscaling (mean=0, std=1) |
| Mean Corpuscular Haemoglobin |  | Continuous, picograms | Blood laboratory assessment at recruitment | 30050 | Log - standardscaling (mean=0, std=1) |
| Mean Corpuscular Volume |  | Continuous, femtolitres | Blood laboratory assessment at recruitment | 30040 | Log - standardscaling (mean=0, std=1) |
| Mean Corpuscular Haemoglobin Concentration |  | Continuous, grams/decilitre | Blood laboratory assessment at recruitment | 30060 | Log - standardscaling (mean=0, std=1) |
| Antihypertensives | 1140860332, 1140860334, 1140860336, 1140860338, 1140860340, 1140860342, 1140860348, 1140860352, 1140860356, 1140860358, 1140860362, 1140860380, 1140860390, 1140860394, 1140860396, 1140860398, 1140860402, 1140860410, 1140860418, 1140860422, 1140860426, 1140860434, 1140860478, 1140860492, 1140860498, 1140860520, 1140860532, 1140860552, 1140860558, 1140860562, 1140860564, 1140860580, 1140860628, 1140860632, 1140860638, 1140860654, 1140860658, 1140860706, 1140860714, 1140860728, 1140860736, 1140860738, 1140860758, 1140860764, 1140860776, 1140860784, 1140860790, 1140860828, 1140860830, 1140860834, 1140860836, 1140860838, 1140860846, 1140860848, 1140860862, 1140860878, 1140860882, 1140860912, 1140860918, 1140860938, 1140860942, 1140860952, 1140860972, 1140860976, 1140860982, 1140860988, 1140860994, 1140861008, 1140861010, 1140861016, 1140861022, 1140861024, 1140861068, 1140861070, 1140861088, 1140861090, 1140861106, 1140861120, 1140861128, 1140861130, 1140861136, 1140861138, 1140861190, 1140861194, 1140861202, 1140861266, 1140861268, 1140861326, 1140861384, 1140864950, 1140864952, 1140866072, 1140866084, 1140866086, 1140866090, 1140866092, 1140866094, 1140866104, 1140866108, 1140866110, 1140866116, 1140866122, 1140866136, 1140866138, 1140866140, 1140866144, 1140866146, 1140866162, 1140866164, 1140866168, 1140866182, 1140866192, 1140866202, 1140866206, 1140866210, 1140866212, 1140866220, 1140866230, 1140866232, 1140866236, 1140866244, 1140866248, 1140866282, 1140866306, 1140866308, 1140866312, 1140866318, 1140866330, 1140866332, 1140866334, 1140866340, 1140866352, 1140866360, 1140866388, 1140866390, 1140866396, 1140866400, 1140866406, 1140866408, 1140866410, 1140866412, 1140866416, 1140866422, 1140866426, 1140866438, 1140866440, 1140866442, 1140866448, 1140866450, 1140866460, 1140866466, 1140866484, 1140866554, 1140866692, 1140866704, 1140866712, 1140866724, 1140866756, 1140866758, 1140866764, 1140866766, 1140866778, 1140866798, 1140866800, 1140866802, 1140866804, 1140875808, 1140879762, 1140879778, 1140879782, 1140879786, 1140879794, 1140879806, 1140879810, 1140879818, 1140879822, 1140879824, 1140879834, 1140879842, 1140879854, 1140879866, 1140888510, 1140888556, 1140888560, 1140888578, 1140888582, 1140888586, 1140888760, 1140888762, 1140909368, 1140911698, 1140916356, 1140923572, 1140923712, 1140923718, 1140926778, 1140926780, 1141145668, 1141151016, 1141151018, 1141151382, 1141152600, 1141153026, 1141153032, 1141153328, 1141156754, 1141156808, 1141157252, 1141157254, 1141164148, 1141164154, 1141164276, 1141165476, 1141166006, 1141167822, 1141167832, 1141171152, 1141172682, 1141172686, 1141172698, 1141173888, 1141180592, 1141187790, 1141190160, 1141192064, 1141193282, 1141193346, 1141194804, 1141194808, 1141194810, 1141201038, 1141201040, 1140860382, 1140860386, 1140860404, 1140860406, 1140860454, 1140860470, 1140860534, 1140860544, 1140860590, 1140860610, 1140860690, 1140860696, 1140860750, 1140860752, 1140860802, 1140860806, 1140860840, 1140860842, 1140860892, 1140860904, 1140860954, 1140860966, 1140861000, 1140861002, 1140861034, 1140861046, 1140861110, 1140861114, 1140861166, 1140861176, 1140861276, 1140861282, 1140866074, 1140866078, 1140866096, 1140866102, 1140866128, 1140866132, 1140866156, 1140866158, 1140866194, 1140866200, 1140866222, 1140866226, 1140866262, 1140866280, 1140866324, 1140866328, 1140866354, 1140866356, 1140866402, 1140866404, 1140866418, 1140866420, 1140866444, 1140866446, 1140866506, 1140866546, 1140866726, 1140866738, 1140866782, 1140866784, 1140879758, 1140879760, 1140879798, 1140879802, 1140879826, 1140879830, 1140888512, 1140888552, 1140888646, 1140888686, 1140916362, 1140917428, 1141145658, 1141145660, 1141152998, 1141153006, 1141156836, 1141156846, 1141164280, 1141165470, 1141171336, 1141171344, 1141180598, 1141187788, 1141194794, 1141194800 | Boolean | Questionnaire/Interview at recruitment | 6153, 6177, 20003 | Boolean |

**Supplementary Table 8: Predictors: Clinical risk:** The table details each predictor along with its measurement unit, coding scheme, data source, corresponding UKB Field ID, and any transformation applied prior to modelling. All transformations were estimated within the training fold and then applied to the held-out data to avoid information leakage.

| **Predictor** | **Unit** | **Source** | **Field ID** |
| --- | --- | --- | --- |
| Standard PRS for Alzheimer’s disease (AD) | Continuous, relative risk | External GWAS training | 26206 |
| Standard PRS for cardiovascular disease (CVD) | Continuous, relative risk | External GWAS training | 26223 |
| Standard PRS for type 1 diabetes (T1D) | Continuous, relative risk | External GWAS training | 26283 |
| Standard PRS for type 2 diabetes (T2D) | Continuous, relative risk | External GWAS training | 26285 |
| Standard PRS for atrial fibrillation (AF) | Continuous, relative risk | External GWAS training | 26212 |
| Standard PRS for coronary artery disease (CAD) | Continuous, relative risk | External GWAS training | 26227 |
| Standard PRS for venous thromboembolic disease (VTE) | Continuous, relative risk | External GWAS training | 26289 |
| Standard PRS for ischaemic stroke (ISS) | Continuous, relative risk | External GWAS training | 26248 |
| Standard PRS for asthma (AST) | Continuous, relative risk | External GWAS training | 26210 |
| Standard PRS for melanoma (MEL) | Continuous, relative risk | External GWAS training | 26252 |
| Standard PRS for bowel cancer (CRC) | Continuous, relative risk | External GWAS training | 26218 |
| Standard PRS for prostate cancer (PC) | Continuous, relative risk | External GWAS training | 26267 |
| Standard PRS for breast cancer (BC) | Continuous, relative risk | External GWAS training | 26220 |
| Standard PRS for Parkinson's disease (PD) | Continuous, relative risk | External GWAS training | 26260 |
| Standard PRS for osteoporosis (OP) | Continuous, relative risk | External GWAS training | 26258 |
| Standard PRS for primary open angle glaucoma (POAG) | Continuous, relative risk | External GWAS training | 26265 |
| Standard PRS for hypertension (HT) | Continuous, relative risk | External GWAS training | 26244 |

**Supplementary Table 9: Predictors: Polygenic Risk Scores:** The table details each predictor along with its measurement unit, coding scheme, data source, and corresponding UKB Field ID. No transformation was applied.

| **Predictor** | **Unit** | **Source** | **Field ID** | **Transformation** |
| --- | --- | --- | --- | --- |
| 3-Hydroxybutyrate | Continuous, mmol/l | Blood laboratory assessment at recruitment | 23474 | Log-standardscaling (mean=0, std=1) |
| Acetate | Continuous, mmol/l | Blood laboratory assessment at recruitment | 23475 | Log-standardscaling (mean=0, std=1) |
| Acetoacetate | Continuous, mmol/l | Blood laboratory assessment at recruitment | 23476 | Log-standardscaling (mean=0, std=1) |
| Acetone | Continuous, mmol/l | Blood laboratory assessment at recruitment | 23477 | Log-standardscaling (mean=0, std=1) |
| Alanine | Continuous, mmol/l | Blood laboratory assessment at recruitment | 23460 | Log-standardscaling (mean=0, std=1) |
| Albumin | Continuous, g/l | Blood laboratory assessment at recruitment | 23479 | Log-standardscaling (mean=0, std=1) |
| Apolipoprotein A1 | Continuous, g/l | Blood laboratory assessment at recruitment | 23440 | Log-standardscaling (mean=0, std=1) |
| Apolipoprotein B | Continuous, g/l | Blood laboratory assessment at recruitment | 23439 | Log-standardscaling (mean=0, std=1) |
| Average Diameter for HDL Particles | Continuous, nm | Blood laboratory assessment at recruitment | 23433 | Log-standardscaling (mean=0, std=1) |
| Average Diameter for LDL Particles | Continuous, nm | Blood laboratory assessment at recruitment | 23432 | Log-standardscaling (mean=0, std=1) |
| Average Diameter for VLDL Particles | Continuous, nm | Blood laboratory assessment at recruitment | 23431 | Log-standardscaling (mean=0, std=1) |
| Cholesterol in Chylomicrons and Extremely Large VLDL | Continuous, mmol/l | Blood laboratory assessment at recruitment | 23484 | Log-standardscaling (mean=0, std=1) |
| Cholesterol in IDL | Continuous, mmol/l | Blood laboratory assessment at recruitment | 23526 | Log-standardscaling (mean=0, std=1) |
| Cholesterol in Large HDL | Continuous, mmol/l | Blood laboratory assessment at recruitment | 23561 | Log-standardscaling (mean=0, std=1) |
| Cholesterol in Large LDL | Continuous, mmol/l | Blood laboratory assessment at recruitment | 23533 | Log-standardscaling (mean=0, std=1) |
| Cholesterol in Large VLDL | Continuous, mmol/l | Blood laboratory assessment at recruitment | 23498 | Log-standardscaling (mean=0, std=1) |
| Cholesterol in Medium HDL | Continuous, mmol/l | Blood laboratory assessment at recruitment | 23568 | Log-standardscaling (mean=0, std=1) |
| Cholesterol in Medium LDL | Continuous, mmol/l | Blood laboratory assessment at recruitment | 23540 | Log-standardscaling (mean=0, std=1) |
| Cholesterol in Medium VLDL | Continuous, mmol/l | Blood laboratory assessment at recruitment | 23505 | Log-standardscaling (mean=0, std=1) |
| Cholesterol in Small HDL | Continuous, mmol/l | Blood laboratory assessment at recruitment | 23575 | Log-standardscaling (mean=0, std=1) |
| Cholesterol in Small LDL | Continuous, mmol/l | Blood laboratory assessment at recruitment | 23547 | Log-standardscaling (mean=0, std=1) |
| Cholesterol in Small VLDL | Continuous, mmol/l | Blood laboratory assessment at recruitment | 23512 | Log-standardscaling (mean=0, std=1) |
| Cholesterol in Very Large HDL | Continuous, mmol/l | Blood laboratory assessment at recruitment | 23554 | Log-standardscaling (mean=0, std=1) |
| Cholesterol in Very Large VLDL | Continuous, mmol/l | Blood laboratory assessment at recruitment | 23491 | Log-standardscaling (mean=0, std=1) |
| Cholesterol in Very Small VLDL | Continuous, mmol/l | Blood laboratory assessment at recruitment | 23519 | Log-standardscaling (mean=0, std=1) |
| Cholesteryl Esters in Chylomicrons and Extremely Large VLDL | Continuous, mmol/l | Blood laboratory assessment at recruitment | 23485 | Log-standardscaling (mean=0, std=1) |
| Cholesteryl Esters in HDL | Continuous, mmol/l | Blood laboratory assessment at recruitment | 23418 | Log-standardscaling (mean=0, std=1) |
| Cholesteryl Esters in IDL | Continuous, mmol/l | Blood laboratory assessment at recruitment | 23527 | Log-standardscaling (mean=0, std=1) |
| Cholesteryl Esters in Large HDL | Continuous, mmol/l | Blood laboratory assessment at recruitment | 23562 | Log-standardscaling (mean=0, std=1) |
| Cholesteryl Esters in Large LDL | Continuous, mmol/l | Blood laboratory assessment at recruitment | 23534 | Log-standardscaling (mean=0, std=1) |
| Cholesteryl Esters in Large VLDL | Continuous, mmol/l | Blood laboratory assessment at recruitment | 23499 | Log-standardscaling (mean=0, std=1) |
| Cholesteryl Esters in LDL | Continuous, mmol/l | Blood laboratory assessment at recruitment | 23417 | Log-standardscaling (mean=0, std=1) |
| Cholesteryl Esters in Medium HDL | Continuous, mmol/l | Blood laboratory assessment at recruitment | 23569 | Log-standardscaling (mean=0, std=1) |
| Cholesteryl Esters in Medium LDL | Continuous, mmol/l | Blood laboratory assessment at recruitment | 23541 | Log-standardscaling (mean=0, std=1) |
| Cholesteryl Esters in Medium VLDL | Continuous, mmol/l | Blood laboratory assessment at recruitment | 23506 | Log-standardscaling (mean=0, std=1) |
| Cholesteryl Esters in Small HDL | Continuous, mmol/l | Blood laboratory assessment at recruitment | 23576 | Log-standardscaling (mean=0, std=1) |
| Cholesteryl Esters in Small LDL | Continuous, mmol/l | Blood laboratory assessment at recruitment | 23548 | Log-standardscaling (mean=0, std=1) |
| Cholesteryl Esters in Small VLDL | Continuous, mmol/l | Blood laboratory assessment at recruitment | 23513 | Log-standardscaling (mean=0, std=1) |
| Cholesteryl Esters in Very Large HDL | Continuous, mmol/l | Blood laboratory assessment at recruitment | 23555 | Log-standardscaling (mean=0, std=1) |
| Cholesteryl Esters in Very Large VLDL | Continuous, mmol/l | Blood laboratory assessment at recruitment | 23492 | Log-standardscaling (mean=0, std=1) |
| Cholesteryl Esters in Very Small VLDL | Continuous, mmol/l | Blood laboratory assessment at recruitment | 23520 | Log-standardscaling (mean=0, std=1) |
| Cholesteryl Esters in VLDL | Continuous, mmol/l | Blood laboratory assessment at recruitment | 23416 | Log-standardscaling (mean=0, std=1) |
| Citrate | Continuous, mmol/l | Blood laboratory assessment at recruitment | 23473 | Log-standardscaling (mean=0, std=1) |
| Clinical LDL Cholesterol | Continuous, mmol/l | Blood laboratory assessment at recruitment | 23404 | Log-standardscaling (mean=0, std=1) |
| Concentration of Chylomicrons and Extremely Large VLDL Particles | Continuous, mmol/l | Blood laboratory assessment at recruitment | 23481 | Log-standardscaling (mean=0, std=1) |
| Concentration of HDL Particles | Continuous, mmol/l | Blood laboratory assessment at recruitment | 23430 | Log-standardscaling (mean=0, std=1) |
| Concentration of IDL Particles | Continuous, mmol/l | Blood laboratory assessment at recruitment | 23523 | Log-standardscaling (mean=0, std=1) |
| Concentration of Large HDL Particles | Continuous, mmol/l | Blood laboratory assessment at recruitment | 23558 | Log-standardscaling (mean=0, std=1) |
| Concentration of Large LDL Particles | Continuous, mmol/l | Blood laboratory assessment at recruitment | 23530 | Log-standardscaling (mean=0, std=1) |
| Concentration of Large VLDL Particles | Continuous, mmol/l | Blood laboratory assessment at recruitment | 23495 | Log-standardscaling (mean=0, std=1) |
| Concentration of LDL Particles | Continuous, mmol/l | Blood laboratory assessment at recruitment | 23429 | Log-standardscaling (mean=0, std=1) |
| Concentration of Medium HDL Particles | Continuous, mmol/l | Blood laboratory assessment at recruitment | 23565 | Log-standardscaling (mean=0, std=1) |
| Concentration of Medium LDL Particles | Continuous, mmol/l | Blood laboratory assessment at recruitment | 23537 | Log-standardscaling (mean=0, std=1) |
| Concentration of Medium VLDL Particles | Continuous, mmol/l | Blood laboratory assessment at recruitment | 23502 | Log-standardscaling (mean=0, std=1) |
| Concentration of Small HDL Particles | Continuous, mmol/l | Blood laboratory assessment at recruitment | 23572 | Log-standardscaling (mean=0, std=1) |
| Concentration of Small LDL Particles | Continuous, mmol/l | Blood laboratory assessment at recruitment | 23544 | Log-standardscaling (mean=0, std=1) |
| Concentration of Small VLDL Particles | Continuous, mmol/l | Blood laboratory assessment at recruitment | 23509 | Log-standardscaling (mean=0, std=1) |
| Concentration of Very Large HDL Particles | Continuous, mmol/l | Blood laboratory assessment at recruitment | 23551 | Log-standardscaling (mean=0, std=1) |
| Concentration of Very Large VLDL Particles | Continuous, mmol/l | Blood laboratory assessment at recruitment | 23488 | Log-standardscaling (mean=0, std=1) |
| Concentration of Very Small VLDL Particles | Continuous, mmol/l | Blood laboratory assessment at recruitment | 23516 | Log-standardscaling (mean=0, std=1) |
| Concentration of VLDL Particles | Continuous, mmol/l | Blood laboratory assessment at recruitment | 23428 | Log-standardscaling (mean=0, std=1) |
| Creatinine | Continuous, mmol/l | Blood laboratory assessment at recruitment | 23478 | Log-standardscaling (mean=0, std=1) |
| Degree of Unsaturation | Continuous, degree | Blood laboratory assessment at recruitment | 23443 | Log-standardscaling (mean=0, std=1) |
| Docosahexaenoic Acid | Continuous, mmol/l | Blood laboratory assessment at recruitment | 23450 | Log-standardscaling (mean=0, std=1) |
| Free Cholesterol in Chylomicrons and Extremely Large VLDL | Continuous, mmol/l | Blood laboratory assessment at recruitment | 23486 | Log-standardscaling (mean=0, std=1) |
| Free Cholesterol in HDL | Continuous, mmol/l | Blood laboratory assessment at recruitment | 23422 | Log-standardscaling (mean=0, std=1) |
| Free Cholesterol in IDL | Continuous, mmol/l | Blood laboratory assessment at recruitment | 23528 | Log-standardscaling (mean=0, std=1) |
| Free Cholesterol in Large HDL | Continuous, mmol/l | Blood laboratory assessment at recruitment | 23563 | Log-standardscaling (mean=0, std=1) |
| Free Cholesterol in Large LDL | Continuous, mmol/l | Blood laboratory assessment at recruitment | 23535 | Log-standardscaling (mean=0, std=1) |
| Free Cholesterol in Large VLDL | Continuous, mmol/l | Blood laboratory assessment at recruitment | 23500 | Log-standardscaling (mean=0, std=1) |
| Free Cholesterol in LDL | Continuous, mmol/l | Blood laboratory assessment at recruitment | 23421 | Log-standardscaling (mean=0, std=1) |
| Free Cholesterol in Medium HDL | Continuous, mmol/l | Blood laboratory assessment at recruitment | 23570 | Log-standardscaling (mean=0, std=1) |
| Free Cholesterol in Medium LDL | Continuous, mmol/l | Blood laboratory assessment at recruitment | 23542 | Log-standardscaling (mean=0, std=1) |
| Free Cholesterol in Medium VLDL | Continuous, mmol/l | Blood laboratory assessment at recruitment | 23507 | Log-standardscaling (mean=0, std=1) |
| Free Cholesterol in Small HDL | Continuous, mmol/l | Blood laboratory assessment at recruitment | 23577 | Log-standardscaling (mean=0, std=1) |
| Free Cholesterol in Small LDL | Continuous, mmol/l | Blood laboratory assessment at recruitment | 23549 | Log-standardscaling (mean=0, std=1) |
| Free Cholesterol in Small VLDL | Continuous, mmol/l | Blood laboratory assessment at recruitment | 23514 | Log-standardscaling (mean=0, std=1) |
| Free Cholesterol in Very Large HDL | Continuous, mmol/l | Blood laboratory assessment at recruitment | 23556 | Log-standardscaling (mean=0, std=1) |
| Free Cholesterol in Very Large VLDL | Continuous, mmol/l | Blood laboratory assessment at recruitment | 23493 | Log-standardscaling (mean=0, std=1) |
| Free Cholesterol in Very Small VLDL | Continuous, mmol/l | Blood laboratory assessment at recruitment | 23521 | Log-standardscaling (mean=0, std=1) |
| Free Cholesterol in VLDL | Continuous, mmol/l | Blood laboratory assessment at recruitment | 23420 | Log-standardscaling (mean=0, std=1) |
| Glucose | Continuous, mmol/l | Blood laboratory assessment at recruitment | 23470 | Log-standardscaling (mean=0, std=1) |
| Glutamine | Continuous, mmol/l | Blood laboratory assessment at recruitment | 23461 | Log-standardscaling (mean=0, std=1) |
| Glycine | Continuous, mmol/l | Blood laboratory assessment at recruitment | 23462 | Log-standardscaling (mean=0, std=1) |
| Glycoprotein Acetyls | Continuous, mmol/l | Blood laboratory assessment at recruitment | 23480 | Log-standardscaling (mean=0, std=1) |
| HDL Cholesterol | Continuous, mmol/l | Blood laboratory assessment at recruitment | 23406 | Log-standardscaling (mean=0, std=1) |
| Histidine | Continuous, mmol/l | Blood laboratory assessment at recruitment | 23463 | Log-standardscaling (mean=0, std=1) |
| Isoleucine | Continuous, mmol/l | Blood laboratory assessment at recruitment | 23465 | Log-standardscaling (mean=0, std=1) |
| Lactate | Continuous, mmol/l | Blood laboratory assessment at recruitment | 23471 | Log-standardscaling (mean=0, std=1) |
| LDL Cholesterol | Continuous, mmol/l | Blood laboratory assessment at recruitment | 23405 | Log-standardscaling (mean=0, std=1) |
| Leucine | Continuous, mmol/l | Blood laboratory assessment at recruitment | 23466 | Log-standardscaling (mean=0, std=1) |
| Linoleic Acid | Continuous, mmol/l | Blood laboratory assessment at recruitment | 23449 | Log-standardscaling (mean=0, std=1) |
| Monounsaturated Fatty Acids | Continuous, mmol/l | Blood laboratory assessment at recruitment | 23447 | Log-standardscaling (mean=0, std=1) |
| Omega-3 Fatty Acids | Continuous, mmol/l | Blood laboratory assessment at recruitment | 23444 | Log-standardscaling (mean=0, std=1) |
| Omega-6 Fatty Acids | Continuous, mmol/l | Blood laboratory assessment at recruitment | 23445 | Log-standardscaling (mean=0, std=1) |
| Phenylalanine | Continuous, mmol/l | Blood laboratory assessment at recruitment | 23468 | Log-standardscaling (mean=0, std=1) |
| Phosphatidylcholines | Continuous, mmol/l | Blood laboratory assessment at recruitment | 23437 | Log-standardscaling (mean=0, std=1) |
| Phosphoglycerides | Continuous, mmol/l | Blood laboratory assessment at recruitment | 23434 | Log-standardscaling (mean=0, std=1) |
| Phospholipids in Chylomicrons and Extremely Large VLDL | Continuous, mmol/l | Blood laboratory assessment at recruitment | 23483 | Log-standardscaling (mean=0, std=1) |
| Phospholipids in HDL | Continuous, mmol/l | Blood laboratory assessment at recruitment | 23414 | Log-standardscaling (mean=0, std=1) |
| Phospholipids in IDL | Continuous, mmol/l | Blood laboratory assessment at recruitment | 23525 | Log-standardscaling (mean=0, std=1) |
| Phospholipids in Large HDL | Continuous, mmol/l | Blood laboratory assessment at recruitment | 23560 | Log-standardscaling (mean=0, std=1) |
| Phospholipids in Large LDL | Continuous, mmol/l | Blood laboratory assessment at recruitment | 23532 | Log-standardscaling (mean=0, std=1) |
| Phospholipids in Large VLDL | Continuous, mmol/l | Blood laboratory assessment at recruitment | 23497 | Log-standardscaling (mean=0, std=1) |
| Phospholipids in LDL | Continuous, mmol/l | Blood laboratory assessment at recruitment | 23413 | Log-standardscaling (mean=0, std=1) |
| Phospholipids in Medium HDL | Continuous, mmol/l | Blood laboratory assessment at recruitment | 23567 | Log-standardscaling (mean=0, std=1) |
| Phospholipids in Medium LDL | Continuous, mmol/l | Blood laboratory assessment at recruitment | 23539 | Log-standardscaling (mean=0, std=1) |
| Phospholipids in Medium VLDL | Continuous, mmol/l | Blood laboratory assessment at recruitment | 23504 | Log-standardscaling (mean=0, std=1) |
| Phospholipids in Small HDL | Continuous, mmol/l | Blood laboratory assessment at recruitment | 23574 | Log-standardscaling (mean=0, std=1) |
| Phospholipids in Small LDL | Continuous, mmol/l | Blood laboratory assessment at recruitment | 23546 | Log-standardscaling (mean=0, std=1) |
| Phospholipids in Small VLDL | Continuous, mmol/l | Blood laboratory assessment at recruitment | 23511 | Log-standardscaling (mean=0, std=1) |
| Phospholipids in Very Large HDL | Continuous, mmol/l | Blood laboratory assessment at recruitment | 23553 | Log-standardscaling (mean=0, std=1) |
| Phospholipids in Very Large VLDL | Continuous, mmol/l | Blood laboratory assessment at recruitment | 23490 | Log-standardscaling (mean=0, std=1) |
| Phospholipids in Very Small VLDL | Continuous, mmol/l | Blood laboratory assessment at recruitment | 23518 | Log-standardscaling (mean=0, std=1) |
| Phospholipids in VLDL | Continuous, mmol/l | Blood laboratory assessment at recruitment | 23412 | Log-standardscaling (mean=0, std=1) |
| Polyunsaturated Fatty Acids | Continuous, mmol/l | Blood laboratory assessment at recruitment | 23446 | Log-standardscaling (mean=0, std=1) |
| Pyruvate | Continuous, mmol/l | Blood laboratory assessment at recruitment | 23472 | Log-standardscaling (mean=0, std=1) |
| Remnant Cholesterol (Non-HDL, Non-LDL -Cholesterol) | Continuous, mmol/l | Blood laboratory assessment at recruitment | 23402 | Log-standardscaling (mean=0, std=1) |
| Saturated Fatty Acids | Continuous, mmol/l | Blood laboratory assessment at recruitment | 23448 | Log-standardscaling (mean=0, std=1) |
| Sphingomyelins | Continuous, mmol/l | Blood laboratory assessment at recruitment | 23438 | Log-standardscaling (mean=0, std=1) |
| Total Cholesterol | Continuous, mmol/l | Blood laboratory assessment at recruitment | 23400 | Log-standardscaling (mean=0, std=1) |
| Total Cholesterol Minus HDL-C | Continuous, mmol/l | Blood laboratory assessment at recruitment | 23401 | Log-standardscaling (mean=0, std=1) |
| Total Cholines | Continuous, mmol/l | Blood laboratory assessment at recruitment | 23436 | Log-standardscaling (mean=0, std=1) |
| Total Concentration of Branched-Chain Amino Acids (Leucine + Isoleucine + Valine) | Continuous, mmol/l | Blood laboratory assessment at recruitment | 23464 | Log-standardscaling (mean=0, std=1) |
| Total Concentration of Lipoprotein Particles | Continuous, mmol/l | Blood laboratory assessment at recruitment | 23427 | Log-standardscaling (mean=0, std=1) |
| Total Esterified Cholesterol | Continuous, mmol/l | Blood laboratory assessment at recruitment | 23415 | Log-standardscaling (mean=0, std=1) |
| Total Fatty Acids | Continuous, mmol/l | Blood laboratory assessment at recruitment | 23442 | Log-standardscaling (mean=0, std=1) |
| Total Free Cholesterol | Continuous, mmol/l | Blood laboratory assessment at recruitment | 23419 | Log-standardscaling (mean=0, std=1) |
| Total Lipids in Chylomicrons and Extremely Large VLDL | Continuous, mmol/l | Blood laboratory assessment at recruitment | 23482 | Log-standardscaling (mean=0, std=1) |
| Total Lipids in HDL | Continuous, mmol/l | Blood laboratory assessment at recruitment | 23426 | Log-standardscaling (mean=0, std=1) |
| Total Lipids in IDL | Continuous, mmol/l | Blood laboratory assessment at recruitment | 23524 | Log-standardscaling (mean=0, std=1) |
| Total Lipids in Large HDL | Continuous, mmol/l | Blood laboratory assessment at recruitment | 23559 | Log-standardscaling (mean=0, std=1) |
| Total Lipids in Large LDL | Continuous, mmol/l | Blood laboratory assessment at recruitment | 23531 | Log-standardscaling (mean=0, std=1) |
| Total Lipids in Large VLDL | Continuous, mmol/l | Blood laboratory assessment at recruitment | 23496 | Log-standardscaling (mean=0, std=1) |
| Total Lipids in LDL | Continuous, mmol/l | Blood laboratory assessment at recruitment | 23425 | Log-standardscaling (mean=0, std=1) |
| Total Lipids in Lipoprotein Particles | Continuous, mmol/l | Blood laboratory assessment at recruitment | 23423 | Log-standardscaling (mean=0, std=1) |
| Total Lipids in Medium HDL | Continuous, mmol/l | Blood laboratory assessment at recruitment | 23566 | Log-standardscaling (mean=0, std=1) |
| Total Lipids in Medium LDL | Continuous, mmol/l | Blood laboratory assessment at recruitment | 23538 | Log-standardscaling (mean=0, std=1) |
| Total Lipids in Medium VLDL | Continuous, mmol/l | Blood laboratory assessment at recruitment | 23503 | Log-standardscaling (mean=0, std=1) |
| Total Lipids in Small HDL | Continuous, mmol/l | Blood laboratory assessment at recruitment | 23573 | Log-standardscaling (mean=0, std=1) |
| Total Lipids in Small LDL | Continuous, mmol/l | Blood laboratory assessment at recruitment | 23545 | Log-standardscaling (mean=0, std=1) |
| Total Lipids in Small VLDL | Continuous, mmol/l | Blood laboratory assessment at recruitment | 23510 | Log-standardscaling (mean=0, std=1) |
| Total Lipids in Very Large HDL | Continuous, mmol/l | Blood laboratory assessment at recruitment | 23552 | Log-standardscaling (mean=0, std=1) |
| Total Lipids in Very Large VLDL | Continuous, mmol/l | Blood laboratory assessment at recruitment | 23489 | Log-standardscaling (mean=0, std=1) |
| Total Lipids in Very Small VLDL | Continuous, mmol/l | Blood laboratory assessment at recruitment | 23517 | Log-standardscaling (mean=0, std=1) |
| Total Lipids in VLDL | Continuous, mmol/l | Blood laboratory assessment at recruitment | 23424 | Log-standardscaling (mean=0, std=1) |
| Total Phospholipids in Lipoprotein Particles | Continuous, mmol/l | Blood laboratory assessment at recruitment | 23411 | Log-standardscaling (mean=0, std=1) |
| Total Triglycerides | Continuous, mmol/l | Blood laboratory assessment at recruitment | 23407 | Log-standardscaling (mean=0, std=1) |
| Triglycerides in Chylomicrons and Extremely Large VLDL | Continuous, mmol/l | Blood laboratory assessment at recruitment | 23487 | Log-standardscaling (mean=0, std=1) |
| Triglycerides in HDL | Continuous, mmol/l | Blood laboratory assessment at recruitment | 23410 | Log-standardscaling (mean=0, std=1) |
| Triglycerides in IDL | Continuous, mmol/l | Blood laboratory assessment at recruitment | 23529 | Log-standardscaling (mean=0, std=1) |
| Triglycerides in Large HDL | Continuous, mmol/l | Blood laboratory assessment at recruitment | 23564 | Log-standardscaling (mean=0, std=1) |
| Triglycerides in Large LDL | Continuous, mmol/l | Blood laboratory assessment at recruitment | 23536 | Log-standardscaling (mean=0, std=1) |
| Triglycerides in Large VLDL | Continuous, mmol/l | Blood laboratory assessment at recruitment | 23501 | Log-standardscaling (mean=0, std=1) |
| Triglycerides in LDL | Continuous, mmol/l | Blood laboratory assessment at recruitment | 23409 | Log-standardscaling (mean=0, std=1) |
| Triglycerides in Medium HDL | Continuous, mmol/l | Blood laboratory assessment at recruitment | 23571 | Log-standardscaling (mean=0, std=1) |
| Triglycerides in Medium LDL | Continuous, mmol/l | Blood laboratory assessment at recruitment | 23543 | Log-standardscaling (mean=0, std=1) |
| Triglycerides in Medium VLDL | Continuous, mmol/l | Blood laboratory assessment at recruitment | 23508 | Log-standardscaling (mean=0, std=1) |
| Triglycerides in Small HDL | Continuous, mmol/l | Blood laboratory assessment at recruitment | 23578 | Log-standardscaling (mean=0, std=1) |
| Triglycerides in Small LDL | Continuous, mmol/l | Blood laboratory assessment at recruitment | 23550 | Log-standardscaling (mean=0, std=1) |
| Triglycerides in Small VLDL | Continuous, mmol/l | Blood laboratory assessment at recruitment | 23515 | Log-standardscaling (mean=0, std=1) |
| Triglycerides in Very Large HDL | Continuous, mmol/l | Blood laboratory assessment at recruitment | 23557 | Log-standardscaling (mean=0, std=1) |
| Triglycerides in Very Large VLDL | Continuous, mmol/l | Blood laboratory assessment at recruitment | 23494 | Log-standardscaling (mean=0, std=1) |
| Triglycerides in Very Small VLDL | Continuous, mmol/l | Blood laboratory assessment at recruitment | 23522 | Log-standardscaling (mean=0, std=1) |
| Triglycerides in VLDL | Continuous, mmol/l | Blood laboratory assessment at recruitment | 23408 | Log-standardscaling (mean=0, std=1) |
| Tyrosine | Continuous, mmol/l | Blood laboratory assessment at recruitment | 23469 | Log-standardscaling (mean=0, std=1) |
| Valine | Continuous, mmol/l | Blood laboratory assessment at recruitment | 23467 | Log-standardscaling (mean=0, std=1) |
| VLDL Cholesterol | Continuous, mmol/l | Blood laboratory assessment at recruitment | 23403 | Log-standardscaling (mean=0, std=1) |

**Supplementary Table 10: Predictors: 1H-NMR Metabolomics:** The table details each predictor along with its measurement unit, coding scheme, data source, corresponding UKB Field ID, and any transformation applied prior to modelling. All transformations were estimated within the training fold and then applied to the held-out data to avoid information leakage.

| **Predictor (ICD-10 codes)** | **Data coding** | **Source** | **Field ID** | **Transformation** |
| --- | --- | --- | --- | --- |
| A04 | Other bacterial intestinal infections | Prior HES records | 41270 | Boolean |
| A41 | Other septicaemia | Prior HES records | 41270 | Boolean |
| B07 | Viral warts | Prior HES records | 41270 | Boolean |
| B34 | Viral infection of unspecified site | Prior HES records | 41270 | Boolean |
| B37 | Candidiasis | Prior HES records | 41270 | Boolean |
| B95 | Streptococcus and staphylococcus as the cause of diseases classified to other chapters | Prior HES records | 41270 | Boolean |
| B96 | Other bacterial agents as the cause of diseases classified to other chapters | Prior HES records | 41270 | Boolean |
| C18 | Malignant neoplasm of colon | Prior HES records | 41270 | Boolean |
| C20 | Malignant neoplasm of rectum | Prior HES records | 41270 | Boolean |
| C43 | Malignant melanoma of skin | Prior HES records | 41270 | Boolean |
| C44 | Other malignant neoplasms of skin | Prior HES records | 41270 | Boolean |
| C50 | Malignant neoplasm of breast | Prior HES records | 41270 | Boolean |
| C54 | Malignant neoplasm of corpus uteri | Prior HES records | 41270 | Boolean |
| C61 | Malignant neoplasm of prostate | Prior HES records | 41270 | Boolean |
| C67 | Malignant neoplasm of bladder | Prior HES records | 41270 | Boolean |
| C77 | Secondary and unspecified malignant neoplasm of lymph nodes | Prior HES records | 41270 | Boolean |
| C78 | Secondary malignant neoplasm of respiratory and digestive organs | Prior HES records | 41270 | Boolean |
| C85 | Other and unspecified types of non-Hodgkin's lymphoma | Prior HES records | 41270 | Boolean |
| D05 | Carcinoma in situ of breast | Prior HES records | 41270 | Boolean |
| D06 | Carcinoma in situ of cervix uteri | Prior HES records | 41270 | Boolean |
| D12 | Benign neoplasm of colon, rectum, anus and anal canal | Prior HES records | 41270 | Boolean |
| D13 | Benign neoplasm of other and ill-defined parts of digestive system | Prior HES records | 41270 | Boolean |
| D17 | Benign lipomatous neoplasm | Prior HES records | 41270 | Boolean |
| D18 | Haemangioma and lymphangioma, any site | Prior HES records | 41270 | Boolean |
| D22 | Melanocytic naevi | Prior HES records | 41270 | Boolean |
| D23 | Other benign neoplasms of skin | Prior HES records | 41270 | Boolean |
| D24 | Benign neoplasm of breast | Prior HES records | 41270 | Boolean |
| D25 | Leiomyoma of uterus | Prior HES records | 41270 | Boolean |
| D27 | Benign neoplasm of ovary | Prior HES records | 41270 | Boolean |
| D36 | Benign neoplasm of other and unspecified sites | Prior HES records | 41270 | Boolean |
| D37 | Neoplasm of uncertain or unknown behaviour of oral cavity and digestive organs | Prior HES records | 41270 | Boolean |
| D48 | Neoplasm of uncertain or unknown behaviour of other and unspecified sites | Prior HES records | 41270 | Boolean |
| D50 | Iron deficiency anaemia | Prior HES records | 41270 | Boolean |
| D64 | Other anaemias | Prior HES records | 41270 | Boolean |
| D68 | Other coagulation defects | Prior HES records | 41270 | Boolean |
| D69 | Purpura and other haemorrhagic conditions | Prior HES records | 41270 | Boolean |
| D70 | Agranulocytosis | Prior HES records | 41270 | Boolean |
| E03 | Other hypothyroidism | Prior HES records | 41270 | Boolean |
| E04 | Other non-toxic goitre | Prior HES records | 41270 | Boolean |
| E05 | Thyrotoxicosis [hyperthyroidism] | Prior HES records | 41270 | Boolean |
| E10 | Insulin-dependent diabetes mellitus | Prior HES records | 41270 | Boolean |
| E11 | Non-insulin-dependent diabetes mellitus | Prior HES records | 41270 | Boolean |
| E14 | Unspecified diabetes mellitus | Prior HES records | 41270 | Boolean |
| E66 | Obesity | Prior HES records | 41270 | Boolean |
| E78 | Disorders of lipoprotein metabolism and other lipidaemias | Prior HES records | 41270 | Boolean |
| E83 | Disorders of mineral metabolism | Prior HES records | 41270 | Boolean |
| E86 | Volume depletion | Prior HES records | 41270 | Boolean |
| E87 | Other disorders of fluid, electrolyte and acid-base balance | Prior HES records | 41270 | Boolean |
| E89 | Postprocedural endocrine and metabolic disorders, not elsewhere classified | Prior HES records | 41270 | Boolean |
| F10 | Mental and behavioural disorders due to use of alcohol | Prior HES records | 41270 | Boolean |
| F17 | Mental and behavioural disorders due to use of tobacco | Prior HES records | 41270 | Boolean |
| F20 | Schizophrenia | Prior HES records | 41270 | Boolean |
| F31 | Bipolar affective disorder | Prior HES records | 41270 | Boolean |
| F32 | Depressive episode | Prior HES records | 41270 | Boolean |
| F33 | Recurrent depressive disorder | Prior HES records | 41270 | Boolean |
| F41 | Other anxiety disorders | Prior HES records | 41270 | Boolean |
| G35 | Multiple sclerosis | Prior HES records | 41270 | Boolean |
| G40 | Epilepsy | Prior HES records | 41270 | Boolean |
| G43 | Migraine | Prior HES records | 41270 | Boolean |
| G44 | Other headache syndromes | Prior HES records | 41270 | Boolean |
| G45 | Transient cerebral ischaemic attacks and related syndromes | Prior HES records | 41270 | Boolean |
| G47 | Sleep disorders | Prior HES records | 41270 | Boolean |
| G51 | Facial nerve disorders | Prior HES records | 41270 | Boolean |
| G55 | Nerve root and plexus compressions in diseases classified elsewhere | Prior HES records | 41270 | Boolean |
| G56 | Mononeuropathies of upper limb | Prior HES records | 41270 | Boolean |
| G57 | Mononeuropathies of lower limb | Prior HES records | 41270 | Boolean |
| G81 | Hemiplegia | Prior HES records | 41270 | Boolean |
| G93 | Other disorders of brain | Prior HES records | 41270 | Boolean |
| G99 | Other disorders of nervous system in diseases classified elsewhere | Prior HES records | 41270 | Boolean |
| H00 | Hordeolum and chalazion | Prior HES records | 41270 | Boolean |
| H02 | Other disorders of eyelid | Prior HES records | 41270 | Boolean |
| H04 | Disorders of lachrymal system | Prior HES records | 41270 | Boolean |
| H11 | Other disorders of conjunctiva | Prior HES records | 41270 | Boolean |
| H25 | Senile cataract | Prior HES records | 41270 | Boolean |
| H26 | Other cataract | Prior HES records | 41270 | Boolean |
| H33 | Retinal detachments and breaks | Prior HES records | 41270 | Boolean |
| H35 | Other retinal disorders | Prior HES records | 41270 | Boolean |
| H36 | Retinal disorders in diseases classified elsewhere | Prior HES records | 41270 | Boolean |
| H40 | Glaucoma | Prior HES records | 41270 | Boolean |
| H43 | Disorders of vitreous body | Prior HES records | 41270 | Boolean |
| H50 | Other strabismus | Prior HES records | 41270 | Boolean |
| H52 | Disorders of refraction and accommodation | Prior HES records | 41270 | Boolean |
| H53 | Visual disturbances | Prior HES records | 41270 | Boolean |
| H61 | Other disorders of external ear | Prior HES records | 41270 | Boolean |
| H65 | Nonsuppurative otitis media | Prior HES records | 41270 | Boolean |
| H66 | Suppurative and unspecified otitis media | Prior HES records | 41270 | Boolean |
| H72 | Perforation of tympanic membrane | Prior HES records | 41270 | Boolean |
| H91 | Other hearing loss | Prior HES records | 41270 | Boolean |
| I10 | Essential (primary) hypertension | Prior HES records | 41270 | Boolean |
| I12 | Hypertensive renal disease | Prior HES records | 41270 | Boolean |
| I20 | Angina pectoris | Prior HES records | 41270 | Boolean |
| I21 | Acute myocardial infarction | Prior HES records | 41270 | Boolean |
| I22 | Subsequent myocardial infarction | Prior HES records | 41270 | Boolean |
| I25 | Chronic ischaemic heart disease | Prior HES records | 41270 | Boolean |
| I26 | Pulmonary embolism | Prior HES records | 41270 | Boolean |
| I31 | Other diseases of pericardium | Prior HES records | 41270 | Boolean |
| I34 | Nonrheumatic mitral valve disorders | Prior HES records | 41270 | Boolean |
| I35 | Nonrheumatic aortic valve disorders | Prior HES records | 41270 | Boolean |
| I42 | Cardiomyopathy | Prior HES records | 41270 | Boolean |
| I44 | Atrioventricular and left bundle-branch block | Prior HES records | 41270 | Boolean |
| I45 | Other conduction disorders | Prior HES records | 41270 | Boolean |
| I47 | Paroxysmal tachycardia | Prior HES records | 41270 | Boolean |
| I48 | Atrial fibrillation and flutter | Prior HES records | 41270 | Boolean |
| I49 | Other cardiac arrhythmias | Prior HES records | 41270 | Boolean |
| I50 | Heart failure | Prior HES records | 41270 | Boolean |
| I51 | Complications and ill-defined descriptions of heart disease | Prior HES records | 41270 | Boolean |
| I60 | Subarachnoid haemorrhage | Prior HES records | 41270 | Boolean |
| I63 | Cerebral infarction | Prior HES records | 41270 | Boolean |
| I64 | Stroke, not specified as haemorrhage or infarction | Prior HES records | 41270 | Boolean |
| I67 | Other cerebrovascular diseases | Prior HES records | 41270 | Boolean |
| I69 | Sequelae of cerebrovascular disease | Prior HES records | 41270 | Boolean |
| I70 | Atherosclerosis | Prior HES records | 41270 | Boolean |
| I73 | Other peripheral vascular diseases | Prior HES records | 41270 | Boolean |
| I77 | Other disorders of arteries and arterioles | Prior HES records | 41270 | Boolean |
| I80 | Phlebitis and thrombophlebitis | Prior HES records | 41270 | Boolean |
| I83 | Varicose veins of lower extremities | Prior HES records | 41270 | Boolean |
| I84 | Haemorrhoids | Prior HES records | 41270 | Boolean |
| I95 | Hypotension | Prior HES records | 41270 | Boolean |
| J06 | Acute upper respiratory infections of multiple and unspecified sites | Prior HES records | 41270 | Boolean |
| J18 | Pneumonia, organism unspecified | Prior HES records | 41270 | Boolean |
| J22 | Unspecified acute lower respiratory infection | Prior HES records | 41270 | Boolean |
| J30 | Vasomotor and allergic rhinitis | Prior HES records | 41270 | Boolean |
| J31 | Chronic rhinitis, nasopharyngitis and pharyngitis | Prior HES records | 41270 | Boolean |
| J32 | Chronic sinusitis | Prior HES records | 41270 | Boolean |
| J33 | Nasal polyp | Prior HES records | 41270 | Boolean |
| J34 | Other disorders of nose and nasal sinuses | Prior HES records | 41270 | Boolean |
| J35 | Chronic diseases of tonsils and adenoids | Prior HES records | 41270 | Boolean |
| J38 | Diseases of vocal cords and larynx, not elsewhere classified | Prior HES records | 41270 | Boolean |
| J43 | Emphysema | Prior HES records | 41270 | Boolean |
| J44 | Other chronic obstructive pulmonary disease | Prior HES records | 41270 | Boolean |
| J45 | Asthma | Prior HES records | 41270 | Boolean |
| J47 | Bronchiectasis | Prior HES records | 41270 | Boolean |
| J90 | Pleural effusion, not elsewhere classified | Prior HES records | 41270 | Boolean |
| J98 | Other respiratory disorders | Prior HES records | 41270 | Boolean |
| K01 | Embedded and impacted teeth | Prior HES records | 41270 | Boolean |
| K02 | Dental caries | Prior HES records | 41270 | Boolean |
| K04 | Diseases of pulp and periapical tissues | Prior HES records | 41270 | Boolean |
| K05 | Gingivitis and periodontal diseases | Prior HES records | 41270 | Boolean |
| K08 | Other disorders of teeth and supporting structures | Prior HES records | 41270 | Boolean |
| K11 | Diseases of salivary glands | Prior HES records | 41270 | Boolean |
| K13 | Other diseases of lip and oral mucosa | Prior HES records | 41270 | Boolean |
| K14 | Diseases of tongue | Prior HES records | 41270 | Boolean |
| K20 | Oesophagitis | Prior HES records | 41270 | Boolean |
| K21 | Gastro-oesophageal reflux disease | Prior HES records | 41270 | Boolean |
| K22 | Other diseases of oesophagus | Prior HES records | 41270 | Boolean |
| K25 | Gastric ulcer | Prior HES records | 41270 | Boolean |
| K26 | Duodenal ulcer | Prior HES records | 41270 | Boolean |
| K29 | Gastritis and duodenitis | Prior HES records | 41270 | Boolean |
| K30 | Dyspepsia | Prior HES records | 41270 | Boolean |
| K31 | Other diseases of stomach and duodenum | Prior HES records | 41270 | Boolean |
| K35 | Acute appendicitis | Prior HES records | 41270 | Boolean |
| K40 | Inguinal hernia | Prior HES records | 41270 | Boolean |
| K42 | Umbilical hernia | Prior HES records | 41270 | Boolean |
| K43 | Ventral hernia | Prior HES records | 41270 | Boolean |
| K44 | Diaphragmatic hernia | Prior HES records | 41270 | Boolean |
| K50 | Crohn's disease [regional enteritis] | Prior HES records | 41270 | Boolean |
| K51 | Ulcerative colitis | Prior HES records | 41270 | Boolean |
| K52 | Other non-infective gastro-enteritis and colitis | Prior HES records | 41270 | Boolean |
| K56 | Paralytic ileus and intestinal obstruction without hernia | Prior HES records | 41270 | Boolean |
| K57 | Diverticular disease of intestine | Prior HES records | 41270 | Boolean |
| K58 | Irritable bowel syndrome | Prior HES records | 41270 | Boolean |
| K59 | Other functional intestinal disorders | Prior HES records | 41270 | Boolean |
| K60 | Fissure and fistula of anal and rectal regions | Prior HES records | 41270 | Boolean |
| K61 | Abscess of anal and rectal regions | Prior HES records | 41270 | Boolean |
| K62 | Other diseases of anus and rectum | Prior HES records | 41270 | Boolean |
| K63 | Other diseases of intestine | Prior HES records | 41270 | Boolean |
| K66 | Other disorders of peritoneum | Prior HES records | 41270 | Boolean |
| K70 | Alcoholic liver disease | Prior HES records | 41270 | Boolean |
| K76 | Other diseases of liver | Prior HES records | 41270 | Boolean |
| K80 | Cholelithiasis | Prior HES records | 41270 | Boolean |
| K81 | Cholecystitis | Prior HES records | 41270 | Boolean |
| K82 | Other diseases of gallbladder | Prior HES records | 41270 | Boolean |
| K83 | Other diseases of biliary tract | Prior HES records | 41270 | Boolean |
| K85 | Acute pancreatitis | Prior HES records | 41270 | Boolean |
| K86 | Other diseases of pancreas | Prior HES records | 41270 | Boolean |
| K90 | Intestinal malabsorption | Prior HES records | 41270 | Boolean |
| K91 | Postprocedural disorders of digestive system, not elsewhere classified | Prior HES records | 41270 | Boolean |
| K92 | Other diseases of digestive system | Prior HES records | 41270 | Boolean |
| L02 | Cutaneous abscess, furuncle and carbuncle | Prior HES records | 41270 | Boolean |
| L03 | Cellulitis | Prior HES records | 41270 | Boolean |
| L05 | Pilonidal cyst | Prior HES records | 41270 | Boolean |
| L08 | Other local infections of skin and subcutaneous tissue | Prior HES records | 41270 | Boolean |
| L29 | Pruritus | Prior HES records | 41270 | Boolean |
| L30 | Other dermatitis | Prior HES records | 41270 | Boolean |
| L40 | Psoriasis | Prior HES records | 41270 | Boolean |
| L57 | Skin changes due to chronic exposure to nonionising radiation | Prior HES records | 41270 | Boolean |
| L60 | Nail disorders | Prior HES records | 41270 | Boolean |
| L72 | Follicular cysts of skin and subcutaneous tissue | Prior HES records | 41270 | Boolean |
| L82 | Seborrhoeic keratosis | Prior HES records | 41270 | Boolean |
| L90 | Atrophic disorders of skin | Prior HES records | 41270 | Boolean |
| L91 | Hypertrophic disorders of skin | Prior HES records | 41270 | Boolean |
| L98 | Other disorders of skin and subcutaneous tissue, not elsewhere classified | Prior HES records | 41270 | Boolean |
| M06 | Other rheumatoid arthritis | Prior HES records | 41270 | Boolean |
| M10 | Gout | Prior HES records | 41270 | Boolean |
| M13 | Other arthritis | Prior HES records | 41270 | Boolean |
| M15 | Polyarthrosis | Prior HES records | 41270 | Boolean |
| M16 | Coxarthrosis [arthrosis of hip] | Prior HES records | 41270 | Boolean |
| M17 | Gonarthrosis [arthrosis of knee] | Prior HES records | 41270 | Boolean |
| M18 | Arthrosis of first carpometacarpal joint | Prior HES records | 41270 | Boolean |
| M19 | Other arthrosis | Prior HES records | 41270 | Boolean |
| M20 | Acquired deformities of fingers and toes | Prior HES records | 41270 | Boolean |
| M21 | Other acquired deformities of limbs | Prior HES records | 41270 | Boolean |
| M22 | Disorders of patella | Prior HES records | 41270 | Boolean |
| M23 | Internal derangement of knee | Prior HES records | 41270 | Boolean |
| M24 | Other specific joint derangements | Prior HES records | 41270 | Boolean |
| M25 | Other joint disorders, not elsewhere classified | Prior HES records | 41270 | Boolean |
| M35 | Other systemic involvement of connective tissue | Prior HES records | 41270 | Boolean |
| M43 | Other deforming dorsopathies | Prior HES records | 41270 | Boolean |
| M47 | Spondylosis | Prior HES records | 41270 | Boolean |
| M48 | Other spondylopathies | Prior HES records | 41270 | Boolean |
| M50 | Cervical disk disorders | Prior HES records | 41270 | Boolean |
| M51 | Other intervertebral disk disorders | Prior HES records | 41270 | Boolean |
| M54 | Dorsalgia | Prior HES records | 41270 | Boolean |
| M65 | Synovitis and tenosynovitis | Prior HES records | 41270 | Boolean |
| M67 | Other disorders of synovium and tendon | Prior HES records | 41270 | Boolean |
| M70 | Soft tissue disorders related to use, overuse and pressure | Prior HES records | 41270 | Boolean |
| M72 | Fibroblastic disorders | Prior HES records | 41270 | Boolean |
| M75 | Shoulder lesions | Prior HES records | 41270 | Boolean |
| M77 | Other enthesopathies | Prior HES records | 41270 | Boolean |
| M79 | Other soft tissue disorders, not elsewhere classified | Prior HES records | 41270 | Boolean |
| M80 | Osteoporosis with pathological fracture | Prior HES records | 41270 | Boolean |
| M81 | Osteoporosis without pathological fracture | Prior HES records | 41270 | Boolean |
| M84 | Disorders of continuity of bone | Prior HES records | 41270 | Boolean |
| M89 | Other disorders of bone | Prior HES records | 41270 | Boolean |
| N12 | Tubulo-interstitial nephritis, not specified as acute or chronic | Prior HES records | 41270 | Boolean |
| N13 | Obstructive and reflux uropathy | Prior HES records | 41270 | Boolean |
| N17 | Acute renal failure | Prior HES records | 41270 | Boolean |
| N18 | Chronic renal failure | Prior HES records | 41270 | Boolean |
| N19 | Unspecified renal failure | Prior HES records | 41270 | Boolean |
| N20 | Calculus of kidney and ureter | Prior HES records | 41270 | Boolean |
| N23 | Unspecified renal colic | Prior HES records | 41270 | Boolean |
| N28 | Other disorders of kidney and ureter, not elsewhere classified | Prior HES records | 41270 | Boolean |
| N30 | Cystitis | Prior HES records | 41270 | Boolean |
| N31 | Neuromuscular dysfunction of bladder, not elsewhere classified | Prior HES records | 41270 | Boolean |
| N32 | Other disorders of bladder | Prior HES records | 41270 | Boolean |
| N35 | Urethral stricture | Prior HES records | 41270 | Boolean |
| N39 | Other disorders of urinary system | Prior HES records | 41270 | Boolean |
| N40 | Hyperplasia of prostate | Prior HES records | 41270 | Boolean |
| N41 | Inflammatory diseases of prostate | Prior HES records | 41270 | Boolean |
| N42 | Other disorders of prostate | Prior HES records | 41270 | Boolean |
| N43 | Hydrocele and spermatocele | Prior HES records | 41270 | Boolean |
| N45 | Orchitis and epididymitis | Prior HES records | 41270 | Boolean |
| N47 | Redundant prepuce, phimosis and paraphimosis | Prior HES records | 41270 | Boolean |
| N48 | Other disorders of penis | Prior HES records | 41270 | Boolean |
| N50 | Other disorders of male genital organs | Prior HES records | 41270 | Boolean |
| N60 | Benign mammary dysplasia | Prior HES records | 41270 | Boolean |
| N62 | Hypertrophy of breast | Prior HES records | 41270 | Boolean |
| N63 | Unspecified lump in breast | Prior HES records | 41270 | Boolean |
| N64 | Other disorders of breast | Prior HES records | 41270 | Boolean |
| N70 | Salpingitis and oophoritis | Prior HES records | 41270 | Boolean |
| N72 | Inflammatory disease of cervix uteri | Prior HES records | 41270 | Boolean |
| N73 | Other female pelvic inflammatory diseases | Prior HES records | 41270 | Boolean |
| N75 | Diseases of Bartholin's gland | Prior HES records | 41270 | Boolean |
| N76 | Other inflammation of vagina and vulva | Prior HES records | 41270 | Boolean |
| N80 | Endometriosis | Prior HES records | 41270 | Boolean |
| N81 | Female genital prolapse | Prior HES records | 41270 | Boolean |
| N83 | Noninflammatory disorders of ovary, Fallopian tube and broad ligament | Prior HES records | 41270 | Boolean |
| N84 | Polyp of female genital tract | Prior HES records | 41270 | Boolean |
| N85 | Other noninflammatory disorders of uterus, except cervix | Prior HES records | 41270 | Boolean |
| N86 | Erosion and ectropion of cervix uteri | Prior HES records | 41270 | Boolean |
| N87 | Dysplasia of cervix uteri | Prior HES records | 41270 | Boolean |
| N88 | Other noninflammatory disorders of cervix uteri | Prior HES records | 41270 | Boolean |
| N89 | Other noninflammatory disorders of vagina | Prior HES records | 41270 | Boolean |
| N90 | Other noninflammatory disorders of vulva and perineum | Prior HES records | 41270 | Boolean |
| N92 | Excessive, frequent and irregular menstruation | Prior HES records | 41270 | Boolean |
| N93 | Other abnormal uterine and vaginal bleeding | Prior HES records | 41270 | Boolean |
| N94 | Pain and other conditions associated with female genital organs and menstrual cycle | Prior HES records | 41270 | Boolean |
| N95 | Menopausal and other perimenopausal disorders | Prior HES records | 41270 | Boolean |
| N97 | Female infertility | Prior HES records | 41270 | Boolean |
| N99 | Postprocedural disorders of genito-urinary system, not elsewhere classified | Prior HES records | 41270 | Boolean |
| O02 | Other abnormal products of conception | Prior HES records | 41270 | Boolean |
| O03 | Spontaneous abortion | Prior HES records | 41270 | Boolean |
| O04 | Medical abortion | Prior HES records | 41270 | Boolean |
| O13 | Gestational [pregnancy-induced] hypertension without significant proteinuria | Prior HES records | 41270 | Boolean |
| O16 | Unspecified maternal hypertension | Prior HES records | 41270 | Boolean |
| O20 | Haemorrhage in early pregnancy | Prior HES records | 41270 | Boolean |
| O26 | Maternal care for other conditions predominantly related to pregnancy | Prior HES records | 41270 | Boolean |
| O32 | Maternal care for known or suspected malpresentation of foetus | Prior HES records | 41270 | Boolean |
| O34 | Maternal care for known or suspected abnormality of pelvic organs | Prior HES records | 41270 | Boolean |
| O36 | Maternal care for other known or suspected foetal problems | Prior HES records | 41270 | Boolean |
| O42 | Premature rupture of membranes | Prior HES records | 41270 | Boolean |
| O46 | Antepartum haemorrhage, not elsewhere classified | Prior HES records | 41270 | Boolean |
| O47 | False labour | Prior HES records | 41270 | Boolean |
| O48 | Prolonged pregnancy | Prior HES records | 41270 | Boolean |
| O60 | Preterm delivery | Prior HES records | 41270 | Boolean |
| O62 | Abnormalities of forces of labour | Prior HES records | 41270 | Boolean |
| O63 | Long labour | Prior HES records | 41270 | Boolean |
| O64 | Obstructed labour due to malposition and malpresentation of foetus | Prior HES records | 41270 | Boolean |
| O68 | Labour and delivery complicated by foetal stress [distress] | Prior HES records | 41270 | Boolean |
| O69 | Labour and delivery complicated by umbilical cord complications | Prior HES records | 41270 | Boolean |
| O70 | Perineal laceration during delivery | Prior HES records | 41270 | Boolean |
| O72 | Postpartum haemorrhage | Prior HES records | 41270 | Boolean |
| O75 | Other complications of labour and delivery, not elsewhere classified | Prior HES records | 41270 | Boolean |
| O80 | Single spontaneous delivery | Prior HES records | 41270 | Boolean |
| O82 | Single delivery by Caesarean section | Prior HES records | 41270 | Boolean |
| O99 | Other maternal diseases classifiable elsewhere but complicating pregnancy, childbirth and the puerperium | Prior HES records | 41270 | Boolean |
| R00 | Abnormalities of heart beat | Prior HES records | 41270 | Boolean |
| R03 | Abnormal blood-pressure reading, without diagnosis | Prior HES records | 41270 | Boolean |
| R04 | Haemorrhage from respiratory passages | Prior HES records | 41270 | Boolean |
| R05 | Cough | Prior HES records | 41270 | Boolean |
| R06 | Abnormalities of breathing | Prior HES records | 41270 | Boolean |
| R07 | Pain in throat and chest | Prior HES records | 41270 | Boolean |
| R09 | Other symptoms and signs involving the circulatory and respiratory systems | Prior HES records | 41270 | Boolean |
| R10 | Abdominal and pelvic pain | Prior HES records | 41270 | Boolean |
| R11 | Nausea and vomiting | Prior HES records | 41270 | Boolean |
| R12 | Heartburn | Prior HES records | 41270 | Boolean |
| R13 | Dysphagia | Prior HES records | 41270 | Boolean |
| R14 | Flatulence and related conditions | Prior HES records | 41270 | Boolean |
| R15 | Faecal incontinence | Prior HES records | 41270 | Boolean |
| R19 | Other symptoms and signs involving the digestive system and abdomen | Prior HES records | 41270 | Boolean |
| R20 | Disturbances of skin sensation | Prior HES records | 41270 | Boolean |
| R21 | Rash and other nonspecific skin eruption | Prior HES records | 41270 | Boolean |
| R22 | Localised swelling, mass and lump of skin and subcutaneous tissue | Prior HES records | 41270 | Boolean |
| R26 | Abnormalities of gait and mobility | Prior HES records | 41270 | Boolean |
| R29 | Other symptoms and signs involving the nervous and musculoskeletal systems | Prior HES records | 41270 | Boolean |
| R30 | Pain associated with micturition | Prior HES records | 41270 | Boolean |
| R31 | Unspecified haematuria | Prior HES records | 41270 | Boolean |
| R32 | Unspecified urinary incontinence | Prior HES records | 41270 | Boolean |
| R33 | Retention of urine | Prior HES records | 41270 | Boolean |
| R35 | Polyuria | Prior HES records | 41270 | Boolean |
| R39 | Other symptoms and signs involving the urinary system | Prior HES records | 41270 | Boolean |
| R41 | Other symptoms and signs involving cognitive functions and awareness | Prior HES records | 41270 | Boolean |
| R42 | Dizziness and giddiness | Prior HES records | 41270 | Boolean |
| R47 | Speech disturbances, not elsewhere classified | Prior HES records | 41270 | Boolean |
| R49 | Voice disturbances | Prior HES records | 41270 | Boolean |
| R50 | Fever of unknown origin | Prior HES records | 41270 | Boolean |
| R51 | Headache | Prior HES records | 41270 | Boolean |
| R52 | Pain, not elsewhere classified | Prior HES records | 41270 | Boolean |
| R53 | Malaise and fatigue | Prior HES records | 41270 | Boolean |
| R55 | Syncope and collapse | Prior HES records | 41270 | Boolean |
| R56 | Convulsions, not elsewhere classified | Prior HES records | 41270 | Boolean |
| R59 | Enlarged lymph nodes | Prior HES records | 41270 | Boolean |
| R63 | Symptoms and signs concerning food and fluid intake | Prior HES records | 41270 | Boolean |
| R69 | Unknown and unspecified causes of morbidity | Prior HES records | 41270 | Boolean |
| R79 | Other abnormal findings of blood chemistry | Prior HES records | 41270 | Boolean |
| R87 | Abnormal findings in specimens from female genital organs | Prior HES records | 41270 | Boolean |
| R91 | Abnormal findings on diagnostic imaging of lung | Prior HES records | 41270 | Boolean |
| R93 | Abnormal findings on diagnostic imaging of other body structures | Prior HES records | 41270 | Boolean |
| R94 | Abnormal results of function studies | Prior HES records | 41270 | Boolean |
| S00 | Superficial injury of head | Prior HES records | 41270 | Boolean |
| S01 | Open wound of head | Prior HES records | 41270 | Boolean |
| S02 | Fracture of skull and facial bones | Prior HES records | 41270 | Boolean |
| S06 | Intracranial injury | Prior HES records | 41270 | Boolean |
| S09 | Other and unspecified injuries of head | Prior HES records | 41270 | Boolean |
| S22 | Fracture of rib(s), sternum and thoracic spine | Prior HES records | 41270 | Boolean |
| S32 | Fracture of lumbar spine and pelvis | Prior HES records | 41270 | Boolean |
| S42 | Fracture of shoulder and upper arm | Prior HES records | 41270 | Boolean |
| S52 | Fracture of forearm | Prior HES records | 41270 | Boolean |
| S61 | Open wound of wrist and hand | Prior HES records | 41270 | Boolean |
| S62 | Fracture at wrist and hand level | Prior HES records | 41270 | Boolean |
| S64 | Injury of nerves at wrist and hand level | Prior HES records | 41270 | Boolean |
| S66 | Injury of muscle and tendon at wrist and hand level | Prior HES records | 41270 | Boolean |
| S72 | Fracture of femur | Prior HES records | 41270 | Boolean |
| S82 | Fracture of lower leg, including ankle | Prior HES records | 41270 | Boolean |
| S83 | Dislocation, sprain and strain of joints and ligaments of knee | Prior HES records | 41270 | Boolean |
| S86 | Injury of muscle and tendon at lower leg level | Prior HES records | 41270 | Boolean |
| S92 | Fracture of foot, except ankle | Prior HES records | 41270 | Boolean |
| T39 | Poisoning by nonopioid analgesics, antipyretics and antirheumatics | Prior HES records | 41270 | Boolean |
| T42 | Poisoning by antiepileptic, sedative-hypnotic and anti-Parkinsonism drugs | Prior HES records | 41270 | Boolean |
| T43 | Poisoning by psychotropic drugs, not elsewhere classified | Prior HES records | 41270 | Boolean |
| T51 | Toxic effect of alcohol | Prior HES records | 41270 | Boolean |
| T78 | Adverse effects, not elsewhere classified | Prior HES records | 41270 | Boolean |
| T81 | Complications of procedures, not elsewhere classified | Prior HES records | 41270 | Boolean |
| T82 | Complications of cardiac and vascular prosthetic devices, implants and grafts | Prior HES records | 41270 | Boolean |
| T83 | Complications of genito-urinary prosthetic devices, implants and grafts | Prior HES records | 41270 | Boolean |
| T84 | Complications of internal orthopaedic prosthetic devices, implants and grafts | Prior HES records | 41270 | Boolean |
| T85 | Complications of other internal prosthetic devices, implants and grafts | Prior HES records | 41270 | Boolean |
| T88 | Other complications of surgical and medical care, not elsewhere classified | Prior HES records | 41270 | Boolean |
| T93 | Sequelae of injuries of lower limb | Prior HES records | 41270 | Boolean |
| V18 | Pedal cyclist injured in noncollision transport accident | Prior HES records | 41270 | Boolean |
| V43 | Car occupant injured in collision with car, pick-up truck or van | Prior HES records | 41270 | Boolean |
| W01 | Fall on same level from slipping, tripping and stumbling | Prior HES records | 41270 | Boolean |
| W10 | Fall on and from stairs and steps | Prior HES records | 41270 | Boolean |
| W11 | Fall on and from ladder | Prior HES records | 41270 | Boolean |
| W18 | Other fall on same level | Prior HES records | 41270 | Boolean |
| W19 | Unspecified fall | Prior HES records | 41270 | Boolean |
| W22 | Striking against or struck by other objects | Prior HES records | 41270 | Boolean |
| W23 | Caught, crushed, jammed or pinched in or between objects | Prior HES records | 41270 | Boolean |
| X50 | Overexertion and strenuous or repetitive movements | Prior HES records | 41270 | Boolean |
| X59 | Exposure to unspecified factor | Prior HES records | 41270 | Boolean |
| X60 | Intentional self-poisoning by and exposure to nonopioid analgesics, antipyretics and antirheumatics | Prior HES records | 41270 | Boolean |
| X61 | Intentional self-poisoning by and exposure to antiepileptic, sedative-hypnotic, anti-Parkinsonism and psychotropic drugs, not elsewhere classified | Prior HES records | 41270 | Boolean |
| X65 | Intentional self-poisoning by and exposure to alcohol | Prior HES records | 41270 | Boolean |
| Y04 | Assault by bodily force | Prior HES records | 41270 | Boolean |
| Y42 | Hormones and their synthetic substitutes and antagonists, not elsewhere classified | Prior HES records | 41270 | Boolean |
| Y43 | Primarily systemic agents | Prior HES records | 41270 | Boolean |
| Y83 | Surgical operation and other surgical procedures as the cause of abnormal reaction of the patient, or of later complication, without mention of misadventure at the time of the procedure | Prior HES records | 41270 | Boolean |
| Y84 | Other medical procedures as the cause of abnormal reaction of the patient, or of later complication, without mention of misadventure at the time of the procedure | Prior HES records | 41270 | Boolean |
| Y86 | Sequelae of other accidents | Prior HES records | 41270 | Boolean |
| Z01 | Other special examinations and investigations of persons without complaint or reported diagnosis | Prior HES records | 41270 | Boolean |
| Z03 | Medical observation and evaluation for suspected diseases and conditions | Prior HES records | 41270 | Boolean |
| Z04 | Examination and observation for other reasons | Prior HES records | 41270 | Boolean |
| Z08 | Follow-up examination after treatment for malignant neoplasm | Prior HES records | 41270 | Boolean |
| Z09 | Follow-up examination after treatment for conditions other than malignant neoplasms | Prior HES records | 41270 | Boolean |
| Z11 | Special screening examination for infectious and parasitic diseases | Prior HES records | 41270 | Boolean |
| Z12 | Special screening examination for neoplasms | Prior HES records | 41270 | Boolean |
| Z13 | Special screening examination for other diseases and disorders | Prior HES records | 41270 | Boolean |
| Z30 | Contraceptive management | Prior HES records | 41270 | Boolean |
| Z33 | Pregnant state, incidental | Prior HES records | 41270 | Boolean |
| Z34 | Supervision of normal pregnancy | Prior HES records | 41270 | Boolean |
| Z35 | Supervision of high-risk pregnancy | Prior HES records | 41270 | Boolean |
| Z36 | Antenatal screening | Prior HES records | 41270 | Boolean |
| Z37 | Outcome of delivery | Prior HES records | 41270 | Boolean |
| Z39 | Postpartum care and examination | Prior HES records | 41270 | Boolean |
| Z41 | Procedures for purposes other than remedying health state | Prior HES records | 41270 | Boolean |
| Z42 | Follow-up care involving plastic surgery | Prior HES records | 41270 | Boolean |
| Z43 | Attention to artificial openings | Prior HES records | 41270 | Boolean |
| Z45 | Adjustment and management of implanted device | Prior HES records | 41270 | Boolean |
| Z46 | Fitting and adjustment of other devices | Prior HES records | 41270 | Boolean |
| Z47 | Other orthopaedic follow-up care | Prior HES records | 41270 | Boolean |
| Z48 | Other surgical follow-up care | Prior HES records | 41270 | Boolean |
| Z50 | Care involving use of rehabilitation procedures | Prior HES records | 41270 | Boolean |
| Z51 | Other medical care | Prior HES records | 41270 | Boolean |
| Z53 | Persons encountering health services for specific procedures, not carried out | Prior HES records | 41270 | Boolean |
| Z60 | Problems related to social environment | Prior HES records | 41270 | Boolean |
| Z71 | Persons encountering health services for other counselling and medical advice, not elsewhere classified | Prior HES records | 41270 | Boolean |
| Z72 | Problems related to lifestyle | Prior HES records | 41270 | Boolean |
| Z80 | Family history of malignant neoplasm | Prior HES records | 41270 | Boolean |
| Z82 | Family history of certain disabilities and chronic diseases leading to disablement | Prior HES records | 41270 | Boolean |
| Z83 | Family history of other specific disorders | Prior HES records | 41270 | Boolean |
| Z85 | Personal history of malignant neoplasm | Prior HES records | 41270 | Boolean |
| Z86 | Personal history of certain other diseases | Prior HES records | 41270 | Boolean |
| Z87 | Personal history of other diseases and conditions | Prior HES records | 41270 | Boolean |
| Z88 | Personal history of allergy to drugs, medicaments and biological substances | Prior HES records | 41270 | Boolean |
| Z90 | Acquired absence of organs, not elsewhere classified | Prior HES records | 41270 | Boolean |
| Z91 | Personal history of risk-factors, not elsewhere classified | Prior HES records | 41270 | Boolean |
| Z92 | Personal history of medical treatment | Prior HES records | 41270 | Boolean |
| Z93 | Artificial opening status | Prior HES records | 41270 | Boolean |
| Z94 | Transplanted organ and tissue status | Prior HES records | 41270 | Boolean |
| Z95 | Presence of cardiac and vascular implants and grafts | Prior HES records | 41270 | Boolean |
| Z96 | Presence of other functional implants | Prior HES records | 41270 | Boolean |
| Z97 | Presence of other devices | Prior HES records | 41270 | Boolean |
| Z98 | Other postsurgical states | Prior HES records | 41270 | Boolean |
| Z99 | Dependence on enabling machines and devices, not elsewhere classified | Prior HES records | 41270 | Boolean |

**Supplementary Table 11: Predictors: Past Medical History:** The table details each predictor along with its measurement unit, coding scheme, data source, corresponding UKB Field ID, and any transformation applied prior to modelling. All transformations were estimated within the training fold and then applied to the held-out data to avoid information leakage.

**Provided as separate Microsoft Excel file; Supplementary Table 12: Predictors: Self-reported information predictors:** The table details each predictor along with its measurement unit, coding scheme, data source, corresponding UKB Field ID, and any transformation applied prior to modelling. All transformations were estimated within the training fold and then applied to the held-out data to avoid information leakage. Due to its size, the table is provided as a separate Microsoft Excel file.

| **Predictor** | **Response options/unit** | **UKB Field ID** | **Transformation** |
| --- | --- | --- | --- |
| Sex | Male \| Female | 31 | Boolean |
| Age at recruitment | years | 21022 | Log-standardscaling (mean=0, std=1) |
| Current smoking status | No \| Yes | 20116 | Boolean |
| Mean SBP (if more than one measurement) | mmHg | 4080, 93 | Log-standardscaling (mean=0, std=1) |
| Total cholesterol | mg/dL | 30690 | Log-standardscaling (mean=0, std=1) |
| HDL cholesterol | mg/dL | 30760 | Log-standardscaling (mean=0, std=1) |
| Risk region (NOT USED - everyone in cohort from same region - UK) | UK (Low risk) |  |  |

**Supplementary Table 13: Predictors: SCORE-2:** The table details each predictor along with its measurement unit, coding scheme, corresponding UKB Field ID, and any transformation applied prior to modelling. All transformations were estimated within the training fold and then applied to the held-out data to avoid information leakage.

| **Predictor** | **Data-coding** | **Response options/unit** | **UKB Field ID** | **Transformation** |
| --- | --- | --- | --- | --- |
| Sex |  | Male \| Female | 31 | Boolean |
| Age at recruitment |  | years | 21022 | Log-standardscaling (mean=0, std=1) |
| Total cholesterol |  | mg/dL | 30690 | Log-standardscaling (mean=0, std=1) |
| HDL cholesterol |  | mg/dL | 30760 | Log-standardscaling (mean=0, std=1) |
| Mean SBP (if more than one measurement) |  | mmHg | 4080, 93 | Log-standardscaling (mean=0, std=1) |
| Diabetes status (both type 1 and type 2) | ICD-10: E10, E11, E12, E13, E14; ICD-9: 250; Non-cancer illness codes: 1220, 1222, 1223 | No \| Yes | 41270, 42171, 20002, 40001, 40002 | Boolean |
| Current smoker |  | No \| Yes | 20116 | Boolean |
| eGFR |  | mL/min/1.73 m² | 30700, 31, 21022 | CKD-EPI 2021  Log-standardscaling (mean=0, std=1) |
| Using anti-hypertensive medication | 1140860332, 1140860334, 1140860336, 1140860338, 1140860340, 1140860342, 1140860348, 1140860352, 1140860356, 1140860358, 1140860362, 1140860380, 1140860390, 1140860394, 1140860396, 1140860398, 1140860402, 1140860410, 1140860418, 1140860422, 1140860426, 1140860434, 1140860478, 1140860492, 1140860498, 1140860520, 1140860532, 1140860552, 1140860558, 1140860562, 1140860564, 1140860580, 1140860628, 1140860632, 1140860638, 1140860654, 1140860658, 1140860706, 1140860714, 1140860728, 1140860736, 1140860738, 1140860758, 1140860764, 1140860776, 1140860784, 1140860790, 1140860828, 1140860830, 1140860834, 1140860836, 1140860838, 1140860846, 1140860848, 1140860862, 1140860878, 1140860882, 1140860912, 1140860918, 1140860938, 1140860942, 1140860952, 1140860972, 1140860976, 1140860982, 1140860988, 1140860994, 1140861008, 1140861010, 1140861016, 1140861022, 1140861024, 1140861068, 1140861070, 1140861088, 1140861090, 1140861106, 1140861120, 1140861128, 1140861130, 1140861136, 1140861138, 1140861190, 1140861194, 1140861202, 1140861266, 1140861268, 1140861326, 1140861384, 1140864950, 1140864952, 1140866072, 1140866084, 1140866086, 1140866090, 1140866092, 1140866094, 1140866104, 1140866108, 1140866110, 1140866116, 1140866122, 1140866136, 1140866138, 1140866140, 1140866144, 1140866146, 1140866162, 1140866164, 1140866168, 1140866182, 1140866192, 1140866202, 1140866206, 1140866210, 1140866212, 1140866220, 1140866230, 1140866232, 1140866236, 1140866244, 1140866248, 1140866282, 1140866306, 1140866308, 1140866312, 1140866318, 1140866330, 1140866332, 1140866334, 1140866340, 1140866352, 1140866360, 1140866388, 1140866390, 1140866396, 1140866400, 1140866406, 1140866408, 1140866410, 1140866412, 1140866416, 1140866422, 1140866426, 1140866438, 1140866440, 1140866442, 1140866448, 1140866450, 1140866460, 1140866466, 1140866484, 1140866554, 1140866692, 1140866704, 1140866712, 1140866724, 1140866756, 1140866758, 1140866764, 1140866766, 1140866778, 1140866798, 1140866800, 1140866802, 1140866804, 1140875808, 1140879762, 1140879778, 1140879782, 1140879786, 1140879794, 1140879806, 1140879810, 1140879818, 1140879822, 1140879824, 1140879834, 1140879842, 1140879854, 1140879866, 1140888510, 1140888556, 1140888560, 1140888578, 1140888582, 1140888586, 1140888760, 1140888762, 1140909368, 1140911698, 1140916356, 1140923572, 1140923712, 1140923718, 1140926778, 1140926780, 1141145668, 1141151016, 1141151018, 1141151382, 1141152600, 1141153026, 1141153032, 1141153328, 1141156754, 1141156808, 1141157252, 1141157254, 1141164148, 1141164154, 1141164276, 1141165476, 1141166006, 1141167822, 1141167832, 1141171152, 1141172682, 1141172686, 1141172698, 1141173888, 1141180592, 1141187790, 1141190160, 1141192064, 1141193282, 1141193346, 1141194804, 1141194808, 1141194810, 1141201038, 1141201040, 1140860382, 1140860386, 1140860404, 1140860406, 1140860454, 1140860470, 1140860534, 1140860544, 1140860590, 1140860610, 1140860690, 1140860696, 1140860750, 1140860752, 1140860802, 1140860806, 1140860840, 1140860842, 1140860892, 1140860904, 1140860954, 1140860966, 1140861000, 1140861002, 1140861034, 1140861046, 1140861110, 1140861114, 1140861166, 1140861176, 1140861276, 1140861282, 1140866074, 1140866078, 1140866096, 1140866102, 1140866128, 1140866132, 1140866156, 1140866158, 1140866194, 1140866200, 1140866222, 1140866226, 1140866262, 1140866280, 1140866324, 1140866328, 1140866354, 1140866356, 1140866402, 1140866404, 1140866418, 1140866420, 1140866444, 1140866446, 1140866506, 1140866546, 1140866726, 1140866738, 1140866782, 1140866784, 1140879758, 1140879760, 1140879798, 1140879802, 1140879826, 1140879830, 1140888512, 1140888552, 1140888646, 1140888686, 1140916362, 1140917428, 1141145658, 1141145660, 1141152998, 1141153006, 1141156836, 1141156846, 1141164280, 1141165470, 1141171336, 1141171344, 1141180598, 1141187788, 1141194794, 1141194800 | No \| Yes | 20003, 6177, 6153 | Boolean |
| Using lipid-lowering medications | 1141146234, 1140888594, 1140888648, 1141192410, 1140861958, 1140865576, 1140865576, 1140861936, 1140861944, 1140862028, 1141175908, 1140851880, 1140851882, 1140861324, 1140861868, 1140861892, 1141162544, 1141192736, 1141192740, 1141157416, 1140861924, 1141157260, 1140861926, 1140861928, 1140861922, 1140861942, 1140861946, 1140861954, 1140862026, 1141168568, 1141171548, 1141201306, 1140888590, 1140861848, 1140861856, 1141157262, 1140861858, 1140926582, 1140861866, 1141188546, 1140861876, 1140861878, 1140861884, 1141181868, 1141172214, 1141182910, 1140865752, 1141157494, 1141145830 | No \| Yes | 20003, 6177, 6153 | Boolean |
| BMI |  | kg/m² | 21001 | Log-standardscaling (mean=0, std=1) |
| uACR |  | mg/g | 30500, 30510 | Log-standardscaling (mean=0, std=1) |
| HbA1c |  | mmol/mol | 30750 | Log-standardscaling (mean=0, std=1) |
| Townsend deprivation index at recruitment |  | score | 22189 |  |

**Supplementary Table 14: Predictors: PREVENT:** The table details each predictor along with its measurement unit, coding scheme, data source, corresponding UKB Field ID, and any transformation applied prior to modelling. All transformations were estimated within the training fold and then applied to the held-out data to avoid information leakage.

| **Predictor** | **Data coding** | **Response options/unit** | **Field ID** | **Transformation** |
| --- | --- | --- | --- | --- |
| Age at recruitment |  | years | 21022 | Log-standardscaling (mean=0, std=1) |
| Sex |  | Male \| Female | 31 | Boolean |
| Ethnicity |  | White or not stated \| Indian \| Pakistani \| Bangladeshi \| Other Asian \| Black Caribbean \| Black African \| Chinese \| Other ethnicity | 21000 | One-Hot-Encoding |
| Townsend deprivation index at recruitment |  | score | 22189 |  |
| Smoking status |  | Non-smoker \| Ex-smoker \| Light smoker \| Moderate smoker \| Heavy smoker | 20116 | One-Hot-Encoding |
| Diabetes status | Type 1 Diabetes  ICD10: E10,O240  ICD9: 25001, 25011, 25021, 25031, 25041, 25051, 25061, 25071, 25081, 25091, 25003, 25013, 25023, 25033, 25043, 25053, 25063, 25073, 25083, 25093  NCIC: 1222   Type 2 Diabetes  ICD10: E11, O241  ICD9: 25000, 25010, 25020, 25030, 25040, 25050, 25060, 25070, 25080, 25090, 25002, 25012, 25022, 25032, 25042, 25052, 25062, 25072, 25082, 25092  NCIC: 1220, 1223 | None \| Type 1 \| Type 2 | 41270, 42171, 20003, 40001, 40002 | Boolean |
| Heart disease in father/mother/siblings |  | No \| Yes | 20107, 20110, 20111 | Boolean |
| Chronic kidney disease | ICD10: N183, N184, N185  NCIC: 1192, 1519, 1609 | No \| Yes | 41270, 42171, 20003, 40001, 40002 | Boolean |
| Atrial fibrillation | ICD10: I48, I480, I481, I482, I483, I484, I489  OPCS4: K571, K621, K622, K623, K624  NCIC: 1471, 1483  OPC: 1524 | No \| Yes | 41270, 42171, 20003, 40001, 40002 | Boolean |
| On anti-hypertensives? | 1140860332, 1140860334, 1140860336, 1140860338, 1140860340, 1140860342, 1140860348, 1140860352, 1140860356, 1140860358, 1140860362, 1140860380, 1140860390, 1140860394, 1140860396, 1140860398, 1140860402, 1140860410, 1140860418, 1140860422, 1140860426, 1140860434, 1140860478, 1140860492, 1140860498, 1140860520, 1140860532, 1140860552, 1140860558, 1140860562, 1140860564, 1140860580, 1140860628, 1140860632, 1140860638, 1140860654, 1140860658, 1140860706, 1140860714, 1140860728, 1140860736, 1140860738, 1140860758, 1140860764, 1140860776, 1140860784, 1140860790, 1140860828, 1140860830, 1140860834, 1140860836, 1140860838, 1140860846, 1140860848, 1140860862, 1140860878, 1140860882, 1140860912, 1140860918, 1140860938, 1140860942, 1140860952, 1140860972, 1140860976, 1140860982, 1140860988, 1140860994, 1140861008, 1140861010, 1140861016, 1140861022, 1140861024, 1140861068, 1140861070, 1140861088, 1140861090, 1140861106, 1140861120, 1140861128, 1140861130, 1140861136, 1140861138, 1140861190, 1140861194, 1140861202, 1140861266, 1140861268, 1140861326, 1140861384, 1140864950, 1140864952, 1140866072, 1140866084, 1140866086, 1140866090, 1140866092, 1140866094, 1140866104, 1140866108, 1140866110, 1140866116, 1140866122, 1140866136, 1140866138, 1140866140, 1140866144, 1140866146, 1140866162, 1140866164, 1140866168, 1140866182, 1140866192, 1140866202, 1140866206, 1140866210, 1140866212, 1140866220, 1140866230, 1140866232, 1140866236, 1140866244, 1140866248, 1140866282, 1140866306, 1140866308, 1140866312, 1140866318, 1140866330, 1140866332, 1140866334, 1140866340, 1140866352, 1140866360, 1140866388, 1140866390, 1140866396, 1140866400, 1140866406, 1140866408, 1140866410, 1140866412, 1140866416, 1140866422, 1140866426, 1140866438, 1140866440, 1140866442, 1140866448, 1140866450, 1140866460, 1140866466, 1140866484, 1140866554, 1140866692, 1140866704, 1140866712, 1140866724, 1140866756, 1140866758, 1140866764, 1140866766, 1140866778, 1140866798, 1140866800, 1140866802, 1140866804, 1140875808, 1140879762, 1140879778, 1140879782, 1140879786, 1140879794, 1140879806, 1140879810, 1140879818, 1140879822, 1140879824, 1140879834, 1140879842, 1140879854, 1140879866, 1140888510, 1140888556, 1140888560, 1140888578, 1140888582, 1140888586, 1140888760, 1140888762, 1140909368, 1140911698, 1140916356, 1140923572, 1140923712, 1140923718, 1140926778, 1140926780, 1141145668, 1141151016, 1141151018, 1141151382, 1141152600, 1141153026, 1141153032, 1141153328, 1141156754, 1141156808, 1141157252, 1141157254, 1141164148, 1141164154, 1141164276, 1141165476, 1141166006, 1141167822, 1141167832, 1141171152, 1141172682, 1141172686, 1141172698, 1141173888, 1141180592, 1141187790, 1141190160, 1141192064, 1141193282, 1141193346, 1141194804, 1141194808, 1141194810, 1141201038, 1141201040, 1140860382, 1140860386, 1140860404, 1140860406, 1140860454, 1140860470, 1140860534, 1140860544, 1140860590, 1140860610, 1140860690, 1140860696, 1140860750, 1140860752, 1140860802, 1140860806, 1140860840, 1140860842, 1140860892, 1140860904, 1140860954, 1140860966, 1140861000, 1140861002, 1140861034, 1140861046, 1140861110, 1140861114, 1140861166, 1140861176, 1140861276, 1140861282, 1140866074, 1140866078, 1140866096, 1140866102, 1140866128, 1140866132, 1140866156, 1140866158, 1140866194, 1140866200, 1140866222, 1140866226, 1140866262, 1140866280, 1140866324, 1140866328, 1140866354, 1140866356, 1140866402, 1140866404, 1140866418, 1140866420, 1140866444, 1140866446, 1140866506, 1140866546, 1140866726, 1140866738, 1140866782, 1140866784, 1140879758, 1140879760, 1140879798, 1140879802, 1140879826, 1140879830, 1140888512, 1140888552, 1140888646, 1140888686, 1140916362, 1140917428, 1141145658, 1141145660, 1141152998, 1141153006, 1141156836, 1141156846, 1141164280, 1141165470, 1141171336, 1141171344, 1141180598, 1141187788, 1141194794, 1141194800 | No \| Yes | 20003, 6177, 6153 | Boolean |
| Migraines? | ICD10: G43, G440, N943  ICD9: 3461  NCIC: 1265 | No \| Yes | 41270, 42171, 20003, 40001, 40002 | Boolean |
| Rheumatoid arthritis? | ICD10: M05, M06  ICD9: 7141, 7142, 7149  NCIC: 1464 | No \| Yes | 41270, 42171, 20003, 40001, 40002 | Boolean |
| Systemic lupus erythematosus? | ICD10: M32  NCIC: 1381 | No \| Yes | 41270, 42171, 20003, 40001, 40002 | Boolean |
| Severe mental illness? (schizophrenia, bipolar disorder and moderate/severe depression) | ICD10: F03, F068, F09, F20, F22, F23, F259, F28, F29, F31, F333,F39, F53  ICD9: 2950, 2951, 2952, 2953, 2956, 2963  NCIC: 1289, 1291 | No \| Yes | 41270, 42171, 20003, 40001, 40002 | Boolean |
| On atypical antipsychotic medication? | 1140867420,1140867432,1140867444,1140927956,1140927970,1140928916,1141152848,1141152860,1141153490,1141167976,1141177762,1141195974,1141202024 | No \| Yes | 20003 | Boolean |
| On regular corticosteroids? | 1140874790, 1140874816, 1140874896, 1140874930,1140874976, 1141145782, 1141173346 | No \| Yes | 20003 |  |
| Diagnosis of or treatment for erectile disfunction? | ICD10: N484,  NCIC: 1518,  medication: 1141168936, 1141168948, 1141168944, 1141168946, 1140869100, 1140883010 | No \| Yes | 41270, 42171, 20003, 40001, 40002, 20003 | Boolean |
| Cholesterol/HDL ratio |  | ratio | 30690, 30760 | Log-standardscaling (mean=0, std=1) |
| Mean SBP |  | mmHg | 4080, 93 | Log-standardscaling (mean=0, std=1) |
| Standard deviation of at least two most recent SBP readings |  | mmHg | 4080, 93 | Log-standardscaling (mean=0, std=1) |
| BMI |  | kg/m² | 21001 | Log-standardscaling (mean=0, std=1) |

**Supplementary Table 15: Predictors: QRISK3:** The table details each predictor along with its measurement unit, coding scheme, data source, corresponding UKB Field ID, and any transformation applied prior to modelling. All transformations were estimated within the training fold and then applied to the held-out data to avoid information leakage.

| **Predictor** | **Data-coding** | **Response options/unit** | **Field ID** |
| --- | --- | --- | --- |
| Age at recruitment |  | years | 21022 |
| Sex |  | Male \| Female | 31 |
| Smoking status |  | Non-smoker \| Ex-smoker \| Light smoker \| Moderate smoker \| Heavy smoker | 20116 |
| Heart disease in father/mother/siblings |  | No \| Yes | 20107, 20110, 20111 |
| Ethnicity |  | White or not stated \| Indian \| Pakistani \| Bangladeshi \| Other Asian \| Black Caribbean \| Black African \| Chinese \| Other ethnicity | 21000 |
| BMI |  | kg/m² | 21001 |
| Cholesterol/HDL ratio |  | ratio | 30690, 30760 |
| Mean SBP |  | mmHg | 4080, 93 |
| Mean DBP |  | mmHg | 4079, 94 |
| GPPAQ | Inactive: no job-related physical activity and 0 hours/week moderate or vigorous activity  Moderately inactive: lightly active job or job involves walking/standing and <1 hour/week activity  Moderately active: job involves physical work or standing and 1–3 hours/week activity  Active: physical job or ≥3 hours/week moderate or vigorous activity  Ties: Highest Level | Inactive \| Moderately inactive \| Moderately active \| Active | 806, 816, 884, 894, 904, 914 |
| AUDIT score | AUDIT-C Q1: alcohol drinking frequency: never = 0, monthly or less = 1, 2–4 times/month = 2, 2–3 times/week = 3, ≥4 times/week = 4  AUDIT-C Q2: average alcohol units per day: 0–2 units = 0, >2–4 units = 1, >4–6 units = 2, >6–9 units = 3, >9 units = 4  AUDIT-C score: (Q1 + Q2) × 1.5  AUDIT-C result: positive if score ≥5, negative if score <5 | score of 0-12 | 1568/1578/1588/1598/1608 |
| Cardiovascular risk score (QRISK3) | QRISK3 | risk in% | Refer to QRISK3 tab |

**Supplementary Table 16: Predictors: NHS-HC:** The table details each predictor along with its measurement unit, coding scheme, data source, corresponding UKB Field ID, and any transformation applied prior to modelling. All transformations were estimated within the training fold and then applied to the held-out data to avoid information leakage.

**Provided as separate Microsoft Excel file; Supplementary Table 17: External Validation: Predictor Mapping in EstBB:** The table details the predictors used within the validation effort and how they were mapped/replicated within EstBB. Due to its size, the table is provided as a separate Microsoft Excel file.

| **Endpoint** | **Cases** | **Controls** |
| --- | --- | --- |
| CAD | 1766 | 46220 |
| CVD | 5542 | 38032 |
| HF | 6587 | 37507 |
| ISS | 1769 | 47219 |
| PAD | 2948 | 45095 |

**Supplementary Table 17: External validation: Endpoint frequency in EstBB:** Case and Control numbers across the five assessed cardiovascular disease spectrum endpoints.

**Supplementary Figures**

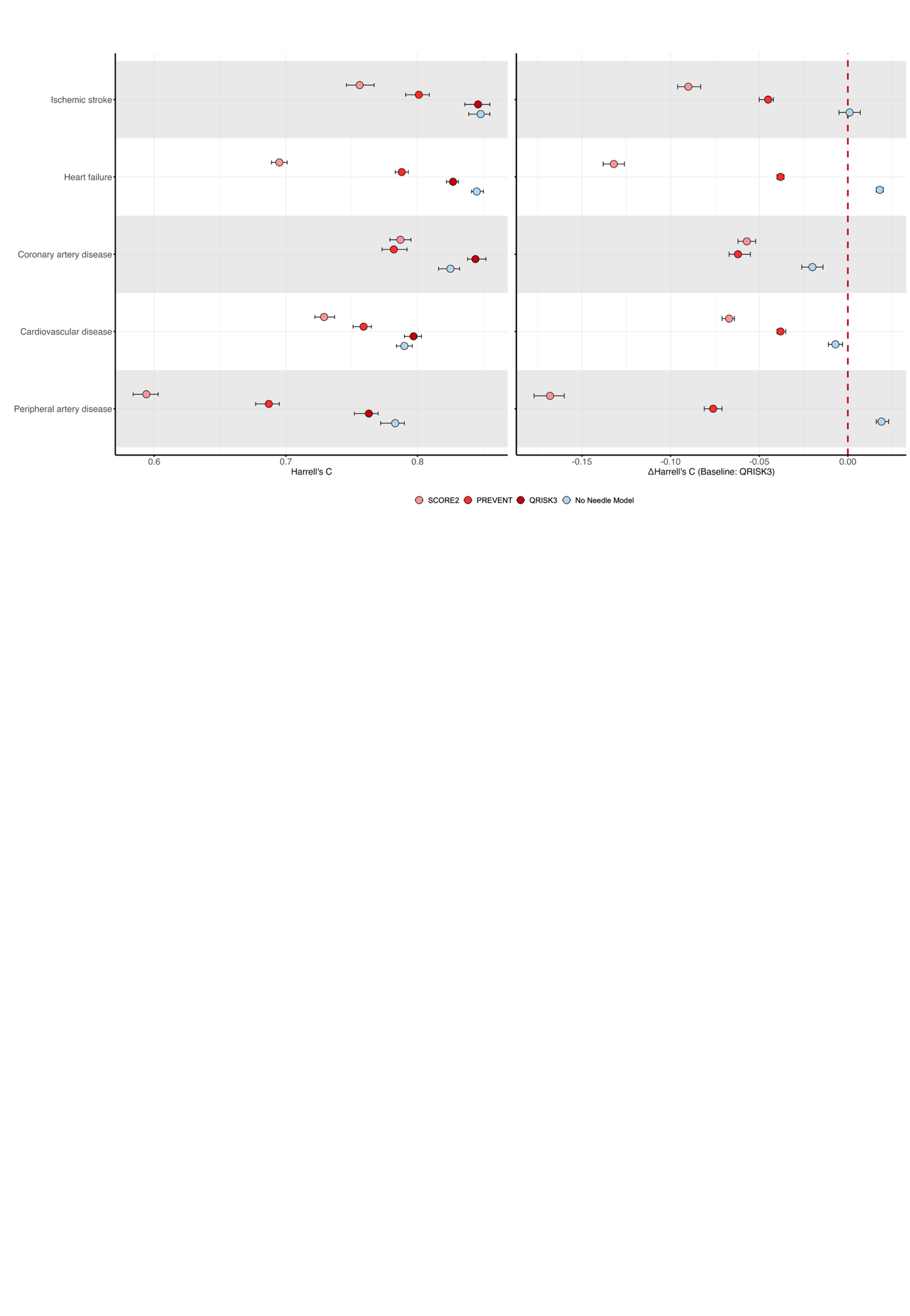

**Supplementary Figure 1: External validation of “No Needle Model”-based risk prediction in the Estonian Biobank:** Coefficients derived from the UK Biobank’s training partition were mapped to self-reported information within Estonian Biobank. We report both absolute (Harrell’s C-index) and relative (ΔHarrell’s C) performance within the Estonian Biobank subset.
